# Using human genetics to understand the effect of modulating targets of antihypertensive drugs in pregnancy

**DOI:** 10.64898/2026.05.12.26352361

**Authors:** Maria Carolina Borges, Helena Urquijo, Qian Yang, Adriaan van der Graaf, Nancy McBride, Eirin Beate Haug, Ana Goncalves Soares, Gemma C Clayton, Tom A Bond, Marwa Al Arab, Julie Horn, Laurent Thomas, Laxmi Bhatta, Bjørn Olav Åsvold, Maria C Magnus, David M Evans, Christy Burden, Katherine Birchenall, Ben Brumpton, Tom R Gaunt, Emma C Hart, Zoltan Kutalik, Deborah A Lawlor

**Affiliations:** MRC Integrative Epidemiology Unit at the University of Bristol, Bristol BS1 5DS, UK; Population Health Sciences, Bristol Medical School, University of Bristol, Bristol BS1 5DS, UK; Department of Endocrine and Metabolic Diseases, Shanghai Institute of Endocrine and Metabolic Diseases, Ruijin Hospital, Shanghai Jiao Tong University School of Medicine, Shanghai, China; Shanghai National Clinical Research Center for Metabolic Diseases, Key Laboratory for Endocrine and Metabolic Diseases of the National Health Commission of the PR China, Shanghai Key Laboratory for Endocrine Tumor, Lifecycle Health Management Center, Ruijin Hospital, Shanghai Jiao Tong University School of Medicine, Shanghai, China; Department of Computational Biology, University of Lausanne, Lausanne, Switzerland; HUNT Center for Molecular and Clinical Epidemiology, Department of Public Health and Nursing, Norwegian University of Science and Technology, Trondheim, Norway; Frazer Institute, University of Queensland, Brisbane, Australia; HUNT Research Centre, Department of Public Health and Nursing, NTNU, Norwegian University of Science and Technology, Levanger 7600, Norway; Department of Obstetrics and Gynecology, Levanger Hospital, Nord-Trøndelag Hospital Trust, Levanger, Norway; Division of Mental Health Care, St. Olavs Hospital, Trondheim, Norway; Department of Endocrinology, Clinic of Medicine, St. Olavs Hospital, Trondheim University Hospital, Trondheim, Norway; Centre for Fertility and Health, Norwegian Institute of Public Health, Oslo, Norway; Institute for Molecular Bioscience, University of Queensland, Brisbane, Australia; Translational Health Sciences, Bristol Medical School, University of Bristol, Bristol, UK; School of Medicine and Public Health, The University of Newcastle, Australia; Clinic of Medicine, St. Olavs Hospital, Trondheim University Hospital, Trondheim 7030, Norway; NIHR Biomedical Research Centre, University of Bristol, Bristol, UK; School of Physiology, Pharmacology and Neuroscience, University of Bristol, UK; Swiss Institute of Bioinformatics, Lausanne, Switzerland; University Center for Primary Care and Public Health, University of Lausanne, Lausanne, Switzerland

**Keywords:** Hypertensive disorders of pregnancy, pre-eclampsia, antihypertensives, drug target, drug safety, pregnancy, Mendelian randomization

## Abstract

**Background and Aims:** Hypertension during pregnancy is a major cause of maternal and neonatal morbidity and mortality, yet the efficacy and safety of antihypertensive treatments in this setting remain uncertain. We evaluated the effects of antihypertensive drug targets on adverse pregnancy-related outcomes using genetic variants to instrument target perturbation.

**Methods:** We performed drug target Mendelian randomization to mimic pharmacological perturbation of targets from six commonly used antihypertensive drug classes, using data from up to 671,922 pregnant women. Genetic variants near drug target genes associated with systolic or diastolic blood pressure were selected as instruments. We estimated effects of target modulation on six primary and eight secondary pregnancy outcomes.

**Results:** Genetically instrumented downregulation of blood pressure through β-blocker (BB) and calcium-channel blocker (CCB) targets, particularly ADRB1 and CACNB2, was associated with a reduced risk of hypertensive disorders of pregnancy, including preeclampsia. For example, CACNB2-instrumented lowering corresponded to a 7% (95% CI: 5–9%) reduction in preeclampsia risk per 1 mmHg decrease in blood pressure. For most of the other targets, estimates were directionally consistent but imprecise. Across additional outcomes, effects varied by target, with suggestive evidence for reduced risks of miscarriage, preterm birth, small-for-gestational age birth, and labour induction, although these estimates were accompanied by substantial uncertainty.

**Conclusions:** These findings support a protective effect of BB and CCB targets on hypertensive disorders of pregnancy and highlight potential target-specific differences in safety. This work illustrates the value of Mendelian randomization in addressing clinical uncertainties where robust trial evidence is limited.

## INTRODUCTION

One in every 10 pregnant women experiences high blood pressure during pregnancy, either due to pre-existing or pregnancy-induced hypertension (1). The incidence of hypertensive disorders of pregnancy is increasing due to rising maternal age, obesity, and a growing burden of comorbidities (2). Hypertension during pregnancy is associated with a three to five-fold higher risk of pre-eclampsia, preterm birth, small-for-gestational age (SGA) babies and perinatal death, as well as a five to ten-fold higher risk of maternal death (3, 4).

Antihypertensive treatment is usually indicated for pregnant women with hypertension (≥ 140/90 mmHg) (5-7). A 2018 Cochrane systematic review of 58 randomised controlled trials (RCTs), including data from 5,909 women with mild to moderate hypertension (140-169/90-109 mmHg), found that the use of any antihypertensive drug in pregnancy halves the risk of an episode of severe hypertension. However, evidence for effects on other clinically important outcomes was unclear, largely due to the poor-to-moderate quality of included trials and limited power to assess key safety outcomes, such as pregnancy loss or fetal growth restriction (8). Following this, an open label RCT enrolling 2,408 women with mild chronic hypertension reported that receiving antihypertensive medications, as opposed to no treatment (unless severe hypertension developed), reduced risk of a composite outcome consisting of pre-eclampsia with severe features, medically indicated preterm birth, placental abruption and fetal/neonatal death (9, 10).

There is very limited evidence on the comparative benefits and safety of different antihypertensive drug classes for pregnancy-related outcomes (6). Several guidelines consider adrenergic receptor alpha-2 agonists (AdrRA) (methyldopa), β-blockers (BB) (labetolol) and calcium channel blockers (CCB) (nifedipine) safe treatment options in pregnancy (5-7). Evidence from the 2018 Cochrane systematic review indicates that BB and CCB may exhibit greater efficacy in preventing severe hypertension episodes in pregnancy compared to AdrRA (8). In contrast, a 2025 network meta-analysis of 23 RCTs (3,989 women) found no clear evidence of superiority among labetalol, methyldopa, and nifedipine in head-to-head comparisons for severe hypertension, although labetalol showed a modest advantage for preeclampsia and preterm birth outcomes (11). Other commonly employed antihypertensive agents, such as angiotensin-converting enzyme inhibitors (ACEi) and angiotensin II receptor blockers (ARB), are contraindicated during pregnancy based on safety concerns from limited evidence (5-7), including reports of increased risks of pregnancy loss and congenital anomalies from case reports and series, cohort studies, and animal models (12-16). Taken together, these inconsistencies and evidence gaps constrain evidence-based decision-making for antihypertensive therapy in pregnancy and may contribute to medication non-adherence, heightened maternal anxiety, and avoidable risks to both mother and fetus, particularly when first-line treatments fail or when contraindicated agents are used before pregnancy recognition.

Given the limited availability of large-scale, high quality RCTs testing the effects of antihypertensive treatments during pregnancy, human genetics can provide useful insights into their on-target effect on important clinical outcomes beyond hypertensive episodes, as well as into potential differences in the efficacy and adverse effects of different classes of antihypertensive drugs. Human genetic evidence supported two-thirds of Food and Drug Administration (FDA) approved drugs in 2021 (17) and offers advantages over using animal models, which might not be directly translatable to human health. Drug target Mendelian randomization harnesses the properties of human genetic variants affecting the expression or function of proteins, which constitute most drug targets, to predict the effect of pharmacologically perturbing these drug targets (18, 19).

Previous studies have used drug target Mendelian randomization to assess the efficacy and safety of antihypertensive drugs in the general population, indicating that downregulating antihypertensive drug targets resulted in the expected lower risk of cardiovascular disease for ACEi, CCB and BB targets (20-22). In contrast, applications of drug target Mendelian randomization in pregnancy remain limited, with existing studies often underpowered (23) and restricted in the range of drug targets and outcomes considered (24, 25). Available evidence suggests that genetically proxied downregulation of BB and CCB targets may reduce the risk of preeclampsia, while downregulation of the β1-adrenergic receptor (ADRB1), targeted by BB, has been associated with lower birthweight (25). However, early analyses did not account for fetal genetic effects. More recent work indicates that the link between ADRB1 and birthweight is largely explained by the correlation between maternal and fetal genotypes, underscoring the importance of accounting for offspring genotype when interpreting drug target Mendelian randomization in the context of pregnancy (23, 24).

This study aimed to improve evidence on (i) the potential effects of antihypertensive drug treatment on a range of maternal and fetal outcomes, and (ii) the comparative benefits and risks of different antihypertensive drug classes in pregnancy. To do that, we used drug target Mendelian randomization to mimic pharmacological perturbation of targets from six commonly used types of antihypertensive drugs. We included data from up to 671,922 pregnant women and explored effects on six primary outcomes (miscarriage, stillbirth, hypertensive disorders of pregnancy, pre-eclampsia, preterm birth, and SGA) and eight secondary outcomes (induction of labour, caesarean section [C-section], elective C-section, very preterm birth, low Apgar at 5 minutes, neonatal intensive care unit [NICU] admission, as well as two continuous traits: birthweight and gestational age).

## METHODS

### Study population

We used data from study participants contributing to the MR-PREG collaboration, which is focused on improving the evidence basis for prevention and treatment of adverse outcomes during pregnancy and the perinatal period (26). At the time this study was conducted, the MR-PREG collaboration included up to 671,922 women of predominantly European ancestry from five studies — i.e. Avon Longitudinal Study of Parents and Children (ALSPAC) (27, 28); Born in Bradford (BiB) (29); the Trøndelag Health Study (HUNT) (30-33); the Norwegian Mother, Father and Child Cohort Study (MoBa) (34, 35); and UK Biobank (UKB) (36) —, as well as summary data from FinnGen (release 12) (37) and previous genome-wide association study (GWAS) metanalyses for pre-eclampsia (38), preterm birth (39) and gestational age (39). Details on study-specific characteristics are provided in **Supplementary text** and **Supplementary table 1**.

### Antihypertensive drugs and corresponding targets

We selected seven classes of drugs recommended in recent guidelines (40-42) for the treatment of hypertension in non-pregnant individuals: alpha-blocker (AB), ACEi, aldosterone receptor antagonist (AlRA), ARB), BB, CCB and thiazide/thiazide-like diuretics (TZD). We also added an eighth drug class widely used for the treatment of hypertension in pregnancy: AdrRA (5, 43). We identified a commonly used drug in each class, which was then mapped to their corresponding target(s) using the DrugBank database (44) (https://go.drugbank.com/). Targets were selected if marked as interacting directly with the drug as part of its mechanism of action (**Table 1** and **Supplementary table 2**).

**Table 1.** List of eligible drugs and corresponding targets

| Drug class | Drug class (full name) | Drug class ATC<sup>1</sup> | Examples of drugs in the class | Drug target(s)<sup>2</sup> | UniProt ID | Recommendation in pregnancy<sup>3</sup> |
| --- | --- | --- | --- | --- | --- | --- |
| AB | Alpha-1 adrenergic receptor antagonist (aka alpha-blocker) | C02CA | Prazosin | ADRA1A<br>ADRA1B<br>ADRA1D | P35348<br>P35368<br>P25100 | No evidence of teratogenicity |
| ACEi | Angiotensin-converting enzyme inhibitors | C09AA | Enalapril | ACE | P12821 | Contraindicated |
| AdrRA | Alpha-2 adrenergic receptor agonist | C02AB | Methyldopa | ADRA2A | P08913 | Methyldopa not known to be harmful |
| AlRA | Aldosterone receptor antagonist | C03DA | Spironolactone | NR3C2 | P08235 | Use only if potential benefit outweighs risk |
| ARB | Angiotensin II receptor blockers | C09CA | Losartan | AGTR1 | P30556 | Contraindicated |
| BB | beta-blockers | C07A | Labetolol<sup>4</sup> | ADRB1<br>ADRB2 | P08588<br>P07550 | Labetolol not known to be harmful, except possibly in 1st trimester |
| CCB | Calcium channel blockers (selective) | C08CA | Nifedipine | CACNA1C<br>CACNA1D<br>CACNB2 | Q13936<br>Q01668<br>Q08289 | Use only if other treatment options are not indicated or have failed |
| TZD | Thiazide or thiazide-like diuretics | C03AA | Hydrochlorothiazide | SLC12A3<br>KCNMA1 | P55017<br>Q12791 | Contraindicated |

<sup>1</sup> ATC: Anatomical Therapeutic Chemical classification system
<sup>2</sup> ACE: Angiotensin-converting enzyme; ADRA1A: Alpha-1A adrenergic receptor; ADRA1B: Alpha-1B adrenergic receptor; ADRA1D: Alpha-1D adrenergic receptor; ADRA2A: Alpha-2A adrenergic receptor; ADRB1: Beta-1 adrenergic receptor; ADRB2: Beta-2 adrenergic receptor; AGTR1: Type-1 angiotensin II receptor; CACNA1C: Voltage-dependent L-type calcium channel subunit alpha-1C; CACNA1D: Voltage-dependent L-type calcium channel subunit alpha-1D; CACNB2: Voltage-dependent L-type calcium channel subunit beta-2; KCNMA1: Calcium-activated potassium channel subunit alpha-1; NR3C2: Nuclear receptor subfamily 3 group C member 2; SLC12A3: Solute carrier family 12 member 3.
<sup>3</sup> Extracted from the British National Formulary (BNF) database: https://bnf.nice.org.uk/drugs/
<sup>4</sup> Labetolol acts as alpha- and beta-blocker, targeting ADRA1A in addition to ADRB1 and ADRB2.

### Study outcomes

Outcomes were selected to align, where possible, with the primary and secondary endpoints evaluated in the largest systematic review of randomized controlled trials of antihypertensive medications (8) (**Supplementary table 3**). We pre-specified six primary outcomes (miscarriage, stillbirth, any hypertensive disorder of pregnancy (HDP), pre-eclampsia, preterm birth and SGA) and six secondary outcomes (induction of labour, C-section, elective C-section, very preterm birth, low Apgar score at 5 minutes, NICU admission). In addition, we analysed two continuous traits, birthweight and gestational age, to increase statistical power and provide complementary information to related binary outcomes. Definitions of all study outcomes, alongside those used in the reference systematic review, are provided in **Supplementary Table 3**. Detailed descriptions of data sources and outcome definitions for contributing studies in the MR-PREG collaboration have been reported previously (45), and sample sizes for each analysis are summarised in **Table 2**.

**Table 2.** Number of cases and controls used for each outcome of interest

| Outcome | Number of cases | Number of controls | Total sample size |
| --- | --- | --- | --- |
| Miscarriage | 91,469 | 426,921 | 518,390 |
| Stillbirth | 6,826 | 227,641 | 234,467 |
| Hypertensive disorders of pregnancy | 32,549 | 509,219 | 541,768 |
| Pre-eclampsia | 19,408 | 652,584 | 671,992 |
| Pre-term birth | 18,225 | 267,497 | 285,722 |
| Small for gestational age | 11,874 | 109,340 | 121,214 |
| Induction of labour | 22,310 | 97,277 | 119,587 |
| Caesarean section | 33,909 | 200,060 | 233,969 |
| Elective C-section | 6,004 | 81,279 | 87,283 |
| Very pre-term birth | 1,765 | 100,596 | 102,361 |
| Low Apgar score at 5 minutes | 914 | 81,718 | 82,293 |
| NICU admission | 6,994 | 70,289 | 77,285 |
| Birth weight |  |  | 313,673 |
| Gestational age |  |  | 189,471 |

Our analytical approach consists of four stages summarised in **Figure 1**, as described in this section. All analyses were performed using R version 4.3.1. The analytical code is available at https://github.com/mcarolborges/mr_bp-drugs_appo/. This study follows the Strengthening the Reporting of Observational Studies in Epidemiology – Mendelian Randomization (STROBE-MR) reporting guidelines (46). The study protocol was not pre-registered.

**Figure 1.**
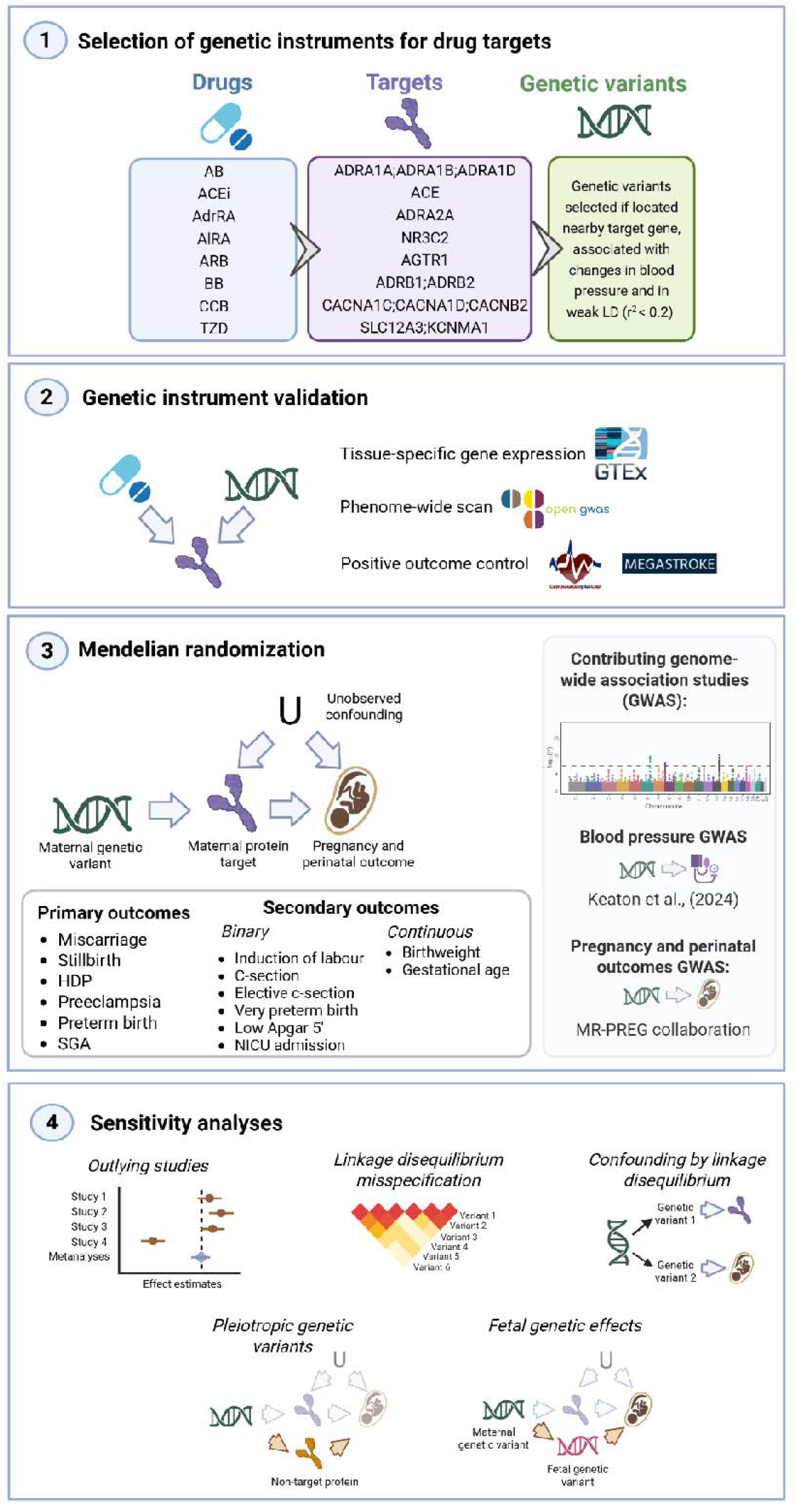
Schematic representation of the study analytical approach. Created with BioRender.com. AB: Alpha-blockers, ACEi: Angiotensin-converting enzyme inhibitors, AdrRA: Adrenergic receptor alpha-2 agonist, AlRA: Aldosterone receptor antagonist, ARB: Angiotensin II receptor blockers, BB: Beta-blockers, CCB: Calcium channel blockers, TZD: Thiazide or thiazide-like diuretics, HDP: hypertensive disorders of pregnancy, SGA, small-for-gestational age, GWAS: genome-wide association study.

#### 1. Selection of genetic instruments for drug targets

In the Supplementary methods, we give a full description of our procedure to select genetic instruments. In brief, our main criteria were based on:

i. location in the vicinity of the drug target encoding gene (aka cis-acting variants) – i.e. within a window of -/+ 500kb around the transcription start site of the target encoding gene
ii. potential impact on a downstream effect of the target – i.e. variants with strong evidence of association with mean changes in systolic (SBP) and/or diastolic blood pressure (DBP) at p < 5 × 10^−8^ in a GWAS metanalysis including 1,028,980 individuals of European ancestry (47)
iii. limited correlation (i.e. linkage disequilibrium) with other variants selected from the same window (r^2^ < 0.2 in the main analysis and r^2^<0.01 as sensitivity analysis)

For each selected variant, we estimated the proportion of variance explained in SBP or DBP (R^2^) and the instrument strength (F-statistics).

#### 2. Characterising genetic instruments

We took a combination of approaches to explore the validity of the chosen genetic variants as suitable instruments for perturbing the drug targets. Specifically, we aimed to explore whether the selected genetic variants were likely to influence blood pressure via their impact on the respective drug targets, were specific and had the expected effect on positive control outcomes.

First, we examined whether these cis-genetic variants were associated with changes in the expression of the corresponding target genes in key tissues involved in blood pressure regulation using genetic association data from the Genotype-Tissue Expression (GTEx) project (version 8) (48). The tissues analysed included artery [aorta (N = 387), coronary (N = 213), and tibial (N = 584)], adrenal gland (N = 233), heart [atrium (N = 372), and left ventricle (N = 386)], hypothalamus (N = 170), kidney cortex (N = 73), and lung (N = 515). Where we could detect suggestive statistical evidence for an association between genetic variant and changes in gene expression in a given tissue (p < 0.05), we further investigated whether the top selected genetic variant in each locus explained more variance in the expression of the target gene than in blood pressure using the Steiger test (which would be expected if the variant primarily affects gene expression) (49) and whether the association of genetic variants with changes in gene expression and blood pressure could be explained by shared causal variants using genetic colocalization (50) (using the same approach described under ‘Confounding by LD’ below). These would be indicative that the effect of a cis-genetic variant on blood pressure is mediated by changes in the expression of the target gene. Second, we conducted a phenome-wide association scan (PheWAS) to explore the specificity of the selected genetic variants against 50,055 traits in OpenGWAS using the ieugwasr R package version (51, 52) (access: 22^nd^ July 2025). We tested whether the genetic variants had the expected effect on two positive control outcomes in non-pregnant individuals – i.e., coronary artery disease and stroke – using publicly available GWAS data from CARDIOGRAM (60,801 cases and 123,504 controls (53) and MEGASTROKE (40,585 cases and 406,111 controls) (54).

#### 3. Mendelian randomization

We used two-sample Mendelian randomization to assess the effects of modulating these drug targets on adverse pregnancy and perinatal outcomes. We present odds ratios (OR) for binary outcomes, or mean change for continuous outcomes, per 1 mmHg decrease in SBP or DBP due to genetically instrumented downregulation of the drug target gene.

For these analyses, we used genetic association data with SBP and DBP from a previous GWAS metanalyses (47) and genetic association data with pregnancy and perinatal outcomes from the MR-PREG collaboration (26). Details on quality control, GWAS analyses and meta-analyses have been published previously for these data sources are described in detail elsewhere (26, 47).

For the main analyses, we used the Wald ratio, or the generalised inverse variance weighted (gIVW) estimators if, respectively, one or multiple genetic variants were selected for a given target. The gIVW is an extension of the standard inverse variance weighted (IVW) method that accounts for linkage disequilibrium between genetic instruments (55). We used a random subsample of 10,000 unrelated UK Biobank participants of European ancestry to estimate the extent of linkage disequilibrium between variants, indicated by the r^2^, as required by gIVW. These were undertaken using the TwoSampleMR and MendelianRandomization R packages (52, 56, 57). We selected results showing some evidence of association in the main analyses (p < 0.05) for further evaluation in sensitivity analyses, and also report results corrected for multiple testing using a 5% false discovery rate (FDR) control. However, our interpretation focuses on the direction and precision of effect estimates from the main analyses, as well as their consistency across the sensitivity analyses described below, to provide a more comprehensive assessment of the evidence.

### Sensitivity analyses

As with other statistical approaches, the validity of Mendelian randomization results relies on certain assumptions, the core ones can be summarized as follows:

I. Relevance: the instrument is statistically strongly associated with the exposure of interest in the relevant population (pregnant women, in this study).
II. Independence: there should be no confounding between the instrument and outcome.
III. Exclusion restriction criteria: any effect of the instrument on the outcome is fully mediated by the exposure.

The relevance assumption can be formally tested, whilst violations of the independence and exclusion restriction assumptions cannot be directly verified and, therefore, their plausibility was assessed through sensitivity analyses (58).

To assess instrument strength (relevance), we approximated the F-statistics as described above under *‘Selection of genetic instruments for drug targets’*. Because large datasets with blood pressure measurements during pregnancy are not available, we assumed that the effects of the selected genetic instruments on blood pressure are comparable in and outside of pregnancy. Supporting this assumption, a previous Mendelian randomization study demonstrated that genetic variants strongly associated with blood pressure in the general population predicted blood pressure changes during pregnancy in the expected direction, albeit in a small but informative sample (59).

We conducted extensive sensitivity analyses to evaluate the plausibility of the exclusion restriction assumption, particularly examining potential bias due to linkage disequilibrium, pleiotropy, and fetal effects, as well as assessing the influence of outlier studies and linkage disequilibrium misspecification, as described below.

#### Outlying studies

We conducted leave-one-study-out analysis to explore potential bias due to outlying studies. First, we performed Mendelian randomization for each study separately using the same methods as in the main analyses. Second, we combined study-specific estimates using fixed effect metanalyses with gIVW after removing one study at a time.

#### Linkage disequilibrium misspecification

To assess whether results from the main analyses (gIVW) were biased by our criteria for variant selection and misspecifications of linkage disequilibrium from the reference population, we repeated analyses using IVW after using a more stringent criterion for selecting genetic variants (linkage disequilibrium clumping threshold: r^2^ < 0.01 instead of r^2^ < 0.2).

#### Confounding by linkage disequilibrium

For each drug target genomic region, we performed genetic colocalization, using the coloc R package (50), to investigate whether Mendelian randomization findings are due to shared or distinct causal genetic variants between blood pressure and pregnancy outcome. The presence of a shared variant is a necessary, although not sufficient, condition for a valid Mendelian randomization finding, whereas the presence of distinct variants would be indicative that findings are confounded by linkage disequilibrium. Coloc uses a Bayesian approach to calculate support for five causal models (H0: no association; H1: association with trait 1 only; H2: association with trait 2 only; H3: association with both traits owing to distinct causal variants; H4: association with both traits owing to a single shared causal variant). We restricted coloc analysis to a genomic region within a 500 kb window around the target gene transcription start site and assumed prior probabilities that any random single nucleotide polymorphism (SNP) in the region is associated with trait 1 (p = 1 × 10^−4^), trait 2 (p = 1 × 10^−4^) or both traits (p = 1 × 10^−5^). A posterior probability of association (PPA) for H4 ≥-70% was considered as evidence supporting colocalization, while PPA-≥-70% for H3 was considered as evidence against colocalization. In cases where the combined PPA for H3 and H4 was lower than 70%, which is indicative of low power for **coloc**, we performed additional sensitivity analyses by relaxing the prior for both traits (p = 1 × 10^−4^). This is justified by the fact that these analyses are based on candidate Mendelian randomization results in contrast to the hypothesis-free context in which coloc was initially proposed.

#### Pleiotropic genetic variants

Our main findings may be biased if selected genetic variants influence genes other than the intended drug target, thereby affecting pregnancy or perinatal outcomes through alternative pathways. To assess the potential impact of horizontal pleiotropy, we applied two cis-Mendelian randomization methods designed to be more robust to such effects. First, we used MR-Link2, a likelihood-based approach that jointly estimates the causal effect and the variance attributable to horizontal pleiotropy. Validation studies indicate that MR-Link2 achieves high precision, albeit with reduced recall, relative to other cis-MR methods (60). Second, we applied Generalised Summary-data-based Mendelian Randomisation (GSMR2), which detects and removes invalid instruments (i.e. defined as variants with outcome effects disproportionate to their exposure effects) while accounting for linkage disequilibrium among the remaining variants (61). Instruments were selected using p < 5 × 10^−8^ and r^2^ < 0.6. Horizontal pleiotropy was assessed using the HEIDI (Heterogeneity In Dependent Instruments) test, with a threshold of p < 0.01 to exclude potentially invalid instruments.

#### Fetal genetic effects

We accounted for a potential effect from fetal genetic variants using a weighted linear model (WLM) (62) implemented in the DONUTS R package (63). First, we used the WLM to estimate conditional genetic effects – i.e. maternal genetic effects on each outcome adjusted for offspring genotype and, for comparison, fetal genetic effects adjusted for maternal genotype. Next, we repeated the gIVW using the maternal (and fetal) conditional genetic effects on each outcome as inputs. As an additional sensitivity analyses, we further repeated this procedure using estimates of maternal genetic effects on outcomes adjusted for both offspring and paternal genotypes, to control for potential bias due to assortative mating or spurious associations between maternal and paternal variants once fetal genotype is controlled for.

## RESULTS

### Genetic instruments strength and validity

We identified genetic instruments proxying perturbation of 11 antihypertensive drug targets; no variants met inclusion criteria for ADRA2A, KCNMA1, or NR3C2. The number of instruments per target ranged from 1 to 25, with individual SNPs explaining 0.002–0.03% of blood pressure variance and indicating adequate instrument strength (F-statistics 30–408) (**Supplementary tables 4** and **5**). For most targets, blood pressure-lowering alleles of lead variants were associated with either no detectable change or reduced gene expression, most commonly in cardiovascular-relevant tissues, although opposing effects in other tissues, most commonly lung, suggest potential tissue- or cell-specific regulation (**Supplementary figure 1**). Steiger filtering supported the expected direction of effect, with variants explaining more variance in gene expression than in blood pressure (**Supplementary table 6**), and colocalisation analyses generally indicated shared causal variants, with limited inconclusive cases and one instance consistent with distinct signals [*ACE* (lung)] (**Supplementary table 7**). Phenome-wide association analyses showed that variants primarily associated with blood pressure and related traits in the anticipated direction, with weaker secondary associations across diverse phenotypes (**Supplementary figure 2** and **Supplementary table 8**). In positive control analyses, genetically proxied blood pressure reduction via most targets associated with lower risk of stroke and/or coronary artery disease, although heterogeneity was observed for specific targets, including an increased stroke risk for ADRA1B (**Supplementary table 9**). A detailed description of findings exploring instrument validity are provided in the Supplementary text.

### Mendelian randomization

#### Primary outcomes

Genetically instrumented lowering of blood pressure via BB and CCB targets, particularly ADRB1 and CACNB2, was associated with a reduced risk of HDP and preeclampsia. For illustration, ORs for preeclampsia were 0.85 (95% confidence interval (CI): 0.79, 0.92) for ADRB1 and 0.93 (95% CI: 0.91, 0.95) for CACNB2 per 1 mmHg decrease in SBP or DBP. Other BB and CCB targets showed mostly directionally consistent protective associations, although estimates were imprecise. For the remaining primary outcomes, we observed suggestive evidence of a lower risk of miscarriage (ADRB1; OR 0.97; 95% CI: 0.95, 1.00) and preterm birth (CACNA1C; OR 0.74; 95% CI: 0.60, 0.92), and a higher risk of stillbirth (CACNB2; OR 1.04; 95% CI: 1.01, 1.07); however, these did not withstand multiple testing correction (**Figure 2A** and **Supplementary table 10**).

**Figure 2A.**
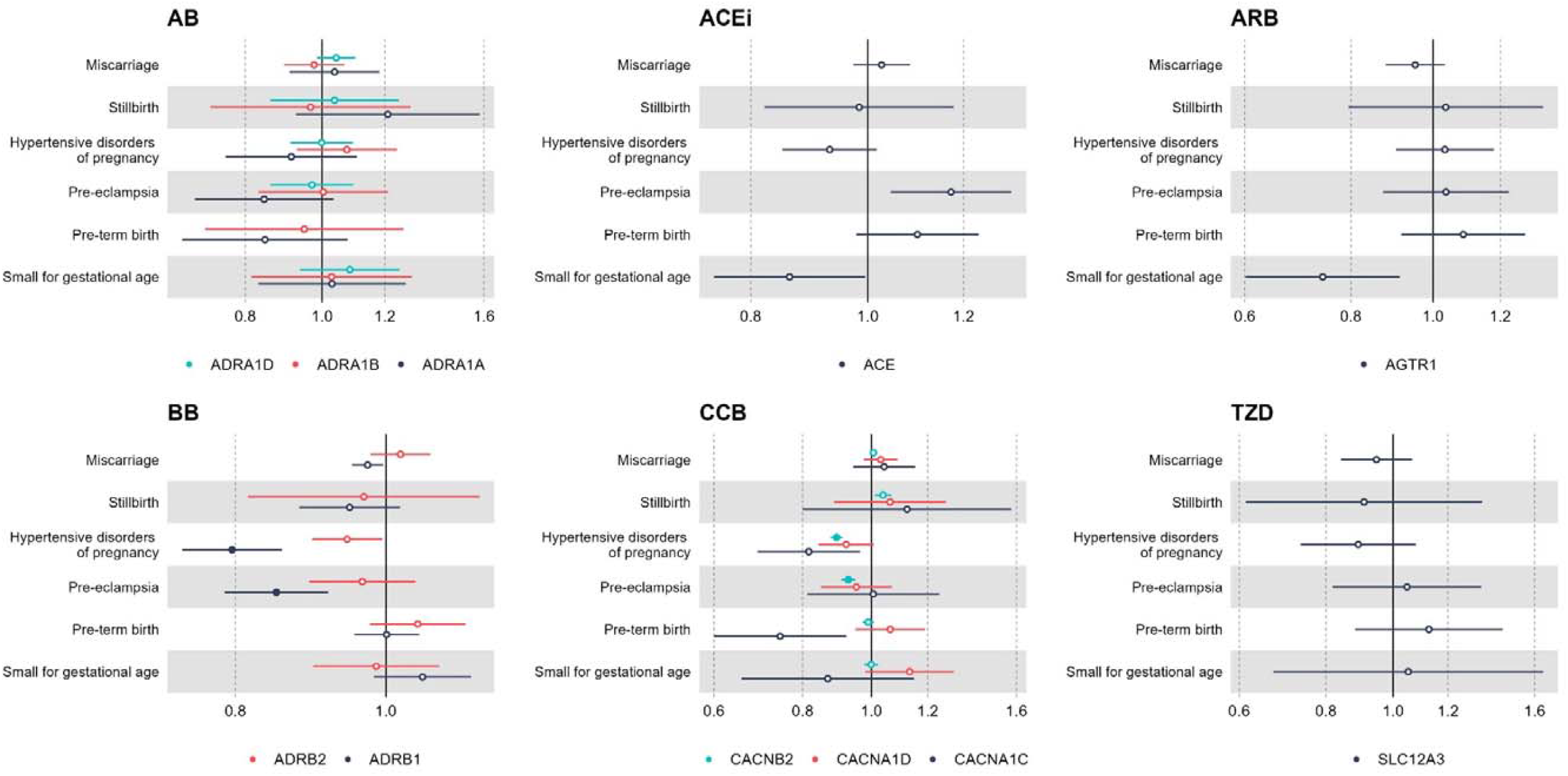
Mendelian randomization estimates for the effect of modulating hypertension drug targets on primary pregnancy-related outcomes Results are expressed as odds ratio (OR) of outcome per 1 mmHg decrease in systolic or diastolic blood pressure (BP) due to genetically-instrumented downregulation of the drug target gene. Point estimates and 95% confidence intervals are represented by circles and bars, respectively. Filled circles indicate estimates with false discovery rate-corrected *p*values < 0.05. AB-alpha-blockers; ACE: Angiotensin-converting enzyme; ACEi: ACE inhibitors; ADRA1A: adrenergic receptor α1A; ADRA1B: adrenergic receptor α1B; ADRA1D: adrenergic receptor α1D; ADRB1: adrenergic receptor β1; ADRB2: adrenergic receptor β2; AGTR1: angiotensin II receptor type 1; ARB: angiotensin II receptor blocker; BB: beta-blockers; CACNA1C: Voltage-dependent L-type calcium channel subunit α1C; CACNA1D: Voltage-dependent L-type calcium channel subunit α1D; CACNB2: Voltage-dependent L-type calcium channel subunit β2; CCB: calcium channel blockers; SLC12A3: Solute carrier family 12 member 3; TZD: thiazide or thiazide-like diuretics.

**Figure 2B.**
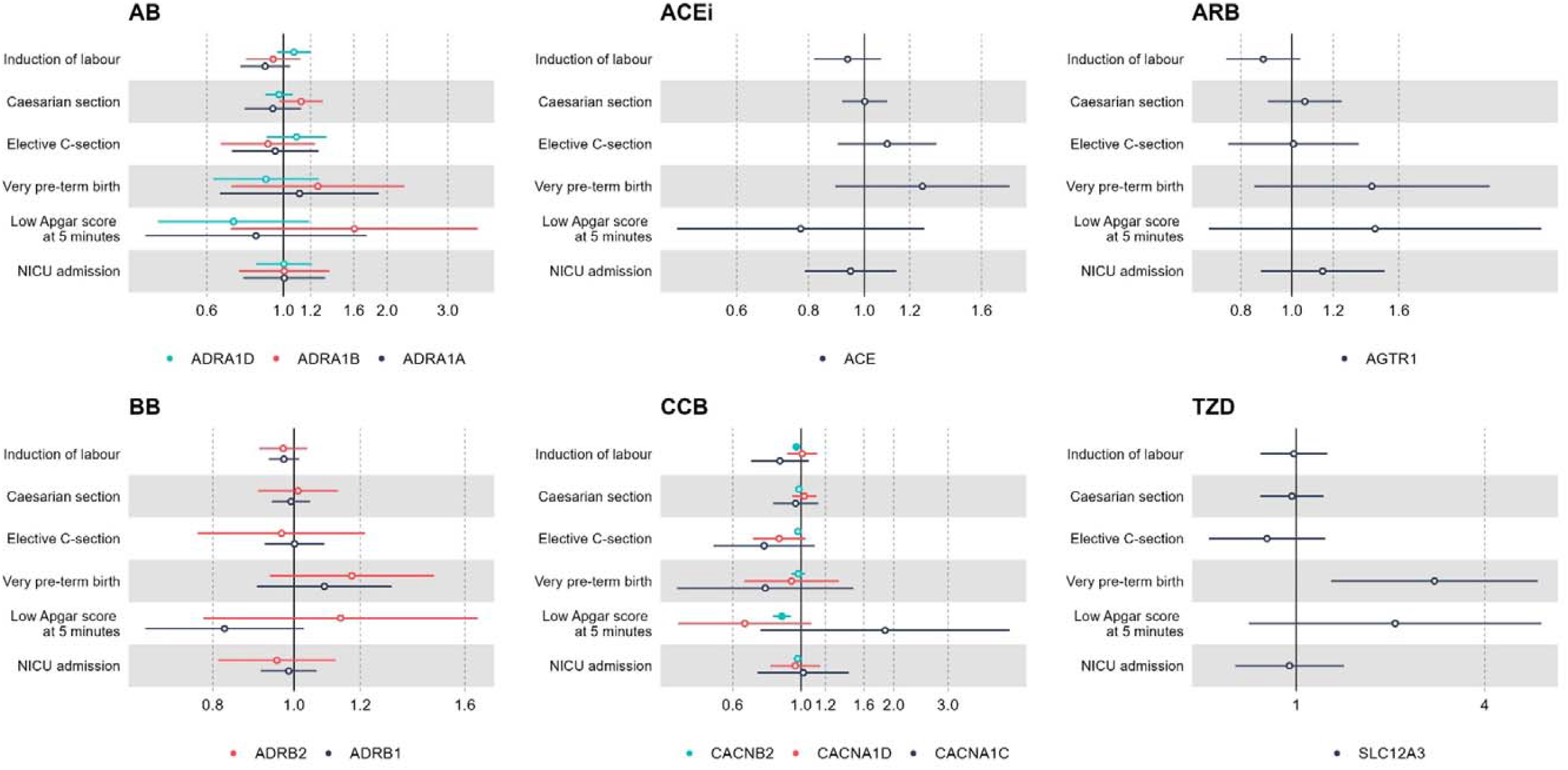
Mendelian randomization estimates for the effect of modulating hypertension drug targets on secondary binary pregnancy-related outcomes Results are expressed as odds ratio (OR) of outcome per 1 mmHg decrease in systolic or diastolic blood pressure (BP) due to genetically-instrumented downregulation of the drug target gene. Point estimates and 95% confidence intervals are represented by circles and bars, respectively. Filled circles indicate estimates with false discovery e-corrected p-values < 0.05. AB-alpha-blockers; ACE: Angiotensin-converting enzyme; ACEi: ACE inhibitors; ADRA1A: adrenergic receptor α1A; ADRA1B: adrenergic receptor α1B; ADRA1D: adrenergic receptor α1D; ADRB1: adrenergic receptor β1; ADRB2: adrenergic receptor β2; AGTR1: angiotensin II receptor type 1; ARB: angiotensin II receptor blocker; BB: beta-blockers; CACNA1C: Voltage-dependent L-type calcium channel subunit α1C; CACNA1D: Voltage-dependent L-type calcium channel subunit α1D; CACNB2: Voltage-dependent L-type calcium channel subunit β2; CCB: calcium channel blockers; SLC12A3: Solute carrier family 12 member 3; TZD: thiazide or thiazide-like diuretics.

For other drug classes (i.e. AB, ACEi, ARB, and TZD), we did not observe a consistent reduction in the risk of HDP and preeclampsia. Some estimates were directionally suggestive of benefit but imprecise, including HDP for ACE (OR 0.93; 95% CI: 0.85, 1.02), SLC12A3 (OR 0.89; 95% CI: 0.74; 1.08), and ADRA1A (OR 0.91; 95% CI: 0.76; 1.11), and preeclampsia for ADRA1A (OR 0.85; 95% CI: 0.69; 1.03). Most other estimates were close to the null with wide confidence intervals. One exception was a potential increase in preeclampsia risk associated with ACE downregulation (OR 1.17; 95% CI: 1.04; 1.31). We also observed suggestive evidence of reduced risk of small-for-gestational-age birth for AGTR1 (OR 0.74; 95% CI: 0.60; 0.91) and ACE (OR 0.86; 95% CI: 0.75; 0.99); however, no estimates for these drug classes passed multiple testing correction (**Figure 2A** and **Supplementary table 10**).

#### Secondary outcomes

For the secondary outcomes, genetically lowered blood pressure through the CCB target CACNB2 was linked to a reduced risk of labour induction (OR 0.97; 95% CI: 0.95; 0.99) and low Apgar score at 5 minutes (OR 0.87; 95% CI: 0.81; 0.93). There was also some indication that lowering blood pressure via SLC12A3 increased the risk of very preterm birth (OR 2.76; 95% CI: 1.29; 5.90) (**Figure 2B**). When examining continuous outcomes to maximise statistical power (i.e. birthweight and gestational age as continuous variables), we observed that genetically lowered blood pressure through ADRA1D, AGTR, ADRB2, and CACNB2 were associated with higher birthweight with estimated effects ranging from 0.01 (95% CI: 0.01; 0.02) to 0.07 (95% CI: 0.03; 0.11) SD units. Additionally, CACNB2 was associated with higher gestational age (0.01 SD; 95% CI: 0.01; 0.02), while results for the other targets were uncertain given the wide confidence intervals (**Supplementary figure 3**). Effect estimates for SLC12A3 and very preterm birth, and for ADRB2 and birthweight did not withstand multiple testing correction.

### Sensitivity analyses

Results from sensitivity analyses are presented in **Figure 3, Table 3**, and **Supplementary figures 4-7**. In the text, we focus on the sensitivity analysis results of drug target and (primary or secondary) outcome pairs for which there was some evidence of an effect in the main Mendelian randomization findings (p < 0.05), as summarized in **Table 4**.

**Figure 3A.**
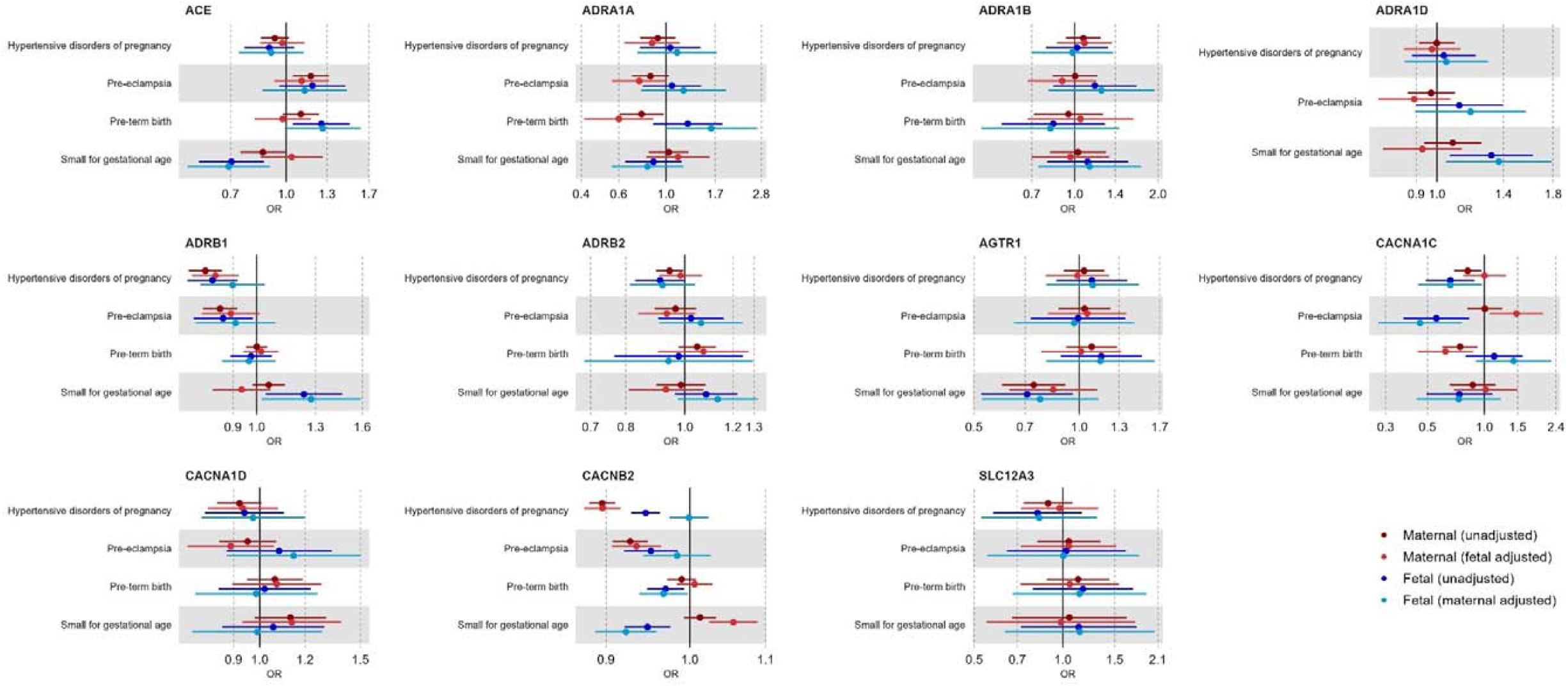
Mendelian randomization estimates for genetically-instrumented downregulation of drug targets on primary pregnancy-related outcomes based maternal genetic effects (unadjusted and adjusted for fetal genotype) and fetal genetic effects (unadjusted and adjusted for maternal genotype) Results are expressed as odds ratio (OR) of outcome per 1 mmHg unit decrease in systolic or diastolic blood pressure (BP) due to genetically-instrumented downregulation of the drug target gene. For comparison, we present the main results for maternal effects, unadjusted and unadjusted for offspring genotype, alongside results for fetal effects, unadjusted and unadjusted for maternal genotype. Point estimates and 95% confidence intervals are represented by circles and bars, respectively. ACE: Angiotensin-converting enzyme; ADRA1A: adrenergic receptor α1A; ADRA1B: adrenergic receptor α1B; ADRA1D: adrenergic receptor α1D; ADRB1: adrenergic receptor β1; ADRB2: adrenergic receptor β2; AGTR1: angiotensin II receptor type 1; CACNA1C: Voltage-dependent L-type calcium channel subunit α1C; CACNA1D: Voltage-dependent L-type calcium channel subunit α1D; CACNB2: Voltage-dependent L-type calcium channel subunit β2; SLC12A3: Solute carrier family 12 member 3.

**Figure 3B.**
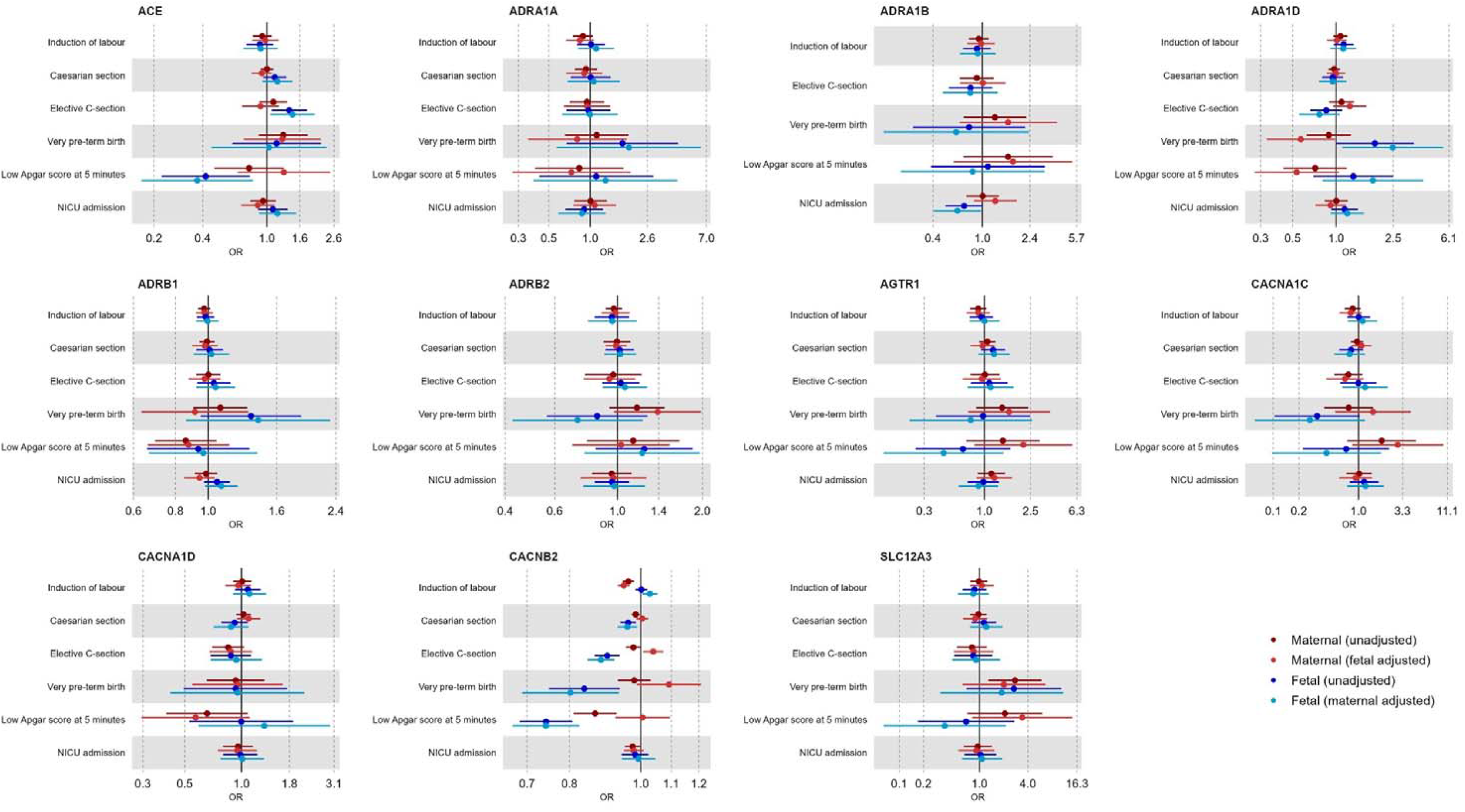
Mendelian randomization estimates for genetically-instrumented downregulation of drug targets on secondary pregnancy-related outcomes based on maternal genetic effects (unadjusted and adjusted for fetal genotype) and fetal genetic effects (unadjusted and adjusted for maternal genotype) Results are expressed as odds ratio (OR) of outcome per 1mmHg unit decrease in systolic or diastolic blood pressure (BP) due to genetically-instrumented downregulation of the drug target gene. For comparison, we present the main results for maternal effects, unadjusted and unadjusted for offspring genotype, alongside results for fetal effects, unadjusted and unadjusted for maternal genotype. Point estimates and 95% confidence intervals are represented by circles and bars, respectively. CE: Angiotensin-converting enzyme; ADRA1A: adrenergic receptor α1A; ADRA1B: adrenergic receptor α1B; ADRA1D: adrenergic receptor α1D; ADRB1: adrenergic receptor β1; ADRB2: adrenergic receptor β2; AGTR1: angiotensin II receptor type 1; CACNA1C: Voltage-dependent L-type calcium channel subunit α1C; CACNA1D: Voltage-dependent L-type calcium channel subunit α1D; CACNB2: Voltage-dependent L-type calcium channel subunit β2; SLC12A3: Solute carrier family 12 member 3.

**Table 3.**
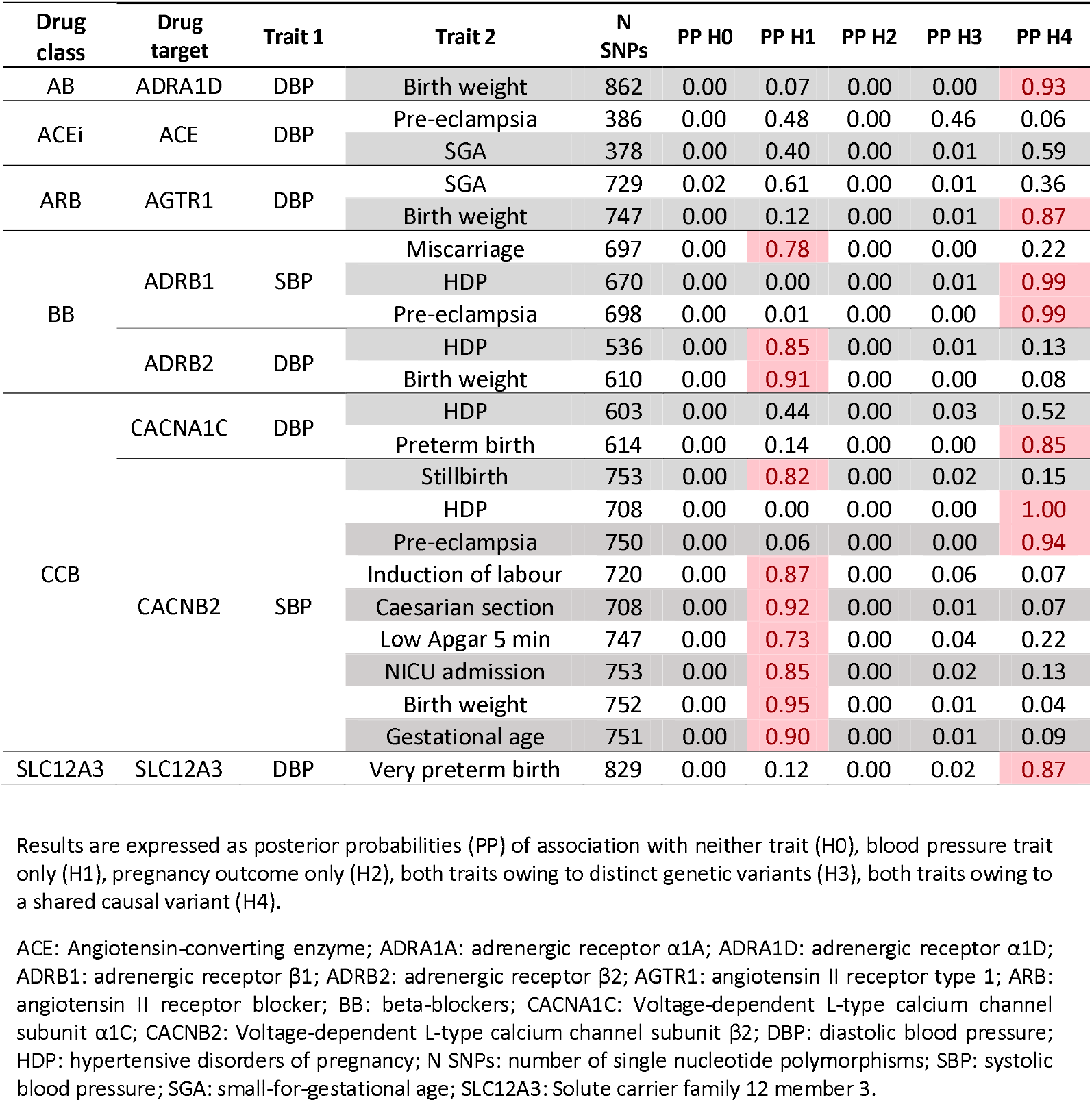
Genetic colocalization results for blood pressure traits and adverse pregnancy outcomes according to drug target gene region

**Table 4.**
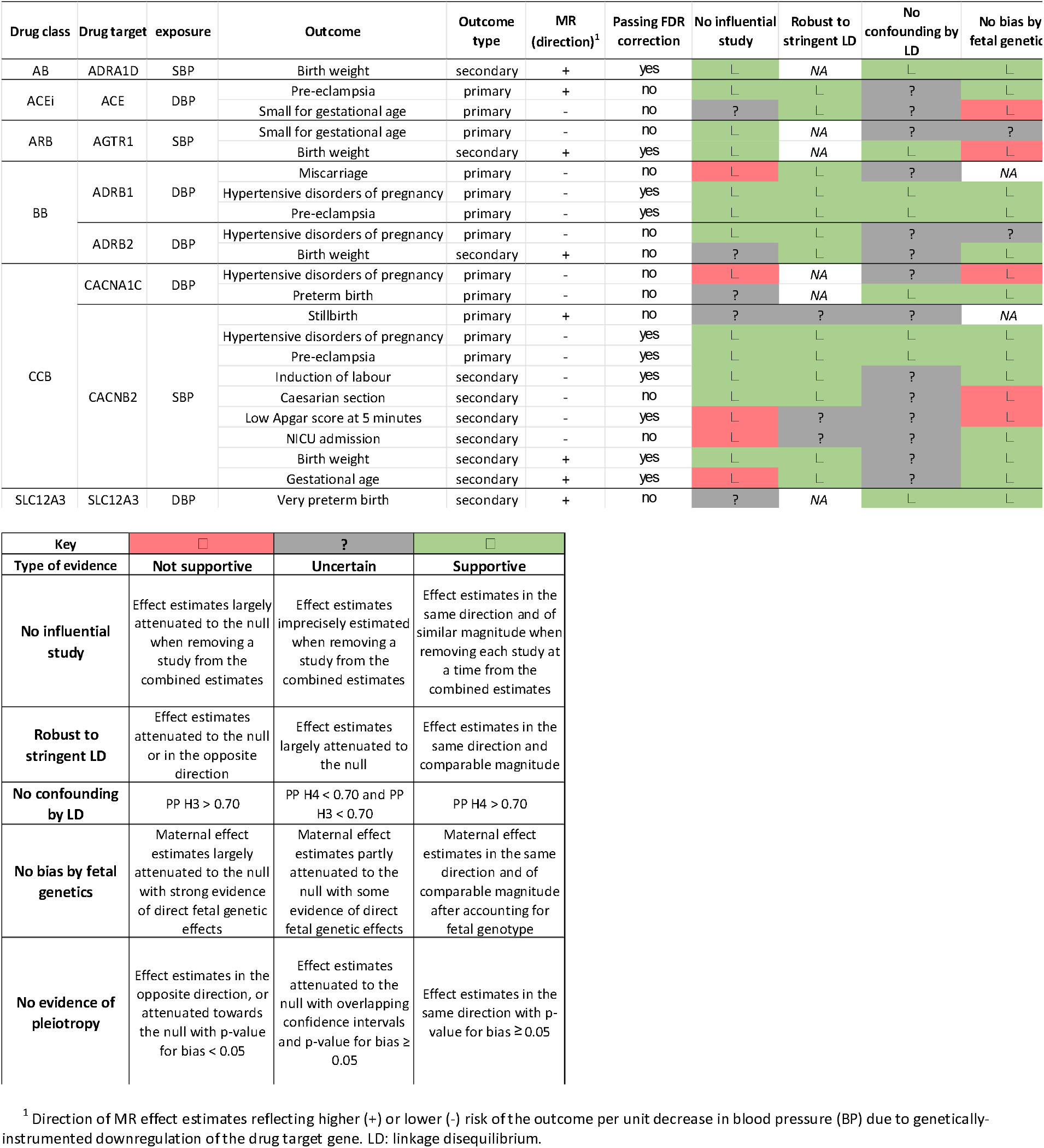
Summary of evidence from sensitivity analyses for MR results selected for follow-up

#### Assessing the presence of influential studies

Overall, we observed consistent results in the leave-one-study-out analyses, indicating that our main findings were not driven by a specific study. In a few instances, effect estimates from the main analyses were largely or completely attenuated when removing specific studies – i.e. ADRB1 with miscarriage (UK Biobank removed); CACNA1C with HDP (FinnGen removed); CACNB2 with low Apgar at 5 minutes, NICU admission, and gestational age (MoBa removed) **(Supplementary figure 4A-C, Table 4)**.

### Examining a more stringent threshold for variant selection

Findings from Mendelian randomization analyses using a more stringent linkage disequilibrium threshold for selecting genetic variants (R^2^ < 0.01 instead of R^2^ < 0.20) were largely similar to the main analyses apart from an expected decrease in the precision of estimates (**Supplementary figure 5A-C, Table 4**).

#### Checking for confounding by linkage disequilibrium

In genetic colocalization analyses, we observed evidence supportive of a shared genetic variant for ADRA1D (birth weight), AGTR1 (birth weight), ADRB1 (HDP and pre-eclampsia), CACNA1C (preterm birth), CACNB2 (HDP and pre-eclampsia), and SLC12A3 (very preterm birth) (PPA for H4: 85-100%). In other instances, evidence for colocalization was uncertain (PPA for H1: 40-95%) rather than supportive of distinct genetic variants (PPA for H3 for other comparisons < 6%, expect for ACE and pre-eclampsia where PPA H3 = 46%) (**Table 3-4, Supplementary figures 6A-G**).

#### Accounting for fetal genetic effects

Mendelian randomization results were generally similar when accounting for potential fetal genetic effects (**Table 4, Figure 4A-B** and **Supplementary figure 7**). In some instances, adjusted estimates were inconsistent with unadjusted estimates (ACE with small-for-gestational age, AGTR1 with birthweight, CACNA1C with hypertensive disorders of pregnancy, CACNB2 with caesarean section and low Apgar score at 5 minutes), indicating that the main findings were explained by fetal, rather than maternal, effects. We could not perform adjusted analyses for miscarriage and stillbirth due to the absence of data on fetal genotype. Maternal effect estimates were similar when additionally accounting for paternal genetics (**Supplementary figure 8A-C**).

#### Exploring horizontal pleiotropy

Overall, effect estimates from our main analyses using standard Mendelian randomization methods were in the same direction when using MR-Link2 and GSMR2. In addition, there was no strong evidence of pleiotropy for most findings, except for ACE with pre-eclampsia (i.e. MR-Link2 p-value for horizontal pleiotropy < 0.05) (**Supplementary figures 9A-C, Table 4**). Estimates across methods for the positive control outcomes are presented for comparison (**Supplementary figure 10**).

## DISCUSSION

Our study utilized drug target Mendelian randomization with large-scale population data to assess the effects of perturbing eleven targets across six classes of antihypertensive drugs on various adverse pregnancy-related outcomes. Overall, our findings contribute to addressing two critical gaps in the evidence from RCTs. First, our results support the notion that downregulating blood pressure via the targets of antihypertensive drugs impacts several adverse outcomes, including preeclampsia, induction, small-for-gestational age births, and preterm deliveries. Second, our study underscores potential differences in the efficacy and safety profiles across antihypertensive drug targets.

Antihypertensive treatment for women with mild to moderate hypertension during pregnancy remains controversial. A 2018 Cochrane systematic review of RCTs found that while antihypertensive drugs halve the risk of developing severe hypertension, their impact on maternal and fetal outcomes was unclear (8). Following this, the Chronic Hypertension and Pregnancy (CHAP) trial reported that, in pregnant women with mild chronic hypertension, antihypertensive treatment (AdrRA, BB, CCB classes) lowered the risk of a composite outcome including preeclampsia, medically indicated preterm birth, placental abruption, and fetal or neonatal death compared to those randomised to no treatment (10). Our previous Mendelian randomization study supports the notion that genetically predicted lowering of systolic and diastolic blood pressure using hundreds of genetic variants across the genome reduces the risk of preterm birth, SGA birth, induction of labour, and NICU admission, but did not identify any impact on miscarriage or stillbirth risk (25). Together, these studies suggest benefits of blood pressure reduction but leave uncertainty regarding heterogeneity across drug targets and specific outcomes.

In line with the CHAP trial, our findings indicate that downregulating targets of BB and CCB is related to lower risk of preeclampsia (ADRB1, CACNB2), preterm birth (CACNA1C), and miscarriage (ADRB1), although findings for miscarriage were strongly influenced by a single study. Unexpectedly, we also identified a potential adverse association between CACNB2 downregulation, proxying CCB use, and stillbirth; however, this finding requires replication given evidence was broadly uncertain from sensitivity analyses and it did not withstand correction for multiple testing. Moreover, our results highlight potential beneficial effects of downregulating BB and CCB targets on other outcomes, including birth weight (ADRB2, CACNB2), induction of labour (CACNB2), caesarean section (CACNB2), low Apgar score at 5 minutes (CACNB2), NICU admission (CACNB2), and gestational age (CACNB2), several of which have not been adequately assessed in previous RCTs, partly due to limited statistical power. Taken together, these findings highlight the value of well-powered, well-conducted RCTs and Mendelian randomization approaches for improving our understanding of the risks and benefits associated with antihypertensive drug use during pregnancy.

Some point estimates appeared relatively large given scaling per 1 mmHg change in systolic or diastolic blood pressure. For example, the odds ratio for preeclampsia was 0.85 (95% CI: 0.79, 0.92) for genetically instrumented downregulation of blood pressure via *ADRB1*, whereas the estimate for coronary artery disease, a positive control outcome, was 0.93 (95% CI: 0.89, 0.96 per 1 mmHg), consistent with prior Mendelian randomization studies (64, 65). These magnitude of the effect estimates should be interpreted cautiously. MR estimates rely on additional assumptions to estimate the magnitude of effect, the genetic instruments may not accurately capture blood pressure changes across gestation, and uncertainty remains, as reflected in the wide confidence intervals for most estimates.

A key insight from our analyses is that direction of effects potentially vary by drug target. We observed divergent effects of blood pressure lowering across targets for preeclampsia and fetal growth outcomes (birth weight and SGA birth). For preeclampsia, downregulation of most targets was associated with the expected reduction in risk, whereas downregulation of ACE showed an apparent increase in risk. This finding should be interpreted cautiously, as it may reflect bias due to linkage disequilibrium and pleiotropy. For fetal growth outcomes, downregulation of some targets was associated with higher birth weight (ADRA1D, AGTR1, ADRB2, CACNB2) or lower risk of SGA (ACE, AGTR1), while there was weaker evidence that downregulation of ADRB1 (−0.05 SD, 95% CI: −0.04, 0; p-value = 0.051) and SLC12A3 (−0.06 SD, 95% CI: −0.12, 0; p-value = 0.053) was associated with lower birth weight. Additional analyses indicated that several of these associations were driven by fetal rather than maternal genetic effects, including ACE with SGA, and ADRB1 and AGTR1 with birth weight. Prior Mendelian randomization work has similarly reported that variants in *ADRB1* associated with lower maternal blood pressure are linked to reduced birth weight (25); our findings extend this by suggesting that the effect is primarily mediated through fetal genotype, consistent with more recent studies (23, 24). Given that several antihypertensive drugs cross the placenta and may directly affect the fetus, the contribution of fetal genetic effects to these associations is clinically relevant.

Our study has several strengths. First, we leveraged large-scale population data, which increased statistical power to assess a broader range of clinically relevant outcomes than previous RCTs and improved the generalizability of our findings. Second, the use of germline genetic variants to mimic pharmacological perturbation of antihypertensive drug targets has important advantages. This approach helps mitigate bias from confounding by indication and reverse causation, which are common in observational studies. It avoids reliance on model organisms to infer effects of target perturbation in humans. It enables the investigation of the impact of modulating targets of drugs not yet approved for use in pregnancy without exposing pregnant women to these drugs. Third, we harnessed the family-based structure of some contributing studies to disentangle maternal and fetal genetic effects. Notably, apparent adverse effects of ARBs (AGTR1) and BBs (ADRB1) on fetal growth were attributable to fetal, rather than maternal, genetic influences. This underscores the importance of accounting for fetal genetic effects in Mendelian randomization studies involving mother– offspring pairs, as well as the complexity of interpreting such findings, particularly given that most drugs cross the placenta and may exert direct effects on the fetus (66).

However, there are several considerations when translating these findings to clinical settings. First, using common genetic variants to proxy pharmacological modulation of drug targets has inherent limitations. Common genetic variants around the drug target gene are likely to reflect modest effects on the target across the life course, and therefore, may not capture non-linear effects associated with drastic downregulation of the target by pharmacological inhibition, or pinpoint critical windows of exposure (e.g. specific trimesters in pregnancy). Moreover, the mechanism of action on the target might differ between genetic variants and drugs – e.g. ACE inhibitors inhibit the activity of the enzyme, while common genetic variants are more likely to act via influencing *ACE* gene expression. Second, despite the granularity and scale of our data, including for safety outcomes such as miscarriage and stillbirth, important limitations remain. These include limited statistical power for some outcomes (e.g. very preterm birth and low Apgar score at 5 minutes), the absence of key outcomes such as congenital anomalies and developmental disorders, and the lack of large datasets with blood pressure measurements during pregnancy to assess the impact of genetic instruments on blood pressure changes during gestation. In addition, reduced power for certain targets introduces uncertainty when comparing benefit–risk profiles across drug classes and lack of offspring genotype data for non-live births precludes assessing the influence of fetal genetic effects in findings for miscarriage and stillbirth.

Our Mendelian randomization study reinforces the idea that targeting some antihypertensive drug pathways can effectively reduce the risk of hypertensive disorders of pregnancy, including pre-eclampsia. Additionally, our findings suggest the presence of target-specific mechanisms that may influence certain outcomes, especially those related to fetal growth. This study underscores the potential of Mendelian randomization as a valuable tool to provide additional evidence on the benefits and risks of antihypertensive drug use in pregnancy. This is particularly crucial given the significant knowledge gaps and the scarcity of high-quality, well-powered RCTs including pregnant people.

## ACKNOWLEDGEMENTS

ALSPAC: We are extremely grateful to all the families who took part in this study, the midwives for their help in recruiting them, and the whole ALSPAC team, which includes data collection staff, data and administrations staff, technical managers and the technical staff with the Bristol Bioresource Laboratory, based within the University of Bristol

BiB: BiB is only possible because of the enthusiasm and commitment of the Children and Parents in BiB. We are grateful to all the participants, teachers, school staff, health professionals and researchers who have made BiB happen.

MoBa: This research has been conducted using MoBa data using application number 2552. MoBa is supported by the Norwegian Ministry of Health and Care services and the Ministry of Education and Research. We are grateful to all the participating families in Norway who take part in this on-going cohort study. We thank the Norwegian Institute of Public Health (NIPH) for generating high-quality genomic data. This research is part of the HARVEST collaboration, supported by the Research Council of Norway (#229624). We also thank the NORMENT Centre for providing genotype data, funded by the Research Council of Norway (#223273), South East Norway Health Authority and KG Jebsen Stiftelsen. We further thank the Center for Diabetes Research, the University of Bergen for providing genotype data and performing QC and imputation of the data funded by the ERC AdG project SELECTionPREDISPOSED, Stiftelsen Kristian Gerhard Jebsen, Trond Mohn Foundation, the Research Council of Norway, the Novo Nordisk Foundation, the University of Bergen, and the Western Norway health Authorities (Helse Vest).

FinnGen: The authors thank the FinnGen investigators for sharing their summary-level data.

UK Biobank: We would like to thank all the participants of UK Biobank for their vital contribution to the resource. This research has been conducted using the UK Biobank Resource under Application Number 23938.

HUNT: The Trøndelag Health Study (HUNT) is a collaboration between HUNT Research Center (Faculty of Medicine and Health Sciences, NTNU, Norwegian University of Science and Technology), Trøndelag County Council, Central Norway Regional Health Authority, and the Norwegian Institute of Public Health.

This work used the computational facilities of the Advanced Computing Research Centre, University of Bristol -http://www.bristol.ac.uk/acrc/. The current analysis was approved under UK Biobank Project 30418 and 81499.

## DATA AVAILABILITY STATEMENT

Genetic association data for the exposure are publicly available on the GWAS Catalog website data repository (https://www.ebi.ac.uk/gwas) with data accession codes GCST90310294 and GCST90310295 for SBP and DBP, respectively.

Genetic association data for the outcomes, generated by the MR-PREG collaboration, can only be used for research that is covered by data agreements with current contributing studies.

The ALSPAC access policy that describes the proposal process in detail including any costs associated with conducting research at ALSPAC, which may be updated from time to time, and is available at: https://www.bristol.ac.uk/media-library/sites/alspac/documents/researchers/data-access/ALSPAC_Access_Policy.pdf.

Data is available upon request from BiB, and information is available at: https://borninbradford.nhs.uk/research/how-to-access-data/-

Data from MoBa are available upon application to Helsedata administered by the Norwegian Institute of Public Health (see its website https://www.fhi.no/en/ch/studies/moba/for-forskere-artikler/research-and-data-access/ for details).

Data from the HUNT Study are available upon application to HUNT Research Centre, and information is available at: https://www.ntnu.edu/hunt/research

Researchers can apply for access to the UK Biobank data via the Access Management System (AMS) (https://www.ukbiobank.ac.uk/enable-your-research/apply-for-access).

## REFERENCES

1. Abalos E, Cuesta C, Grosso AL, Chou D, Say L. Global and regional estimates of preeclampsia and eclampsia: a systematic review. Eur J Obstet Gynecol Reprod Biol. 2013;170(1):1–7.

2. Garovic VD, Dechend R, Easterling T, Karumanchi SA, McMurtry Baird S, Magee LA, et al. Hypertension in Pregnancy: Diagnosis, Blood Pressure Goals, and Pharmacotherapy: A Scientific Statement From the American Heart Association. Hypertension. 2022;79(2):e21–e41.

3. Haug EB, Horn J, Markovitz AR, Fraser A, Vatten LJ, Macdonald-Wallis C, et al. Life Course Trajectories of Cardiovascular Risk Factors in Women With and Without Hypertensive Disorders in First Pregnancy: The HUNT Study in Norway. J Am Heart Assoc. 2018;7(15):e009250.

4. Bakker R, Steegers EA, Hofman A, Jaddoe VW. Blood pressure in different gestational trimesters, fetal growth, and the risk of adverse birth outcomes: the generation R study. Am J Epidemiol. 2011;174(7):797–806.

5. National Institute for Health and Care Excellence. Hypertension in pregnancy: diagnosis and management 2023 [Available from: https://www.nice.org.uk/guidance/ng133.

6. World Health Organization (WHO). WHO recommendations on drug treatment for non-severe hypertension in pregnancy. Geneva: WHO; 2020.

7. Mancia G, Kreutz R, Brunstrom M, Burnier M, Grassi G, Januszewicz A, et al. 2023 ESH Guidelines for the management of arterial hypertension The Task Force for the management of arterial hypertension of the European Society of Hypertension: Endorsed by the International Society of Hypertension (ISH) and the European Renal Association (ERA). J Hypertens. 2023;41(12):1874–2071.

8. Abalos E, Duley L, Steyn DW, Gialdini C. Antihypertensive drug therapy for mild to moderate hypertension during pregnancy. Cochrane Database Syst Rev. 2018;10(10):CD002252.

9. Tita AT, Szychowski JM, Andrews WW. Treatment for Mild Chronic Hypertension during Pregnancy. Reply. N Engl J Med. 2022;387(7):664.

10. Tita AT, Szychowski JM, Boggess K, Dugoff L, Sibai B, Lawrence K, et al. Treatment for Mild Chronic Hypertension during Pregnancy. N Engl J Med. 2022;386(19):1781–92.

11. Hup RJ, Damen JAA, Terstappen J, Klein Haneveld MJ, Terstappen F, Magee LA, et al. Oral antihypertensive treatment during pregnancy: a systematic review and network meta-analysis. Am J Obstet Gynecol. 2025;233(4):250–62.

12. Spaggiari E, Heidet L, Grange G, Guimiot F, Dreux S, Delezoide AL, et al. Prognosis and outcome of pregnancies exposed to renin-angiotensin system blockers. Prenat Diagn. 2012;32(11):1071–6.

13. Cooper WO, Hernandez-Diaz S, Arbogast PG, Dudley JA, Dyer S, Gideon PS, et al. Major congenital malformations after first-trimester exposure to ACE inhibitors. N Engl J Med. 2006;354(23):2443–51.

14. Velazquez-Armenta EY, Han JY, Choi JS, Yang KM, Nava-Ocampo AA. Angiotensin II receptor blockers in pregnancy: a case report and systematic review of the literature. Hypertens Pregnancy. 2007;26(1):51–66.

15. Saji H, Yamanaka M, Hagiwara A, Ijiri R. Losartan and fetal toxic effects. Lancet. 2001;357(9253):363.

16. Buttar HS. An overview of the influence of ACE inhibitors on fetal-placental circulation and perinatal development. Mol Cell Biochem. 1997;176(1-2):61–71.

17. Ochoa D, Karim M, Ghoussaini M, Hulcoop DG, McDonagh EM, Dunham I. Human genetics evidence supports two-thirds of the 2021 FDA-approved drugs. Nat Rev Drug Discov. 2022;21(8):551.

18. Smith GD, Lawlor DA, Harbord R, Timpson N, Day I, Ebrahim S. Clustered environments and randomized genes: a fundamental distinction between conventional and genetic epidemiology. PLoS Med. 2007;4(12):e352.

19. Hingorani AD, Kuan V, Finan C, Kruger FA, Gaulton A, Chopade S, et al. Improving the odds of drug development success through human genomics: modelling study. Sci Rep. 2019;9(1):18911.

20. Lian J, Shi X, Jia X, Fan J, Wang Y, Zhao Y, et al. Genetically predicted blood pressure, antihypertensive drugs and risk of heart failure: a Mendelian randomization study. J Hypertens. 2023;41(1):44–50.

21. Gill D, Georgakis MK, Koskeridis F, Jiang L, Feng Q, Wei WQ, et al. Use of Genetic Variants Related to Antihypertensive Drugs to Inform on Efficacy and Side Effects. Circulation. 2019;140(4):270–9.

22. Fan B, Zhang J, Zhao JV. Systematic review of Mendelian randomization studies on antihypertensive drugs. BMC Med. 2024;22(1):547.

23. Barry CS, Walker VM, Burden C, Havdahl A, Davies NM. Genetic Insights Into Perinatal Outcomes of Maternal Antihypertensive Therapy During Pregnancy. JAMA Netw Open. 2024;7(8):e2426234.

24. Ardissino M, Morley AP, Richards EMF, Zöllner J, Truong B, Williamson C, et al. Effects of Genetically-Proxied Antihypertensive Drug Targets on Preeclampsia and Birth Weight. medRxiv. 2026:2026.03.17.26346752.

25. Ardissino M, Slob EAW, Rajasundaram S, Reddy RK, Woolf B, Girling J, et al. Safety of beta-blocker and calcium channel blocker antihypertensive drugs in pregnancy: a Mendelian randomization study. BMC Med. 2022;20(1):288.

26. McBride N, Clayton GL, Goncalves Soares A, Yang Q, Bond TA, Taylor A, et al. Cohort profile: the Mendelian randomisation in pregnancy (MR-PREG) collaboration - improving evidence for prevention and treatment of adverse pregnancy and perinatal outcomes. BMJ Open. 2026;16(3):e103753.

27. Boyd A, Golding J, Macleod J, Lawlor DA, Fraser A, Henderson J, et al. Cohort Profile: the ‘children of the 90s’--the index offspring of the Avon Longitudinal Study of Parents and Children. Int J Epidemiol. 2013;42(1):111–27.

28. Fraser A, Macdonald-Wallis C, Tilling K, Boyd A, Golding J, Davey Smith G, et al. Cohort Profile: the Avon Longitudinal Study of Parents and Children: ALSPAC mothers cohort. Int J Epidemiol. 2013;42(1):97–110.

29. Wright J, Small N, Raynor P, Tuffnell D, Bhopal R, Cameron N, et al. Cohort Profile: the Born in Bradford multi-ethnic family cohort study. Int J Epidemiol. 2013;42(4):978–91.

30. Krokstad S, Langhammer A, Hveem K, Holmen TL, Midthjell K, Stene TR, et al. Cohort Profile: the HUNT Study, Norway. Int J Epidemiol. 2013;42(4):968–77.

31. Asvold BO, Langhammer A, Rehn TA, Kjelvik G, Grontvedt TV, Sorgjerd EP, et al. Cohort Profile Update: The HUNT Study, Norway. Int J Epidemiol. 2023;52(1):e80–e91.

32. Brumpton BM, Graham S, Surakka I, Skogholt AH, Loset M, Fritsche LG, et al. The HUNT study: A population-based cohort for genetic research. Cell Genom. 2022;2(10):100193.

33. Naess M, Kvaloy K, Sorgjerd EP, Saetermo KS, Noroy L, Rostad AH, et al. Data Resource Profile: The HUNT Biobank. Int J Epidemiol. 2024;53(3).

34. Magnus P, Irgens LM, Haug K, Nystad W, Skjaerven R, Stoltenberg C, et al. Cohort profile: the Norwegian Mother and Child Cohort Study (MoBa). Int J Epidemiol. 2006;35(5):1146–50.

35. Magnus P, Birke C, Vejrup K, Haugan A, Alsaker E, Daltveit AK, et al. Cohort Profile Update: The Norwegian Mother and Child Cohort Study (MoBa). Int J Epidemiol. 2016;45(2):382–8.

36. Sudlow C, Gallacher J, Allen N, Beral V, Burton P, Danesh J, et al. UK biobank: an open access resource for identifying the causes of a wide range of complex diseases of middle and old age. PLoS Med. 2015;12(3):e1001779.

37. Kurki MI, Karjalainen J, Palta P, Sipila TP, Kristiansson K, Donner KM, et al. FinnGen provides genetic insights from a well-phenotyped isolated population. Nature. 2023;613(7944):508–18.

38. Morgan L, McGinnis R, Steinthorsdottir V, Svyatova G, Zakhidova N, Lee WK, et al. InterPregGen: genetic studies of pre-eclampsia in three continents. Nor Epidemiol. 2014;24(1-2):141–6.

39. Sole-Navais P, Flatley C, Steinthorsdottir V, Vaudel M, Juodakis J, Chen J, et al. Genetic effects on the timing of parturition and links to fetal birth weight. Nat Genet. 2023;55(4):559–67.

40. Williams B, Mancia G, Spiering W, Agabiti Rosei E, Azizi M, Burnier M, et al. 2018 ESC/ESH Guidelines for the management of arterial hypertension: The Task Force for the management of arterial hypertension of the European Society of Cardiology and the European Society of Hypertension: The Task Force for the management of arterial hypertension of the European Society of Cardiology and the European Society of Hypertension. J Hypertens. 2018;36(10):1953–2041.

41. National Institute for Health and Care Excellence. Hypertension in adults: diagnosis and management 2023 [Available from: https://www.nice.org.uk/guidance/ng136.

42. National Institute for Health and Care Excellence. Hypertension 2023 [Available from: https://cks.nice.org.uk/topics/hypertension/.

43. Regitz-Zagrosek V, Roos-Hesselink JW, Bauersachs J, Blomström-Lundqvist C, Cífková R, De Bonis M, et al. 2018 ESC Guidelines for the management of cardiovascular diseases during pregnancy: The Task Force for the Management of Cardiovascular Diseases during Pregnancy of the European Society of Cardiology (ESC). European Heart Journal. 2018;39(34):3165–241.

44. Wishart DS, Feunang YD, Guo AC, Lo EJ, Marcu A, Grant JR, et al. DrugBank 5.0: a major update to the DrugBank database for 2018. Nucleic Acids Res. 2018;46(D1):D1074–D82.

45. McBride N, Clayton GL, Goncalves Soares A, Yang Q, Bond TA, Taylor A, et al. Cohort Profile: The Mendelian Randomization in Pregnancy (MR-PREG) collaboration - Improving evidence for prevention and treatment of adverse pregnancy and perinatal outcomes. medRxiv 2025032225324447. 2025.

46. Skrivankova VW, Richmond RC, Woolf BAR, Yarmolinsky J, Davies NM, Swanson SA, et al. Strengthening the Reporting of Observational Studies in Epidemiology Using Mendelian Randomization: The STROBE-MR Statement. JAMA. 2021;326(16):1614–21.

47. Keaton JM, Kamali Z, Xie T, Vaez A, Williams A, Goleva SB, et al. Genome-wide analysis in over 1 million individuals of European ancestry yields improved polygenic risk scores for blood pressure traits. Nat Genet. 2024;56(5):778–91.

48. Consortium GT. The GTEx Consortium atlas of genetic regulatory effects across human tissues. Science. 2020;369(6509):1318–30.

49. Hemani G, Tilling K, Davey Smith G. Orienting the causal relationship between imprecisely measured traits using GWAS summary data. PLoS Genet. 2017;13(11):e1007081.

50. Giambartolomei C, Vukcevic D, Schadt EE, Franke L, Hingorani AD, Wallace C, et al. Bayesian test for colocalisation between pairs of genetic association studies using summary statistics. PLoS Genet. 2014;10(5):e1004383.

51. Ben E, Matthew L, Tessa A, Yi L, Peter M, Jon H, et al. The MRC IEU OpenGWAS data infrastructure. bioRxiv. 2020:2020.08.10.244293.

52. Hemani G, Zheng J, Elsworth B, Wade KH, Haberland V, Baird D, et al. The MR-Base platform supports systematic causal inference across the human phenome. Elife. 2018;7.

53. Nikpay M, Goel A, Won HH, Hall LM, Willenborg C, Kanoni S, et al. A comprehensive 1,000 Genomes-based genome-wide association meta-analysis of coronary artery disease. Nat Genet. 2015;47(10):1121–30.

54. Malik R, Chauhan G, Traylor M, Sargurupremraj M, Okada Y, Mishra A, et al. Multiancestry genome-wide association study of 520,000 subjects identifies 32 loci associated with stroke and stroke subtypes. Nat Genet. 2018;50(4):524–37.

55. Burgess S, Dudbridge F, Thompson SG. Combining information on multiple instrumental variables in Mendelian randomization: comparison of allele score and summarized data methods. Stat Med. 2016;35(11):1880–906.

56. Yavorska OO, Burgess S. MendelianRandomization: an R package for performing Mendelian randomization analyses using summarized data. Int J Epidemiol. 2017;46(6):1734–9.

57. Broadbent JR, Foley CN, Grant AJ, Mason AM, Staley JR, Burgess S. MendelianRandomization v0.5.0: updates to an R package for performing Mendelian randomization analyses using summarized data. Wellcome Open Res. 2020;5:252.

58. Sanderson E, Glymour MM, Holmes MV, Kang H, Morrison J, Munafo MR, et al. Mendelian randomization. Nat Rev Methods Primers. 2022;2.

59. Morales-Berstein F, Goncalves-Soares A, Yang Q, McBride N, Bond T, Al Arab M, et al. Assessing the impact of maternal blood pressure during pregnancy on perinatal health: a wide-angled Mendelian randomization study. BMC Med. 2026;24(1):2.

60. van der Graaf A, Warmerdam R, Auwerx C e QC, Vosa U, Borges MC, et al. MR-link-2: pleiotropy robust cis Mendelian randomization validated in three independent reference datasets of causality. Nat Commun. 2025;16(1):6112.

61. Xue A, Zhu Z, Wang H, Jiang L, Visscher PM, Zeng J, et al. Unravelling the complex causal effects of substance use behaviours on common diseases. Commun Med (Lond). 2024;4(1):43.

62. Warrington NM, Hwang LD, Nivard MG, Evans DM. Estimating direct and indirect genetic effects on offspring phenotypes using genome-wide summary results data. Nat Commun. 2021;12(1):5420.

63. Wu Y, Zhong X, Lin Y, Zhao Z, Chen J, Zheng B, et al. Estimating genetic nurture with summary statistics of multigenerational genome-wide association studies. Proc Natl Acad Sci U S A. 2021;118(25).

64. Gill D, Georgakis MK, Zuber V, Karhunen V, Burgess S, Malik R, et al. Genetically Predicted Midlife Blood Pressure and Coronary Artery Disease Risk: Mendelian Randomization Analysis. J Am Heart Assoc. 2020;9(14):e016773.

65. Georgiou AN, Zagkos L, Markozannes G, Chalitsios CV, Asimakopoulos AG, Xu W, et al. Appraising the Causal Role of Risk Factors in Coronary Artery Disease and Stroke: A Systematic Review of Mendelian Randomization Studies. J Am Heart Assoc. 2023;12(20):e029040.

66. Reisenberger K, Egarter C, Sternberger B, Eckenberger P, Eberle E, Weissenbacher ER. Placental passage of angiotensin-converting enzyme inhibitors. Am J Obstet Gynecol. 1996;174(5):1450–5.

