## Supplementary text for "Using human genetics to understand the effect of modulating targets of antihypertensive drugs in pregnancy"

**SUPPLEMENTARY METHODS**

**Study population**

In this study, we used data from the MR-PREG collaboration (release 4), including up to 671,922 women from five cohort studies — i.e. Avon Longitudinal Study of Parents and Children (ALSPAC) (1, 2); Born in Bradford (BiB) (3); the Norwegian Mother, Father and Child Cohort Study (MoBa) (4, 5), UK Biobank (UKB) (6), and the Trøndelag Health Study (HUNT) —, and public data from FinnGen (7) and previous genome-wide association study (GWAS) metanalyses for pre-eclampsia (8), pre-term birth (9) and gestational age (9).

### *ALSPAC:* *The* Avon Longitudinal Study of Parents and Children

ALSPAC is a prospective birth cohort that started recruiting pregnant women resident in the former county of Avon (centred around the city of Bristol, Southwest England), with delivery dates between April 1991 and December 1992. A total of 14,541 women (ALSPAC-G0) were enrolled during pregnancy (14,676 fetuses) and gave birth to 14,062 live children (ALSPAC-G1). 7 years after the initial recruitment, a further 913 eligible children were enrolled. The total sample size for analyses using any data collected after the age of seven is therefore 15,447 pregnancies (14,833 unique mothers), resulting in 15,658 fetuses. Further details can be found in the published cohort profiles (1, 2). Please note that the study website contains details of all the data that is available through a fully searchable data dictionary and variable search tool (<http://www.bristol.ac.uk/alspac/researchers/our-data/>). Ethical approval for the study was obtained from the ALSPAC Ethics and Law Committee and the Local Research Ethics Committees. Informed consent for the use of all data collected was obtained from participants following the recommendations of the ALSPAC Ethics and Law Committee at the time. Participants can contact the study team at any time to retrospectively withdraw consent for their data to be used. Study participation is voluntary and during all data collection sweeps, information was provided on the intended use of data. Biological samples are collected in accordance with the Human Tissue Act (2004). Specific Research Ethics Committee approval is sought for the consenting process at each collection sweep. Written consent, including permission for future use, is obtained from adult participants or from the parents of children as appropriate. Ethical approval for future use is covered by ALSPAC's Research Tissue Bank approval. All historical consents to hold biological samples have been reviewed as part of the Tissue Bank approval process. Participants can contact the study team at any time to retrospectively withdraw consent for use of their samples.

### *BiB: Born in Bradford*

Born in Bradford (BiB) is a prospective birth cohort that recruited women with expected delivery between March 2007 and December 2010. Most women were recruited at their oral glucose tolerance test (OGTT) at approximately 26–28 weeks’ gestation, which is offered to all women booked for delivery at Bradford Royal Infirmary, except those with known diabetes. In BiB, most of the obstetric population consists of women of White British or Pakistani origin (together accounting for 81%, with the remaining women being of other ancestries). A total of 12,453 women (13,776 pregnancies) were enrolled during pregnancy who gave birth to 13,858 live children. Full details of the study methodology were reported previously (3). Ethical approval for the study was granted by the Bradford National Health Service Research Ethics Committee (ref 06/Q1202/48).

*HUNT: The Trøndelag Health Study*

The Trøndelag Health Study (HUNT) is a longitudinal, population-based health study conducted in Trøndelag County, Norway. Organizationally, HUNT is a collaboration between HUNT Research Centre (Faculty of Medicine and Health Sciences, Norwegian University of Science and Technology NTNU), Trøndelag County Council, Central Norway Regional Health Authority, and the Norwegian Institute of Public Health. All residents aged 20 or older in the northern part of Trøndelag county were invited to take part in the four HUNT survey waves carried out between 1984 and 2019. All HUNT surveys collected comprehensive information on health, lifestyle, anthropometrics and biological parameters through questionnaires, clinical examination and blood sampling (10). Genotyping was performed for participants in the HUNT2, HUNT3 and HUNT4, as described in detail previously (11). For the present study, we included women who participated in the HUNT2 (1995-97) and/or HUNT3 (2006-08) surveys, had available genetic data and had at least one registered birth in the Medical Birth Registry of Norway in the period from 1967-2021 (N = 25,659). Participation in the HUNT study is voluntary, and all participants provided informed consent. Ethical approval for the present study was granted by the Regional Committee for Medical and Health Research Ethics in Mid-Norway (reference 2018/2488).

### *MoBa: The* Norwegian *Mother, Father and Child Cohort Study*

The Norwegian Mother, Father and Child Cohort Study (MoBa) is a prospective birth cohort that recruited pregnant women from all over Norway from 1999-2008. From all pregnant women, 41% consented to participate. The cohort includes approximately 114,500 children, 95,200 mothers and 75,200 fathers. The establishment of MoBa and initial data collection was based on a license from the Norwegian Data Protection Agency and approval from The Regional Committees for Medical and Health Research Ethics. The MoBa cohort is currently regulated by the Norwegian Health Registry Act. Ethical approval for our study was obtained from The Regional Committees for Medical and Health Research Ethics (ref 2018/1256).

### *UKB: UK Biobank*

All people in the UK National Health Service (NHS) registry aged between 40-69 years and living within approximately 25-mile radius from one of the 22 study centers were invited to participate in UK Biobank (UKB) between 2006-2010 (6, 12). In total, 503,325 adults (5.5% of the ~9.2 million invited) were recruited into the study. Ethical approval for UKB was obtained from the Northwest Multi-Centre Research Ethics Committee (MREC), and our study was performed under UKB application number 23938.

### *FinnGen*

FinnGen is the national wide network of Finnish biobanks. At the time of writing, FinnGen included data from 453,733 individuals (254,618 females and 199,115 males) [12^th^ data release (R12)]. The Coordinating Ethics Committee of the Helsinki and Uusimaa Hospital District has approved the FinnGen consortium (Nr HUS/990/2017), and the ethical approval of each individual study has been described in detail elsewhere (PMID: 32066667). The metadata from FinnGen used by the MR-PREG collaboration is publicly available at <https://www.finngen.fi/en/access_results>.

*Published GWAS metanalyses*

In this study, we used public genetic association data from the International Pregnancy Genetics (InterPregGen) and the Early Growth Genetics (EGG) consortium. InterPregGen is a GWAS meta-analysis for HDP, including 9,515 cases of preeclampsia and 157,719 controls (76% European and 24% Central Asian cases). The EGG consortium includes gestational duration-related traits includes GWAS of preterm birth (N = 18,797 cases and 260,246 controls), postterm birth (N = 15,972 cases and 115,307 controls) and gestational duration (N = 195,555) in European ancestry women.

**Selection of genetic instruments for drug targets**

We selected genetic variants as instruments for each antihypertensive drug target if the met the following criteria:

i) located in the vicinity of the gene encoding the respective target (-/+ 500 kb window around the gene’s transcription start site)
ii) present in the outcome data
iii) Minor allele frequency (MAF) > 1%
iv) Not an ambiguous genetic variant - ie non-palindromic or palindromic variant with MAF < 42%
v) Strongly associated with either systolic (SBP) and/or diastolic blood pressure (DBP) (p < $5\times{10}^{-8}$) based on genetic association data from a GWAS metanalyses of 1,028,980 individuals of European ancestry (13)
vi) In limited LD with other variants in the region (r^2^ < 0.2) using as the reference population a random subsample of 10,000 non-related UK Biobank participants of European ancestry

Genetic association data for SBP and DBP used to select genetic instruments was obtained from a GWAS metanalyses including 1,028,980 individuals of European ancestry from a metanalyses combining UK Biobank, the International Consortium for blood pressure (ICBP), the Million Veteran Program (MVP) and the Vanderbilt University Medical Center's biobank (BioVU) (13). The inclusion of the UKB constitutes a sample overlap between the exposure and some outcome datasets no greater than ~20% relative to the exposure GWAS. Prior to performing the GWAS, individual studies adjusted blood pressure traits by antihypertensive medication intake (i.e. by adding 15 and 10 mmHg to SBP and DBP for individuals on medication, respectively) and by study-specific covariables (e.g. sex, age, body mass index, genotyping array, and principal components of ancestry). Association estimates were expressed as mean change in mmHg per effect allele.

**Genetic instruments strength and validity**

We selected genetic variants to mimic perturbation of eleven drug targets: ACE, ADRA1A, ADRA1B, ADRA1D, ADRB1, ADRB2, AGTR1, CACNA1C, CACNA1D, CACNB2, and SLC12A3. There were no genetic variants that met our selection criteria for ADRA2A, KCNMA1 or NR3C2, meaning we could not include these targets in our analyses. Across the remaining eleven targets, the number of available genetic instruments ranged from 1 (ADRA1D, AGTR1, CACNA1C, SLC12A3) to 25 (CACNB2). The amount of variance in blood pressure explained by individual SNPs ranged from 0.002% to 0.03% with corresponding F statistics ranging from 30 to 408 (**Supplementary tables 4 and 5**).

For several drug targets, the blood pressure-lowering allele of the top genetic variant was associated with decreased expression of the target gene in one or more tissues—ACE (adrenal gland, arterial, and heart tissues), ADRA1B (kidney), ADRB1 (arterial tissues), and CACNA1C and CACNB2 (aorta). In contrast, the same allele was associated with increased expression of ACE (kidney and lung), ADRB2 (tibial artery), AGTR1 (lung), and SLC12A3 (lung) (Supplementary figure 1), which might reflect tissue- or cell-specificities in gene expression regulation, or cross-tissue feedback mechanisms. Steiger directionality tests indicated that the top variants explained more variance in gene expression than in blood pressure (Supplementary table 6), and colocalization analyses supported a shared causal variant, with a few exceptions for which the evidence was inconclusive [(ACE (heart), ADRA1B (kidney), CACNB2 (aorta), SLC12A3 (lung)] and one case consistent with distinct causal variants [ACE (lung)] (Supplementary table 7). For other targets—ADRA1A and CACNA1D—no expression changes were detected, whereas for ADRA1D, no suitable genetic instrument or proxy was available in the expression dataset (Supplementary figure 1).

The phenome-wide scan indicated that the selected genetic variants were largely associated with changes in blood pressure and related phenotypes (e.g. antihypertensive medication and cardiovascular disease) in the expected direction. Some genetic variants exhibited associations with other phenotypes, including blood cell traits (ADRA1D, ADRB2, CACNA1C, CACNB2 variant), lipid traits (ADRB1, SLC12A3 variant), height (ADRA1D variant), age at menarche (CACNA1C variant), lung function (ADRA1B, CACNA1D variant), birthweight (ADRB1, AGTR1 variant), schizophrenia (CACNA1C variant), and Alzheimer’s disease (ACE variant), with these being weaker than those for blood pressure and associated traits (Supplementary figure 2 and Supplementary table 8). In positive control analyses, genetically-instrumented downregulation of blood pressure via the various drug targets was related to lower risk of stroke (ACE, CACNA1D, SLC12A3), coronary artery disease (ADRA1A, ADRB1) or both (ADRB2, CACNB2), but higher risk of stroke for ADRA1B (Supplementary table 9).

**STROBE-MR checklist of recommended items to address in reports of Mendelian randomization studies**^1^ ^2^

| **Item No.** | **Section** | **Checklist item** | **Page No.** | **Relevant text from manuscript** |
| --- | --- | --- | --- | --- |
| 1 | **TITLE and ABSTRACT** | Indicate Mendelian randomization (MR) as the study’s design in the title and/or the abstract if that is a main purpose of the study | Abstract | Abstract: “We performed a Mendelian randomization study...” |
|  | **INTRODUCTION** |  |  |  |
| 2 | **Background** | Explain the scientific background and rationale for the reported study. What is the exposure? Is a potential causal relationship between exposure and outcome plausible? Justify why MR is a helpful method to address the study question | 6-7 | e.g.:  “Hypertension during pregnancy is associated with a three to five-fold higher risk of pre-eclampsia, preterm birth, small-for-gestational age (SGA) babies and perinatal death, as well as a five to ten-fold higher risk of maternal death. Antihypertensive treatment is usually indicated for pregnant women with hypertension. …”  “There is very limited evidence on the comparative benefits and adverse effects for different antihypertensive drug classes for pregnancy-related outcomes…”  “Drug target Mendelian randomization harnesses the properties of human genetic variants affecting the expression or function of proteins, which constitute most drug targets, to interrogate the effect of pharmacologically perturbing these drug targets…” |
| 3 | **Objectives** | State specific objectives clearly, including pre-specified causal hypotheses (if any). State that MR is a method that, under specific assumptions, intends to estimate causal effects | 8 | “This study aimed to improve evidence on (i) the potential effects of antihypertensive drug treatment on a range of maternal and fetal outcomes, and (ii) the comparative benefits and risk profile of different antihypertensive drugs in pregnancy. To do that, we used drug target Mendelian randomization to mimic pharmacological perturbation of targets from eight commonly used types of antihypertensive drugs.” |
|  | **METHODS** |  |  |  |
| 4 | **Study design and data sources** | Present key elements of the study design early in the article. Consider including a table listing sources of data for all phases of the study. For each data source contributing to the analysis, describe the following: |  |  |
|  | a) | Setting: Describe the study design and the underlying population, if possible. Describe the setting, locations, and relevant dates, including periods of recruitment, exposure, follow-up, and data collection, when available. | Suppl text (p 1-4) |  |
|  | b) | Participants: Give the eligibility criteria, and the sources and methods of selection of participants. Report the sample size, and whether any power or sample size calculations were carried out prior to the main analysis | Suppl text (p 1-2)  Table 2 | No power calculation was carried out |
|  | c) | Describe measurement, quality control and selection of genetic variants | 12  +  Suppl text (p 3) | Details on study-specific measurement and quality control of genetic variants have been previously published (14) |
|  | d) | For each exposure, outcome, and other relevant variables, describe methods of assessment and diagnostic criteria for diseases | Suppl text (p 3)  Supplementary table 3 | Details on study-specific diagnostic criteria for each outcome of interest have been previously published (14) |
|  | e) | Provide details of ethics committee approval and participant informed consent, if relevant | Suppl text (p 1-2) | “Ethical approval for the study was obtained from the ALSPAC Ethics and Law Committee and the Local Research Ethics Committees.”  “Ethical approval for the study was granted by the Bradford National Health Service Research Ethics Committee (ref 06/Q1202/48).”  “Ethical approval for the present study was granted by the Regional Committee for Medical and Health Research Ethics in Mid-Norway (reference 2018/2488). “  “Ethical approval for our study was obtained from The Regional Committees for Medical and Health Research Ethics (ref 2018/1256)”  “Ethical approval for UKB was obtained from the Northwest Multi-Centre Research Ethics Committee (MREC), …”  “The Coordinating Ethics Committee of the Helsinki and Uusimaa Hospital District has approved the FinnGen consortium (Nr HUS/990/2017), and the ethical approval of each individual study has been described in detail elsewhere (PMID: 32066667). |
| 5 | **Assumptions** | Explicitly state the three core IV assumptions for the main analysis (relevance, independence and exclusion restriction) as well assumptions for any additional or sensitivity analysis | 16 |  |
| 6 | **Statistical methods: main analysis** | Describe statistical methods and statistics used |  |  |
|  | a) | Describe how quantitative variables were handled in the analyses (i.e., scale, units, model) | Suppl text (p 4) | “…Association estimates were expressed as mean change in mmHg per effect allele” |
|  | b) | Describe how genetic variants were handled in the analyses and, if applicable, how their weights were selected | 12  +  Suppl text (p 3) | “In Supplementary methods, we give a full description of our procedure to select genetic instruments. In brief, our main criteria were based on:   - location in the vicinity of the drug target encoding gene (aka cis-acting variants) – i.e. within a window of -/+ 500kb around the transcription start site of the target encoding gene - potential impact on a downstream effect of the target – i.e. variants with strong evidence of association with mean changes in systolic (SBP) and/or diastolic blood pressure (DBP) at p < 5×10^(-8) in a GWAS metanalysis including 1,028,980 individuals of European ancestry (44) - limited correlation (i.e. linkage disequilibrium [LD]) with other variants selected from the same window (r2 < 0.2 in the main analysis and r2<0.01 as sensitivity analysis)” |
|  | c) | Describe the MR estimator (e.g. two-stage least squares, Wald ratio) and related statistics. Detail the included covariates and, in case of two-sample MR, whether the same covariate set was used for adjustment in the two samples | 15 | “For the main analyses, we used the Wald ratio or the generalised inverse variance weighted (gIVW) estimators if, respectively, one or multiple genetic variants were selected for a given target. The gIVW is an extension of the standard IVW method that accounts for LD between genetic variants (47). We used a random subsample of 10,000 unrelated UK Biobank participants of European ancestry to estimate the extent of LD between variants, indicated by the R2, as required by gIVW.” |
|  | d) | Explain how missing data were addressed | NA |  |
|  | e) | If applicable, indicate how multiple testing was addressed | 15 | " We selected results showing some evidence of association in the main analyses (p < 0.05) for further evaluation in sensitivity analyses, and also report results corrected for multiple testing using a 5% false discovery rate (FDR) control. However, our interpretation focuses on the direction and precision of effect estimates from the main analyses, as well as their consistency across the sensitivity analyses described below, to provide a more comprehensive assessment of the evidence.” |
| 7 | **Assessment of assumptions** | Describe any methods or prior knowledge used to assess the assumptions or justify their validity | 13-17 | Please refer to methods sections “Characterising genetic instruments” and “Sensitivity analyses” |
| 8 | **Sensitivity analyses and additional analyses** | Describe any sensitivity analyses or additional analyses performed (e.g. comparison of effect estimates from different approaches, independent replication, bias analytic techniques, validation of instruments, simulations) | 15-17 | Please refer to methods section “Sensitivity analyses” |
| 9 | **Software and pre-registration** |  |  |  |
|  | a) | Name statistical software and package(s), including version and settings used | 11, 14, 15, 17 | “All analyses were performed using R version 4.3.1.”  “using the ieugwasr R package version (48)(47) (access: 22nd July 2025)”  “These were undertaken using the TwoSampleMR and MendelianRandomization R packages”  “using the coloc R package”  “using a weighted linear model (WLM) (53) implemented in the DONUTS R package” |
|  | b) | State whether the study protocol and details were pre-registered (as well as when and where) | 11 | “The study protocol was not pre-registered.” |
|  | **RESULTS** |  |  |  |
| 10 | **Descriptive data** |  |  |  |
|  | a) | Report the numbers of individuals at each stage of included studies and reasons for exclusion. Consider use of a flow diagram | NA | Details on study-specific exclusions have been previously published (14) |
|  | b) | Report summary statistics for phenotypic exposure(s), outcome(s), and other relevant variables (e.g. means, SDs, proportions) | Table 2 | Details on study-specific phenotype distributions have been previously published (14) |
|  | c) | If the data sources include meta-analyses of previous studies, provide the assessments of heterogeneity across these studies | 15 | “We conducted leave-one-study-out analysis to explore potential bias due to outlying studies. First, we performed Mendelian randomization for each study separately using the same methods as in the main analyses. Second, we combined study-specific estimates using fixed effect metanalyses with inverse variance weights after removing one study at a time.” |
|  | d) | For two-sample MR:  i.  Provide justification of the similarity of the genetic variant-exposure associations between the exposure and outcome samples  ii.  Provide information on the number of individuals who overlap between the exposure and outcome studies | Suppl text (p4) | “Genetic association data for SBP and DBP used to select genetic instruments was obtained from a GWAS metanalyses including 1,028,980 individuals of European ancestry…”  Details on the ancestral background of the outcome sample have been previously published (14)  “The inclusion of the UKB constitutes a sample overlap between the exposure and some outcome datasets no greater than ~20% relative to the exposure GWAS.” |
| 11 | **Main results** |  |  |  |
|  | a) | Report the associations between genetic variant and exposure, and between genetic variant and outcome, preferably on an interpretable scale | ST5 |  |
|  | b) | Report MR estimates of the relationship between exposure and outcome, and the measures of uncertainty from the MR analysis, on an interpretable scale, such as odds ratio or relative risk per SD difference | 19-20,  Figures 2, S3  ST9 | Most estimates expressed as OR of outcome per mmHg decrease in BP |
|  | c) | If relevant, consider translating estimates of relative risk into absolute risk for a meaningful time period |  | Not applied |
|  | d) | Consider plots to visualize results (e.g. forest plot, scatterplot of associations between genetic variants and outcome versus between genetic variants and exposure) | Figures 2,3, S3,S4, S5 | Forest plots were used to present and compare MR estimates |
| 12 | **Assessment of assumptions** |  |  |  |
|  | a) | Report the assessment of the validity of the assumptions | 23-25 | Please refer to section “Sensitivity analyses” |
|  | b) | Report any additional statistics (e.g., assessments of heterogeneity across genetic variants, such as *I^2^*, Q statistic or E-value) | ST5, ST8, ST9 | Heterogeneity statistics are reported |
| 13 | **Sensitivity analyses and additional analyses** |  |  |  |
|  | a) | Report any sensitivity analyses to assess the robustness of the main results to violations of the assumptions | 25-27 | Please refer to results sections “Sensitivity analyses” |
|  | b) | Report results from other sensitivity analyses or additional analyses | 25-27 | Please refer to results sections “Sensitivity analyses” |
|  | c) | Report any assessment of direction of causal relationship (e.g., bidirectional MR) | 18, ST5 | “Steiger directionality tests indicated that the top variants explained more variance in gene expression than in blood pressure (Supplementary table 5),…” |
|  | d) | When relevant, report and compare with estimates from non-MR analyses |  | NA |
|  | e) | Consider additional plots to visualize results (e.g., leave-one-out analyses) | Suppl figures | Forest plots were used to show results |
|  | **DISCUSSION** |  |  |  |
| 14 | **Key results** | Summarize key results with reference to study objectives | 32 | Please refer to the 1^st^ paragraph of “Discussion” |
| 15 | **Limitations** | Discuss limitations of the study, taking into account the validity of the IV assumptions, other sources of potential bias, and imprecision. Discuss both direction and magnitude of any potential bias and any efforts to address them | 34-35 | Please refer to the 5^th^ paragraph of “Discussion” |
| 16 | **Interpretation** |  |  |  |
|  | a) | Meaning: Give a cautious overall interpretation of results in the context of their limitations and in comparison with other studies | 35 | Please refer to the last paragraph of “Discussion” |
|  | b) | Mechanism: Discuss underlying biological mechanisms that could drive a potential causal relationship between the investigated exposure and the outcome, and whether the gene-environment equivalence assumption is reasonable. Use causal language carefully, clarifying that IV estimates may provide causal effects only under certain assumptions | 32-34 | Please refer to the 3^rd^ to 4^th^ paragraphs of “Discussion” |
|  | c) | Clinical relevance: Discuss whether the results have clinical or public policy relevance, and to what extent they inform effect sizes of possible interventions | 33-34 | Please refer to the 3^rd^, 4^th^ and last paragraph of “Discussion” |
| 17 | **Generalizability** | Discuss the generalizability of the study results (a) to other populations, (b) across other exposure periods/timings, and (c) across other levels of exposure | 33 | “We utilized large-scale population data, increasing statistical power to assess a broader range of clinically important outcomes compared to previous RCTs and enhancing the generalizability of our findings.” |
|  | **OTHER INFORMATION** |  |  |  |
| 18 | **Funding** | Describe sources of funding and the role of funders in the present study and, if applicable, sources of funding for the databases and original study or studies on which the present study is based | 36 | Please refer to the “Funding” statement. |
| 19 | **Data and data sharing** | Provide the data used to perform all analyses or report where and how the data can be accessed, and reference these sources in the article. Provide the statistical code needed to reproduce the results in the article, or report whether the code is publicly accessible and if so, where | 36 | Please refer to “Data Availability Statement”. |
| 20 | **Conflicts of Interest** | All authors should declare all potential conflicts of interest | 35 | Please refer to “Disclosure Of Interest” |

This checklist is copyrighted by the Equator Network under the Creative Commons Attribution 3.0 Unported (CC BY 3.0) license.

1.	Skrivankova VW, Richmond RC, Woolf BAR, Yarmolinsky J, Davies NM, Swanson SA, et al. Strengthening the Reporting of Observational Studies in Epidemiology using Mendelian Randomization (STROBE-MR) Statement. JAMA. 2021;under review.

2.	Skrivankova VW, Richmond RC, Woolf BAR, Davies NM, Swanson SA, VanderWeele TJ, et al. Strengthening the Reporting of Observational Studies in Epidemiology using Mendelian Randomisation (STROBE-MR): Explanation and Elaboration. BMJ. 2021;375:n2233.

**References**

1.	Boyd A, Golding J, Macleod J, Lawlor DA, Fraser A, Henderson J, et al. Cohort Profile: the 'children of the 90s'--the index offspring of the Avon Longitudinal Study of Parents and Children. Int J Epidemiol. 2013;42(1):111-27.

2.	Fraser A, Macdonald-Wallis C, Tilling K, Boyd A, Golding J, Davey Smith G, et al. Cohort Profile: the Avon Longitudinal Study of Parents and Children: ALSPAC mothers cohort. Int J Epidemiol. 2013;42(1):97-110.

3.	Wright J, Small N, Raynor P, Tuffnell D, Bhopal R, Cameron N, et al. Cohort Profile: the Born in Bradford multi-ethnic family cohort study. Int J Epidemiol. 2013;42(4):978-91.

4.	Magnus P, Irgens LM, Haug K, Nystad W, Skjaerven R, Stoltenberg C, et al. Cohort profile: the Norwegian Mother and Child Cohort Study (MoBa). Int J Epidemiol. 2006;35(5):1146-50.

5.	Magnus P, Birke C, Vejrup K, Haugan A, Alsaker E, Daltveit AK, et al. Cohort Profile Update: The Norwegian Mother and Child Cohort Study (MoBa). Int J Epidemiol. 2016;45(2):382-8.

6.	Sudlow C, Gallacher J, Allen N, Beral V, Burton P, Danesh J, et al. UK biobank: an open access resource for identifying the causes of a wide range of complex diseases of middle and old age. PLoS Med. 2015;12(3):e1001779.

7.	Kurki MI, Karjalainen J, Palta P, Sipila TP, Kristiansson K, Donner KM, et al. FinnGen provides genetic insights from a well-phenotyped isolated population. Nature. 2023;613(7944):508-18.

8.	Morgan L, McGinnis R, Steinthorsdottir V, Svyatova G, Zakhidova N, Lee WK, et al. InterPregGen: genetic studies of pre-eclampsia in three continents. Nor Epidemiol. 2014;24(1-2):141-6.

9.	Sole-Navais P, Flatley C, Steinthorsdottir V, Vaudel M, Juodakis J, Chen J, et al. Genetic effects on the timing of parturition and links to fetal birth weight. Nat Genet. 2023;55(4):559-67.

10.	Krokstad S, Langhammer A, Hveem K, Holmen TL, Midthjell K, Stene TR, et al. Cohort Profile: the HUNT Study, Norway. Int J Epidemiol. 2013;42(4):968-77.

11.	Brumpton BM, Graham S, Surakka I, Skogholt AH, Loset M, Fritsche LG, et al. The HUNT study: A population-based cohort for genetic research. Cell Genom. 2022;2(10):100193.

12.	Littlejohns TJ, Sudlow C, Allen NE, Collins R. UK Biobank: opportunities for cardiovascular research. Eur Heart J. 2019;40(14):1158-66.

13.	Keaton JM, Kamali Z, Xie T, Vaez A, Williams A, Goleva SB, et al. Genome-wide analysis in over 1 million individuals of European ancestry yields improved polygenic risk scores for blood pressure traits. Nat Genet. 2024;56(5):778-91.

14.	McBride N, Clayton GL, Goncalves Soares A, Yang Q, Bond TA, Taylor A, et al. Cohort profile: the Mendelian randomisation in pregnancy (MR-PREG) collaboration - improving evidence for prevention and treatment of adverse pregnancy and perinatal outcomes. BMJ Open. 2026;16(3):e103753.
