## Supplementary figures for "Using human genetics to understand the effect of modulating targets of antihypertensive drugs in pregnancy"

**Supplementary figure 1**. Change in drug target gene expression per blood pressure decreasing allele

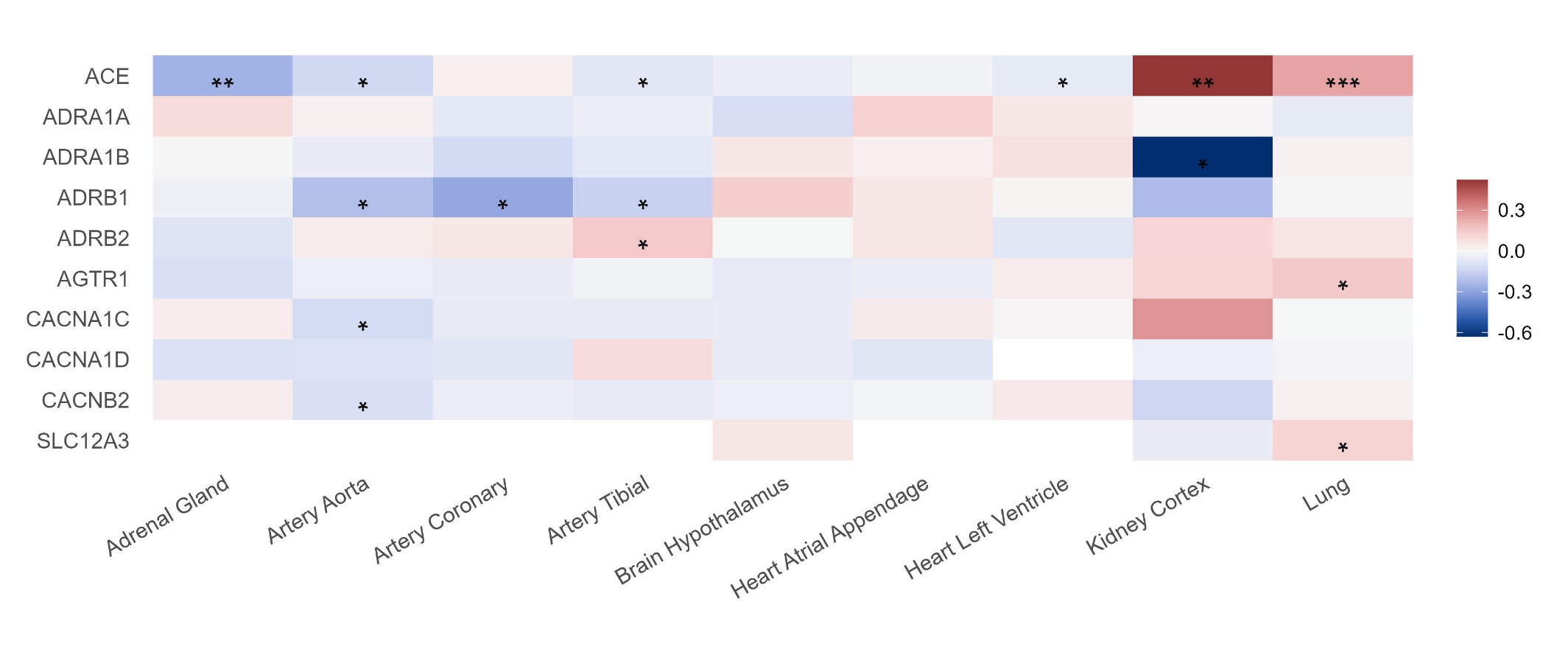

Results are expressed as standardised unit change in gene expression across tissues per blood pressure **decreasing** allele of the top genetic variant selected for each drug target. Blue, red, and grey boxes denote, respectively, decreases, increases, and no change in gene expression, while white boxes represent missing data. Asterisks indicate P value: < $5\times{10}^{-8}$ (***), < $5\times{10}^{-5}$ (**), and < $5\times{10}^{-2}$ (*). Please note that results for *ADRA1D* (ENSG00000171873) are not provided as the selected genetic variant was not present in GTEx v8 data and no suitable proxies (R^2^ > 0.8 in 1000 Genomes Europeans) could be identified. *ACE*: angiotensin-converting enzyme; *ADRA1*: alpha-1A adrenergic receptor; *ADRAB*: alpha-1B adrenergic receptor; *ADRB1*: beta-1 adrenergic receptor; *ADRB2*: beta-2 adrenergic receptor; *AGTR1*: Type-1 angiotensin II receptor; *CACNA1C*: Voltage-dependent L-type calcium channel subunit alpha-1C; *CACNA1D*: Voltage-dependent L-type calcium channel subunit alpha-1D; *CACNB2*: Voltage-dependent L-type calcium channel subunit beta-2; *SLC12A3*: Solute carrier family 12 member 3.

**Supplementary figure 2**. Phenome-wide scan of the top genetic variant in each drug target region

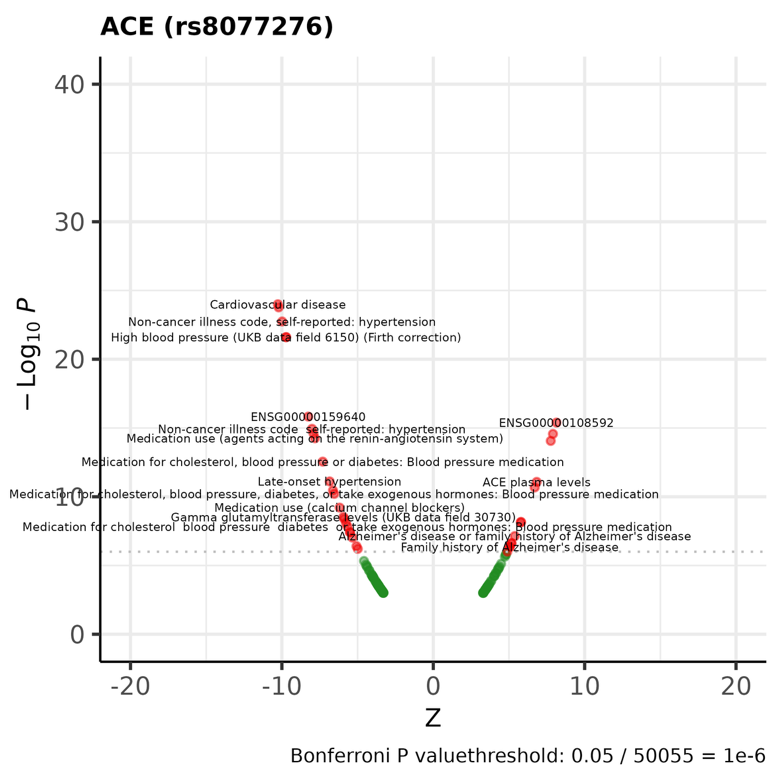

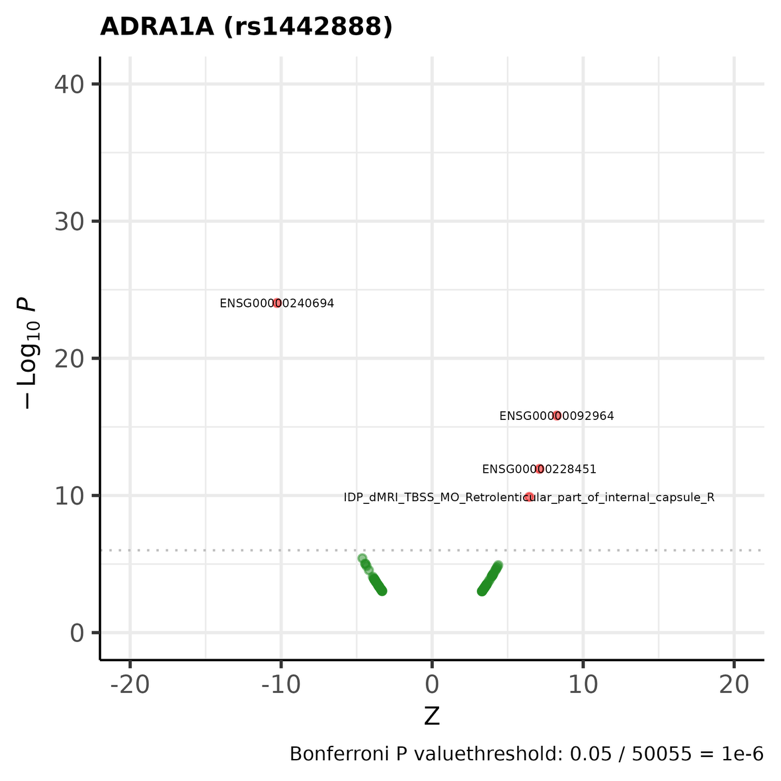

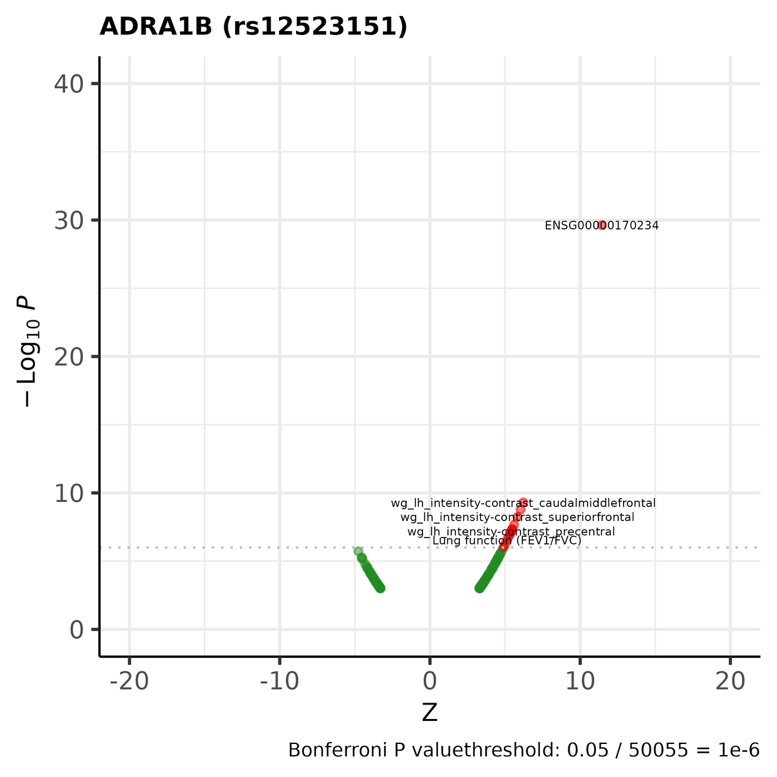

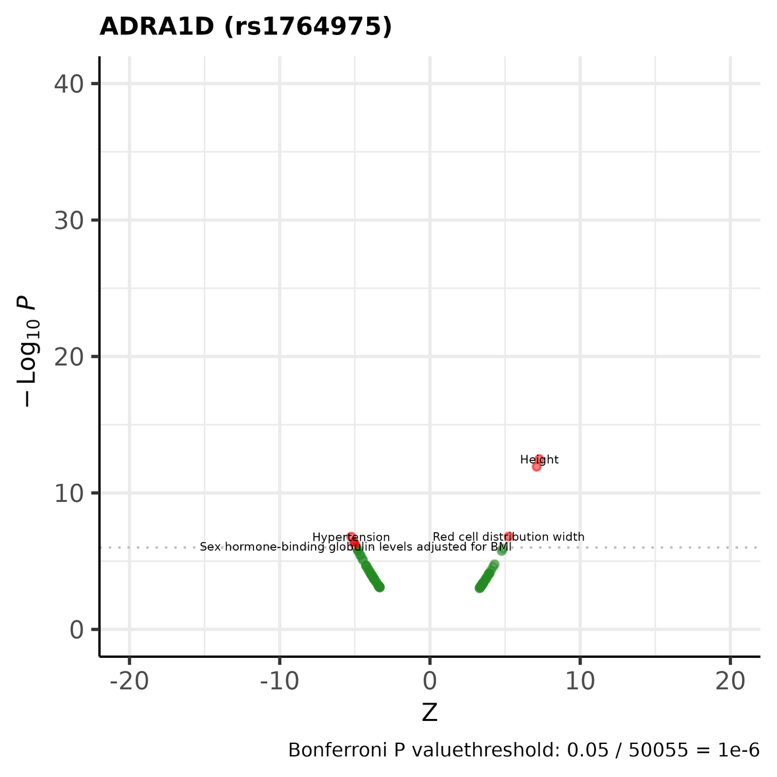

Results are expressed in log10 p-values (y axis) and z statistics (x axis). The SNP effect allele corresponds to the blood pressure decreasing allele. Results passing Bonferroni correction are highlighted in red (total number of traits: 50,055).

**Supplementary figure 2 (cont.)**. Phenome-wide scan of the top genetic variant in each drug target region

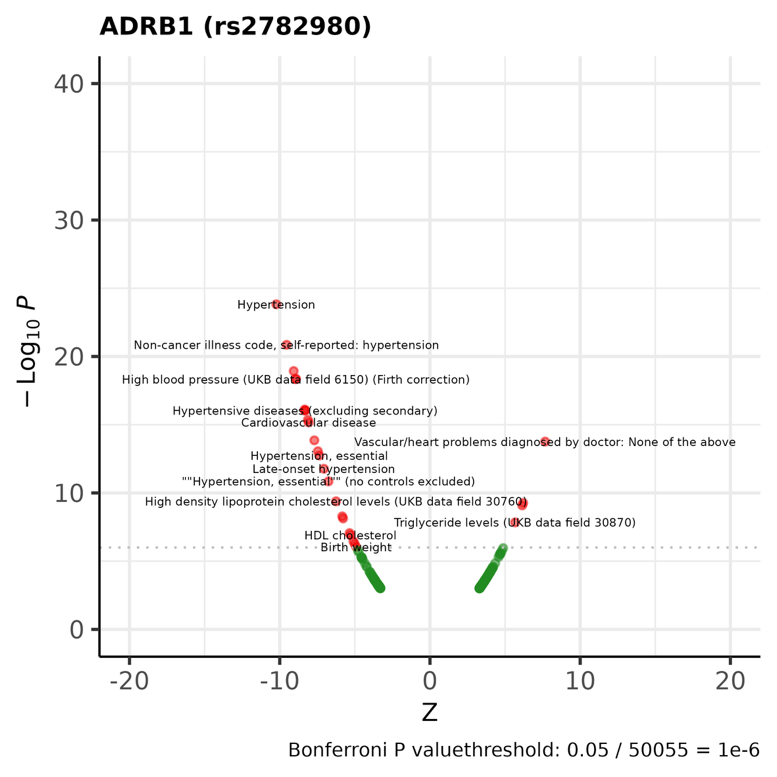

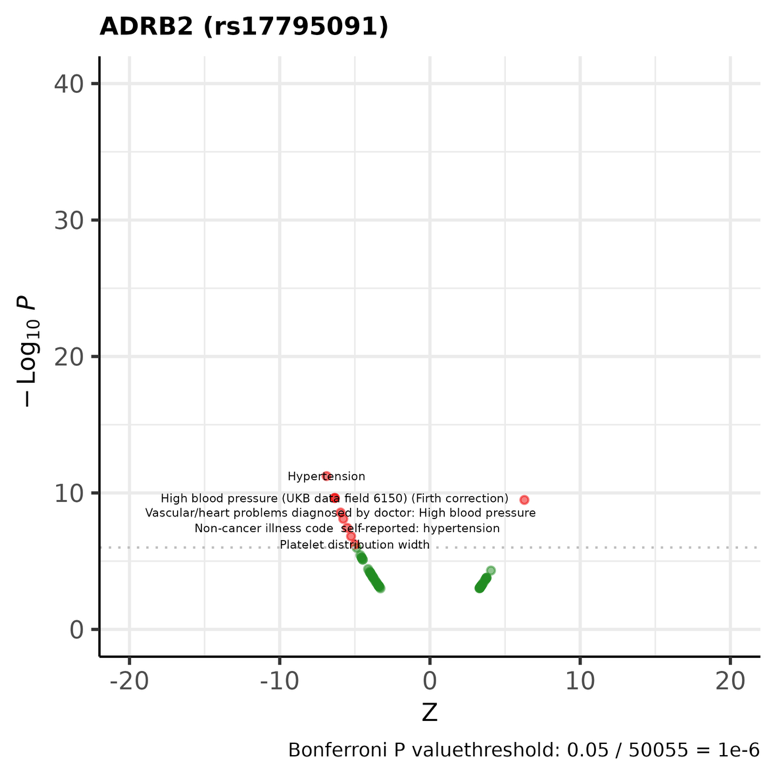

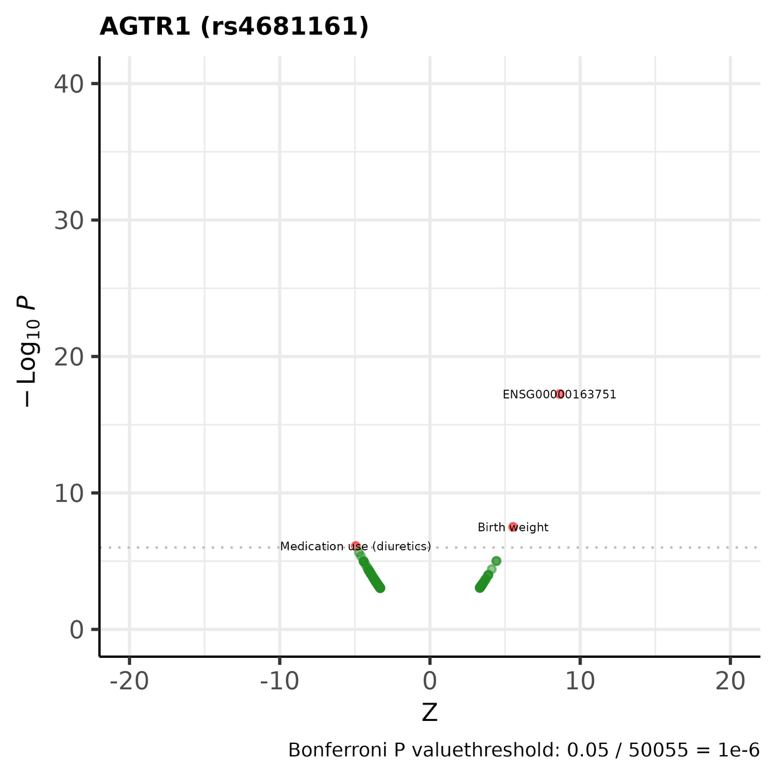

Results are expressed in log10 p-values (y axis) and z statistics (x axis). The SNP effect allele corresponds to the blood pressure decreasing allele. Results passing Bonferroni correction are highlighted in red (total number of traits: 50,055).

**Supplementary figure 2 (cont.)**. Phenome-wide scan of the top genetic variant in each drug target region

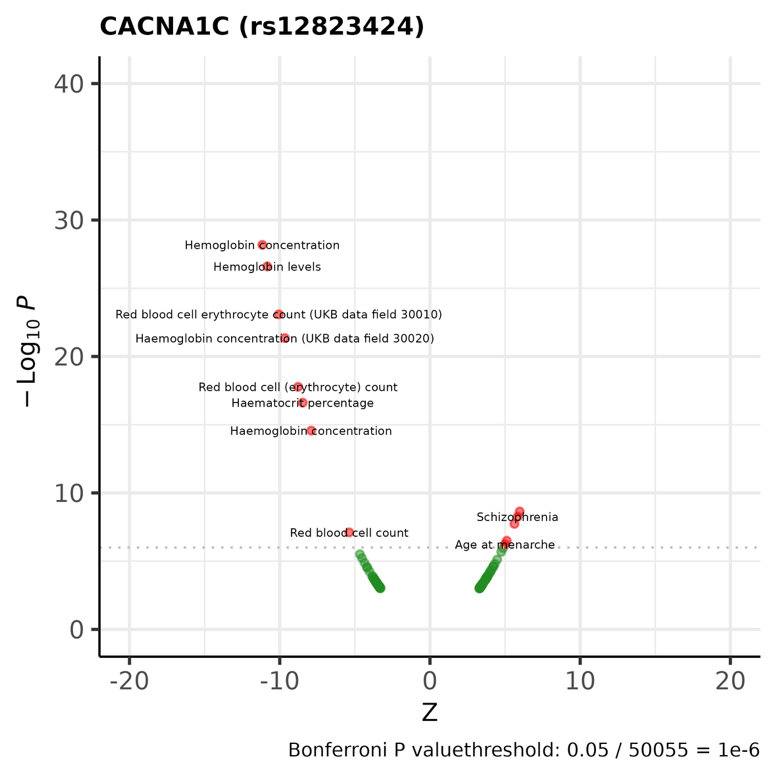

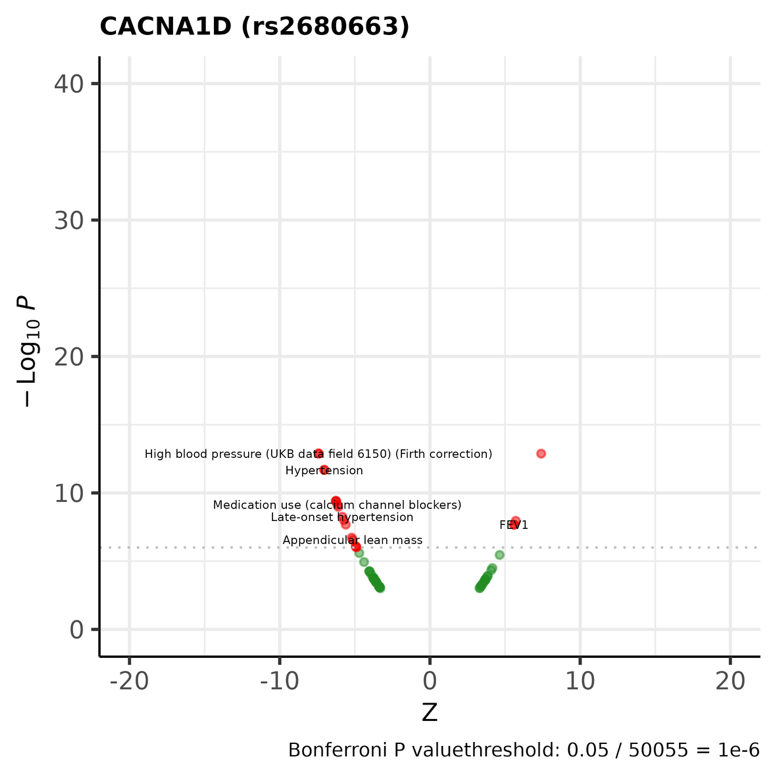

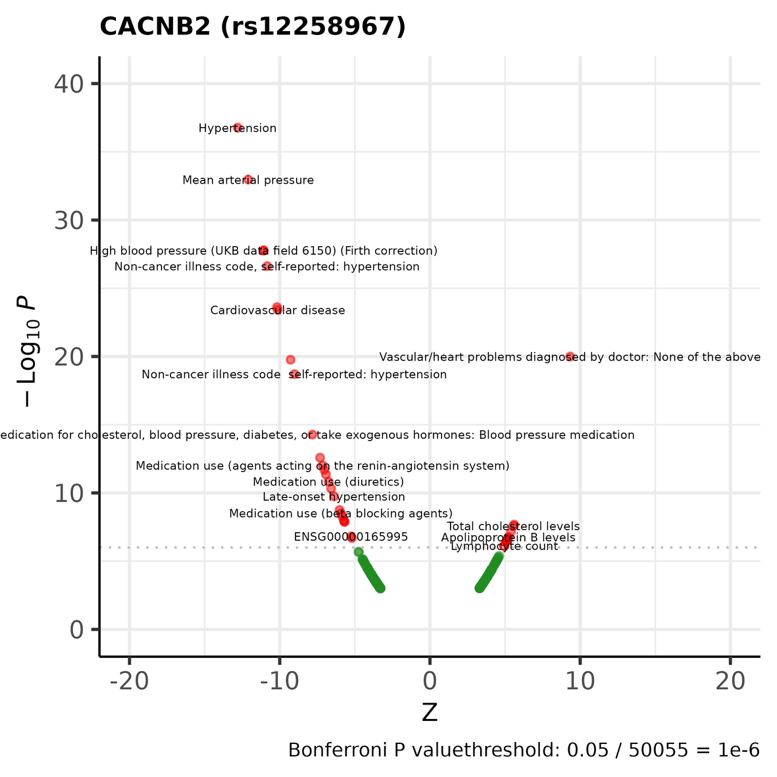

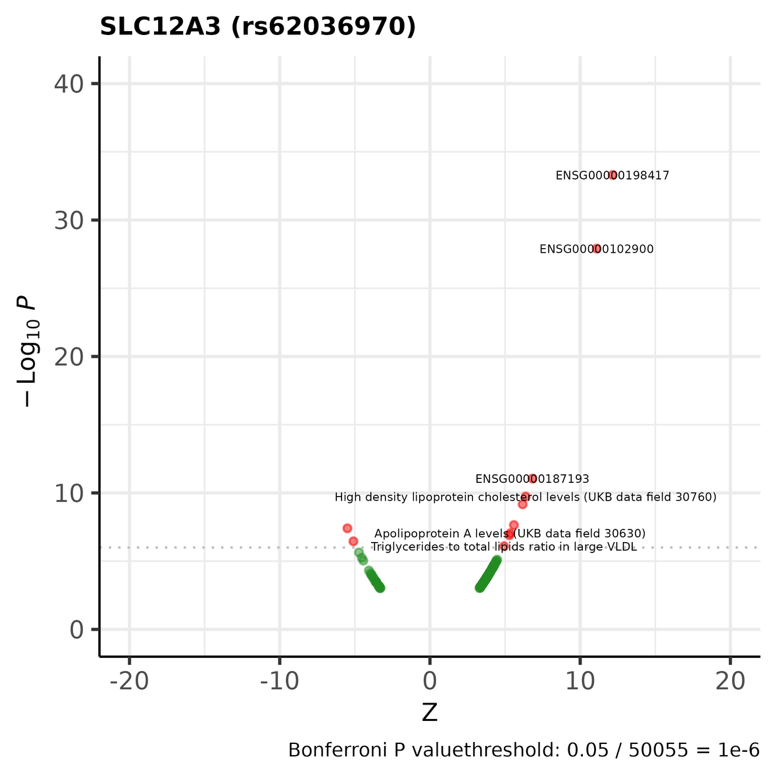

Results are expressed in log10 p-values (y axis) and z statistics (x axis). The SNP effect allele corresponds to the blood pressure decreasing allele. Results passing Bonferroni correction are highlighted in red (total number of traits: 50,055).

**Supplementary figure 3.** Mendelian randomization estimates for the effect of modulating hypertension drug targets on secondary continuous pregnancy-related outcomes

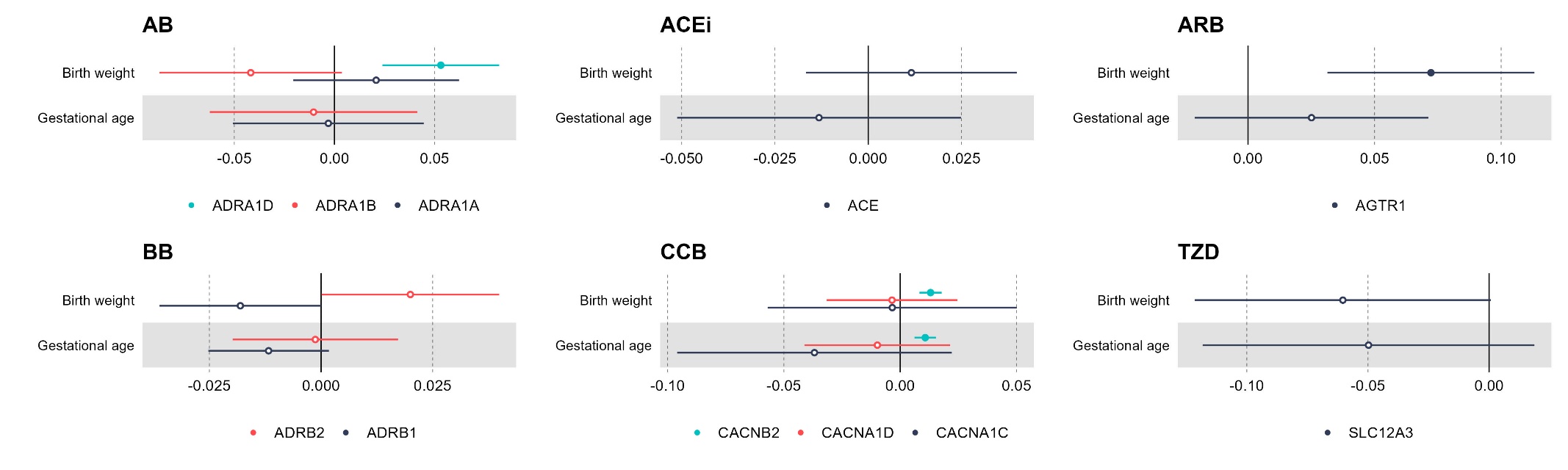
Results are expressed as mean difference in standard deviation (SD) units of outcome per 1 mmHg decrease in systolic or diastolic blood pressure (BP) due to genetically-instrumented downregulation of the drug target gene. Point estimates and 95% confidence intervals are represented by circles and bars, respectively. Filled circles indicate estimates with false discovery rate-corrected *p*values < 0.05.

ACE: Angiotensin-converting enzyme; ADRA1A: adrenergic receptor α1A; ADRA1B: adrenergic receptor α1B; ADRA1D: adrenergic receptor α1D; ADRB1: adrenergic receptor β1; ADRB2: adrenergic receptor β2; AGTR1: angiotensin II receptor type 1; CACNA1C: Voltage-dependent L-type calcium channel subunit α1C; CACNA1D: Voltage-dependent L-type calcium channel subunit α1D; CACNB2: Voltage-dependent L-type calcium channel subunit β2; SLC12A3: Solute carrier family 12 member 3.

**Supplementary figure 4A**. MR estimates for the effect of modulating hypertension drug targets on primary pregnancy-related outcomes removing one study at a time (leave-one-study-out analyses)

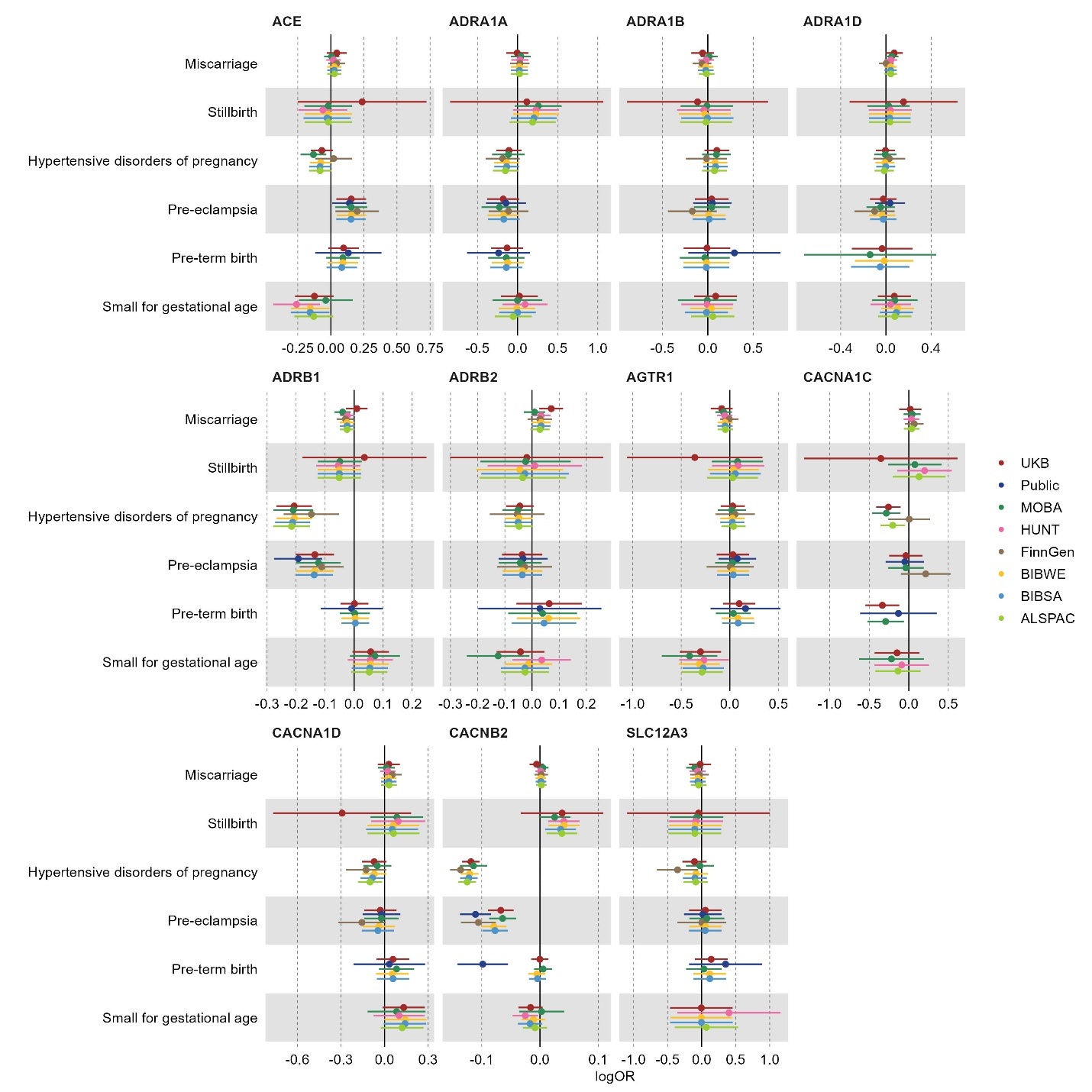

Results are expressed as log odds ratio (OR) of outcome per 1 mmHg unit decrease in systolic or diastolic blood pressure (BP) due to genetically-proxied downregulation of the drug target gene. Point estimates and 95% confidence intervals are represented by circles and bars, respectively.

ACE: Angiotensin-converting enzyme; ADRA1A: adrenergic receptor α1A; ADRA1B: adrenergic receptor α1B; ADRA1D: adrenergic receptor α1D; ADRB1: adrenergic receptor β1; ADRB2: adrenergic receptor β2; AGTR1: angiotensin II receptor type 1; CACNA1C: Voltage-dependent L-type calcium channel subunit α1C; CACNA1D: Voltage-dependent L-type calcium channel subunit α1D; CACNB2: Voltage-dependent L-type calcium channel subunit β2; SLC12A3: Solute carrier family 12 member 3.

**Supplementary figure 4B**. MR estimates for the effect of modulating hypertension drug targets on secondary (binary) pregnancy-related outcomes removing one study at a time (leave-one-study-out analyses)

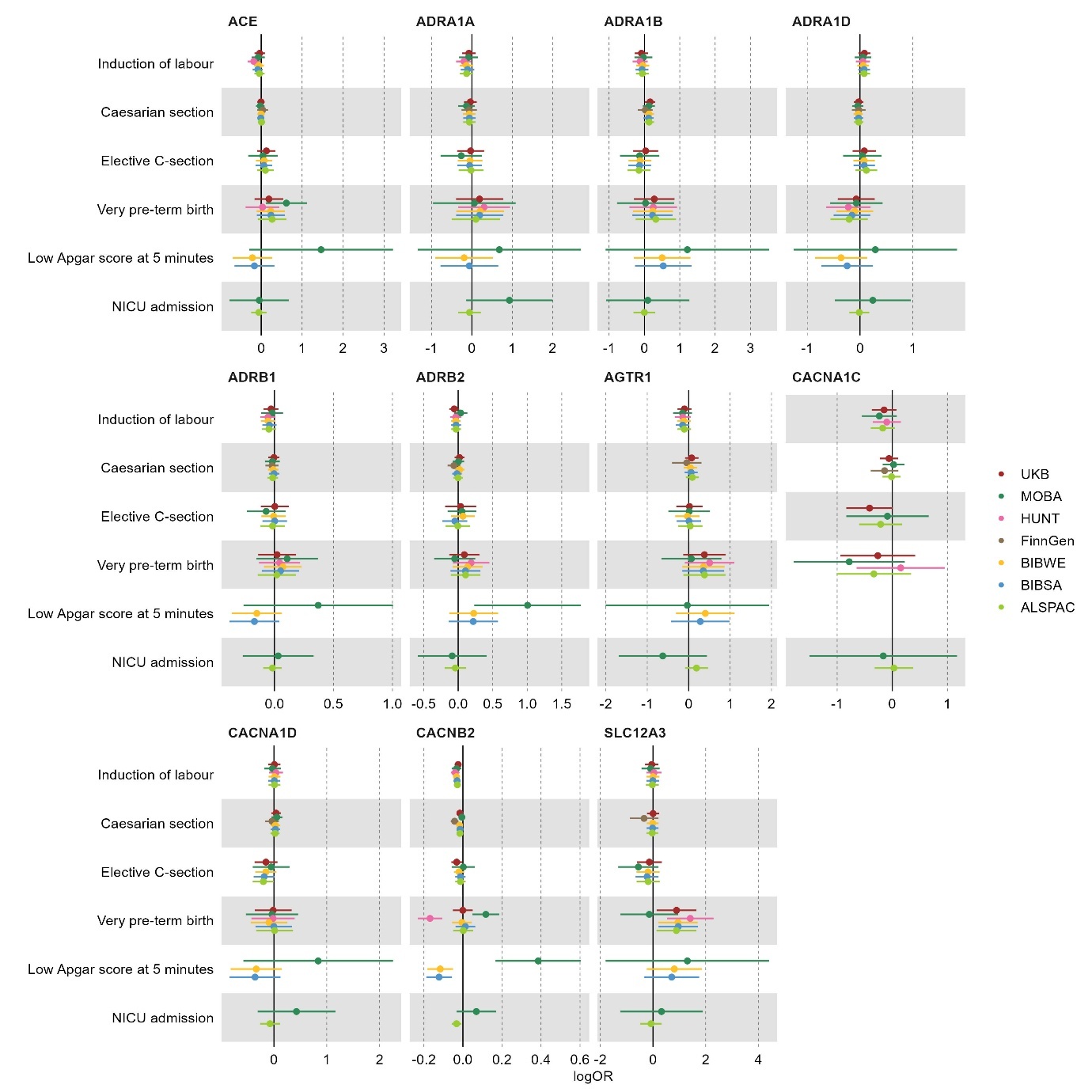

Results are expressed as log odds ratio (OR) of outcome per 1 mmHg unit decrease in systolic or diastolic blood pressure (BP) due to genetically-proxied downregulation of the drug target gene. Point estimates and 95% confidence intervals are represented by circles and bars, respectively.

ACE: Angiotensin-converting enzyme; ADRA1A: adrenergic receptor α1A; ADRA1B: adrenergic receptor α1B; ADRA1D: adrenergic receptor α1D; ADRB1: adrenergic receptor β1; ADRB2: adrenergic receptor β2; AGTR1: angiotensin II receptor type 1; CACNA1C: Voltage-dependent L-type calcium channel subunit α1C; CACNA1D: Voltage-dependent L-type calcium channel subunit α1D; CACNB2: Voltage-dependent L-type calcium channel subunit β2; SLC12A3: Solute carrier family 12 member 3.

**Supplementary figure 4C**. MR estimates for the effect of modulating hypertension drug targets on secondary (continuous) pregnancy-related outcomes removing one study at a time (leave-one-study-out analyses)

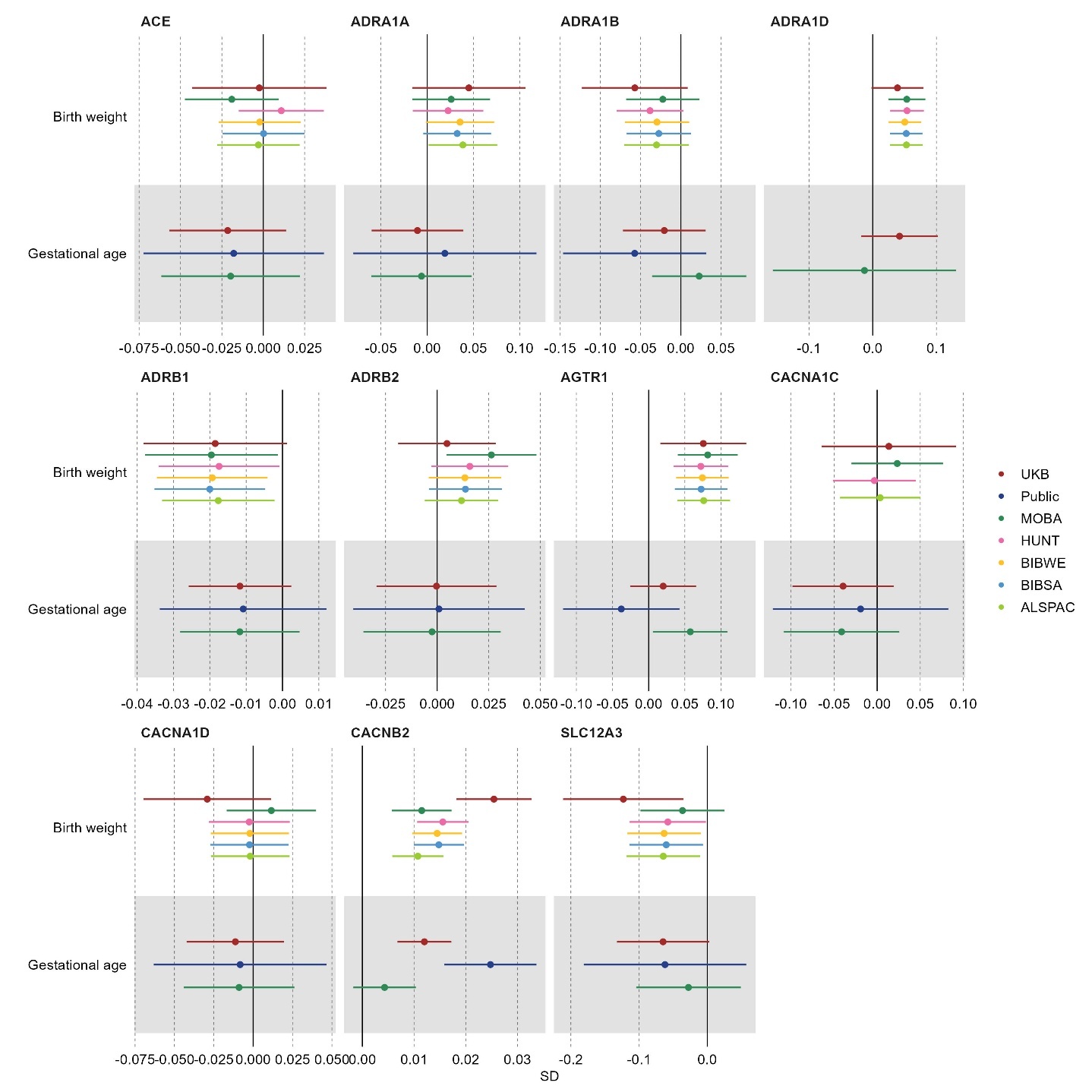

Results are expressed as mean difference in standard deviation (SD) units of outcome per 1 mmHg unit decrease in systolic or diastolic blood pressure (BP) due to genetically-proxied downregulation of the drug target gene. Point estimates and 95% confidence intervals are represented by circles and bars, respectively.

ACE: Angiotensin-converting enzyme; ADRA1A: adrenergic receptor α1A; ADRA1B: adrenergic receptor α1B; ADRA1D: adrenergic receptor α1D; ADRB1: adrenergic receptor β1; ADRB2: adrenergic receptor β2; AGTR1: angiotensin II receptor type 1; CACNA1C: Voltage-dependent L-type calcium channel subunit α1C; CACNA1D: Voltage-dependent L-type calcium channel subunit α1D; CACNB2: Voltage-dependent L-type calcium channel subunit β2; SLC12A3: Solute carrier family 12 member 3.

**Supplementary figure 5A**. MR estimates for the effect of modulating hypertension drug targets on primary pregnancy-related outcomes using different linkage disequilibrium thresholds for selecting genetic variants (R^2^ < 0.20 in the main analyses and R^2^ < 0.01 in additional analyses)

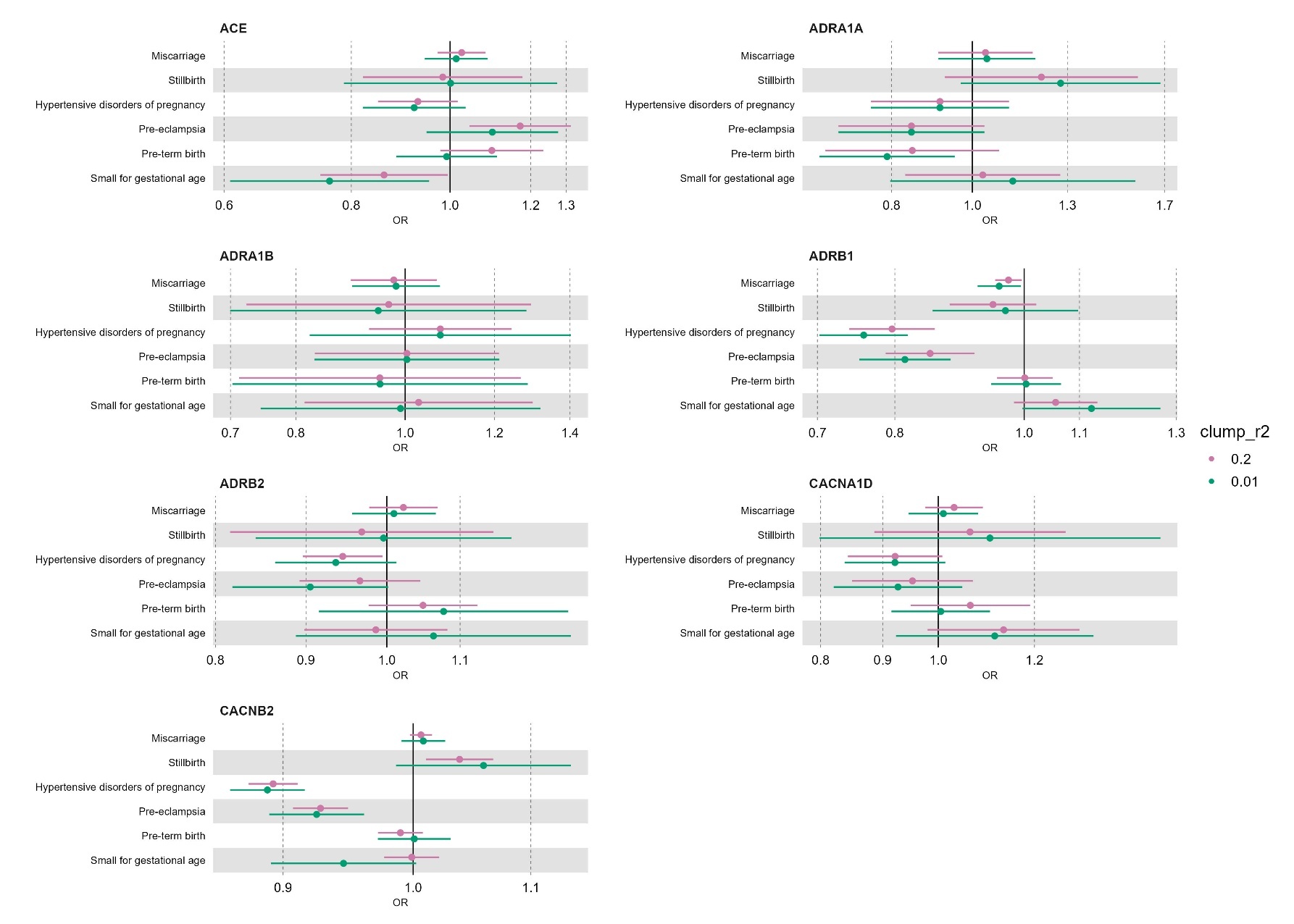

Results are expressed as odds ratio (OR) of outcome per 1 mmHg unit decrease in systolic or diastolic blood pressure (BP) due to genetically-proxied downregulation of the drug target gene. Point estimates and 95% confidence intervals are represented by circles and bars, respectively. Comparison restricted to drug targets for which more than one genetic variant was selected in the main analyses.

ACE: Angiotensin-converting enzyme; ADRA1A: adrenergic receptor α1A; ADRA1B: adrenergic receptor α1B; ADRA1D: adrenergic receptor α1D; ADRB1: adrenergic receptor β1; ADRB2: adrenergic receptor β2; AGTR1: angiotensin II receptor type 1; CACNA1C: Voltage-dependent L-type calcium channel subunit α1C; CACNA1D: Voltage-dependent L-type calcium channel subunit α1D; CACNB2: Voltage-dependent L-type calcium channel subunit β2; SLC12A3: Solute carrier family 12 member 3.

**Supplementary figure 5B**. MR estimates for the effect of modulating hypertension drug targets on secondary (binary) pregnancy-related outcomes using different linkage disequilibrium thresholds for selecting genetic variants (R^2^ < 0.20 in the main analyses and R^2^ < 0.01 in additional analyses)

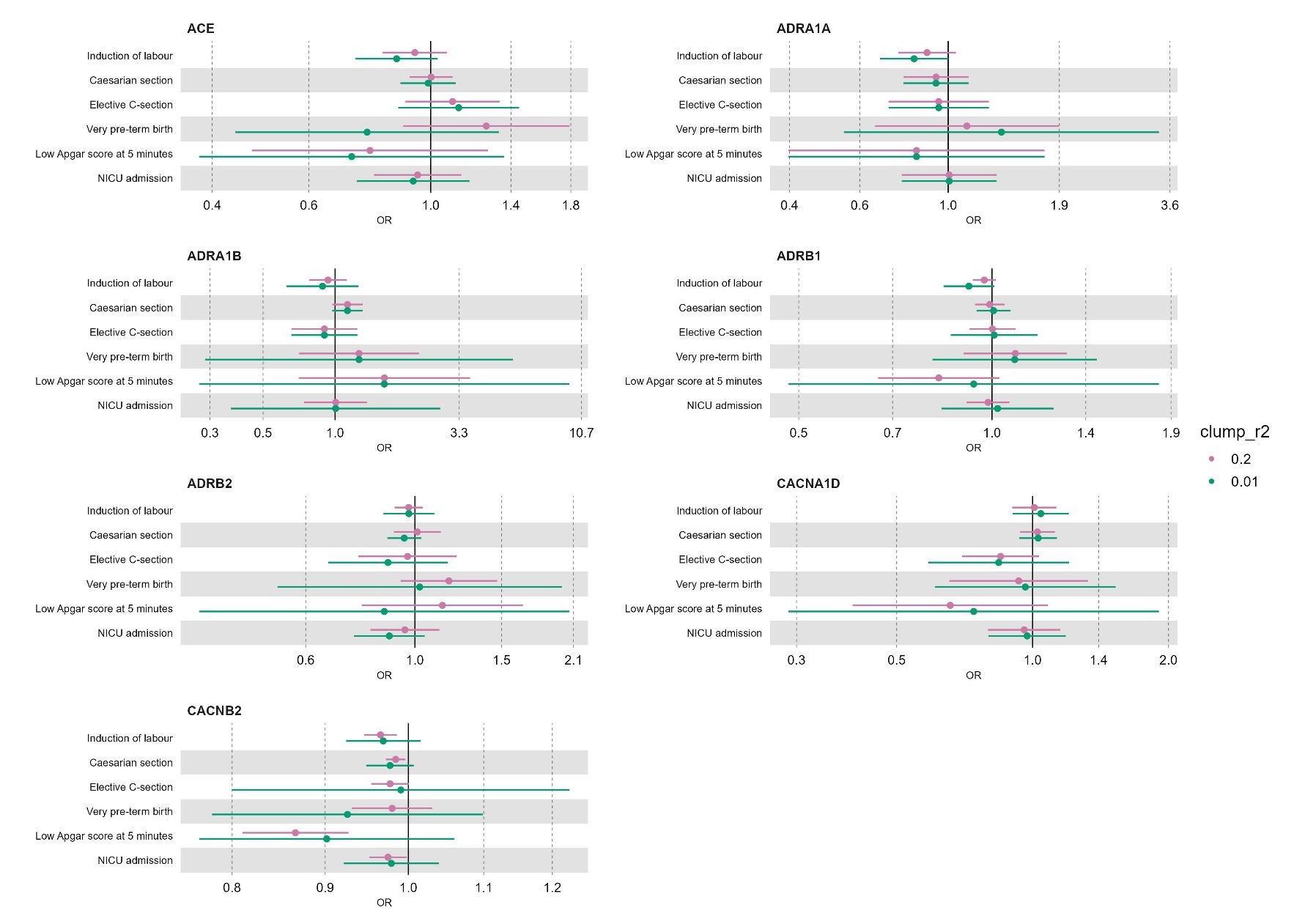

Results are expressed as odds ratio (OR) of outcome per 1 mmHg unit decrease in systolic or diastolic blood pressure (BP) due to genetically-proxied downregulation of the drug target gene. Point estimates and 95% confidence intervals are represented by circles and bars, respectively. Comparison restricted to drug targets for which more than one genetic variant was selected in the main analyses.

ACE: Angiotensin-converting enzyme; ADRA1A: adrenergic receptor α1A; ADRA1B: adrenergic receptor α1B; ADRA1D: adrenergic receptor α1D; ADRB1: adrenergic receptor β1; ADRB2: adrenergic receptor β2; AGTR1: angiotensin II receptor type 1; CACNA1C: Voltage-dependent L-type calcium channel subunit α1C; CACNA1D: Voltage-dependent L-type calcium channel subunit α1D; CACNB2: Voltage-dependent L-type calcium channel subunit β2; SLC12A3: Solute carrier family 12 member 3.

**Supplementary figure 5C**. MR estimates for the effect of modulating hypertension drug targets on secondary (binary) pregnancy-related outcomes using different linkage disequilibrium thresholds for selecting genetic variants (R^2^ < 0.20 in the main analyses and R^2^ < 0.01 in additional analyses)

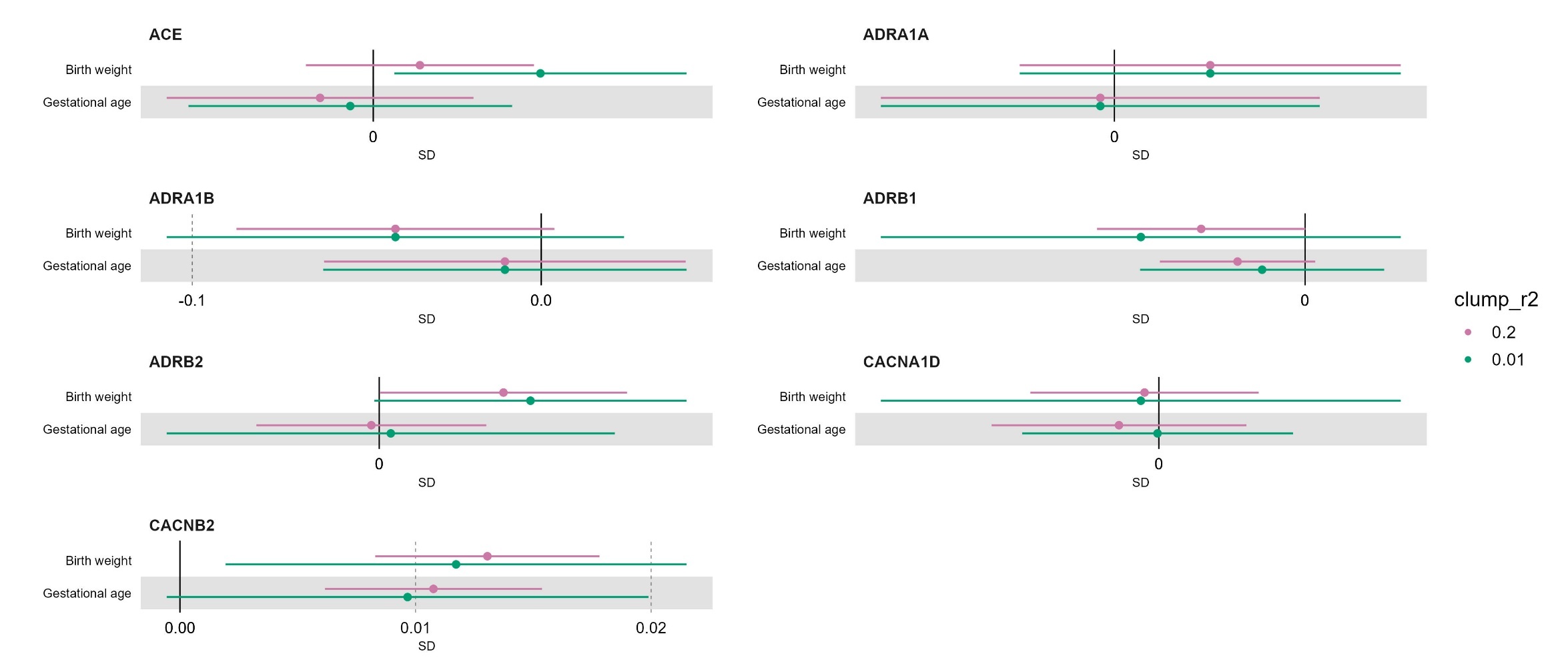

Results are expressed as mean difference in standard deviation (SD) units of outcome per 1 mmHg unit decrease in systolic or diastolic blood pressure (BP) due to genetically-proxied downregulation of the drug target gene. Point estimates and 95% confidence intervals are represented by circles and bars, respectively. Comparison restricted to drug targets for which more than one genetic variant was selected in the main analyses.

ACE: Angiotensin-converting enzyme; ADRA1A: adrenergic receptor α1A; ADRA1B: adrenergic receptor α1B; ADRA1D: adrenergic receptor α1D; ADRB1: adrenergic receptor β1; ADRB2: adrenergic receptor β2; AGTR1: angiotensin II receptor type 1; CACNA1C: Voltage-dependent L-type calcium channel subunit α1C; CACNA1D: Voltage-dependent L-type calcium channel subunit α1D; CACNB2: Voltage-dependent L-type calcium channel subunit β2; SLC12A3: Solute carrier family 12 member 3.

**Supplementary figure 6A**. Genetic association plots for blood pressure and pregnancy related outcomes for the *ADRA1D* genomic region

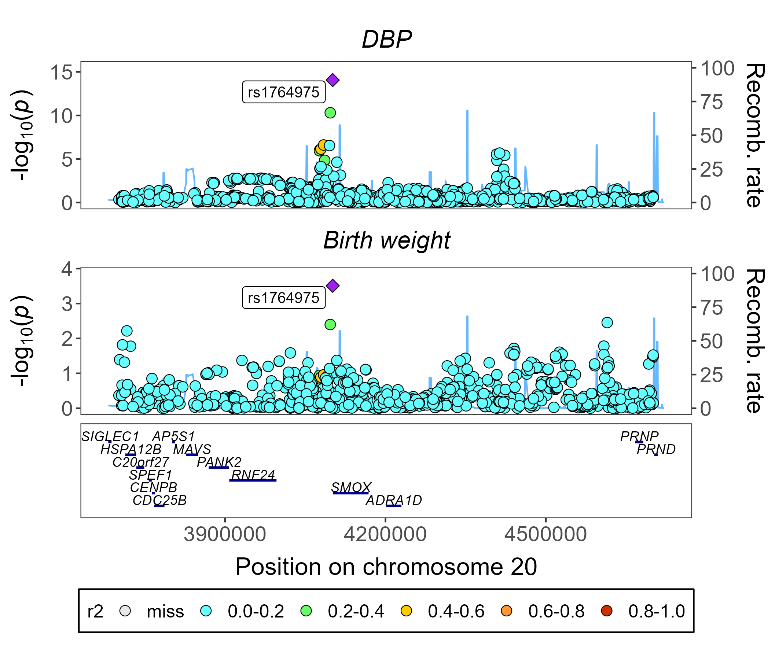

**Supplementary figure 6B**. Genetic association plots for blood pressure and pregnancy related outcomes for the *ACE* genomic region

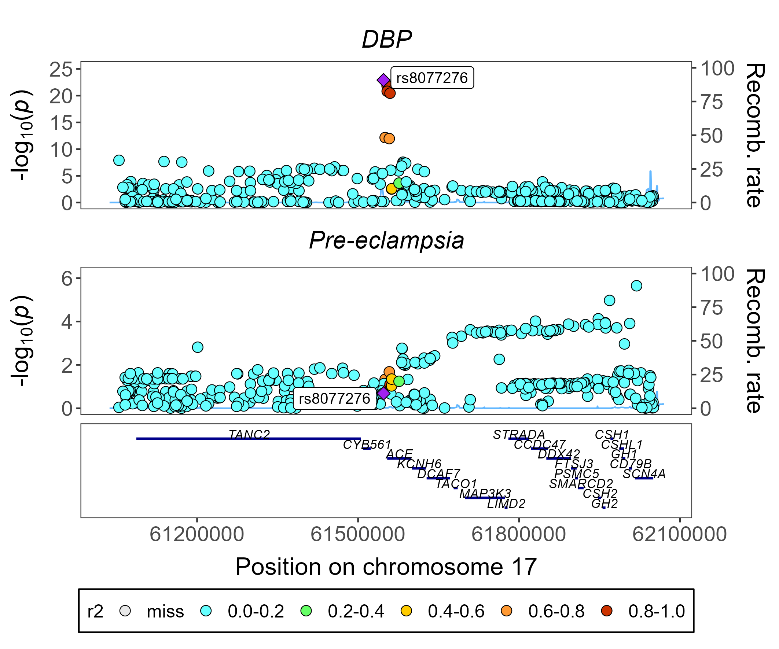

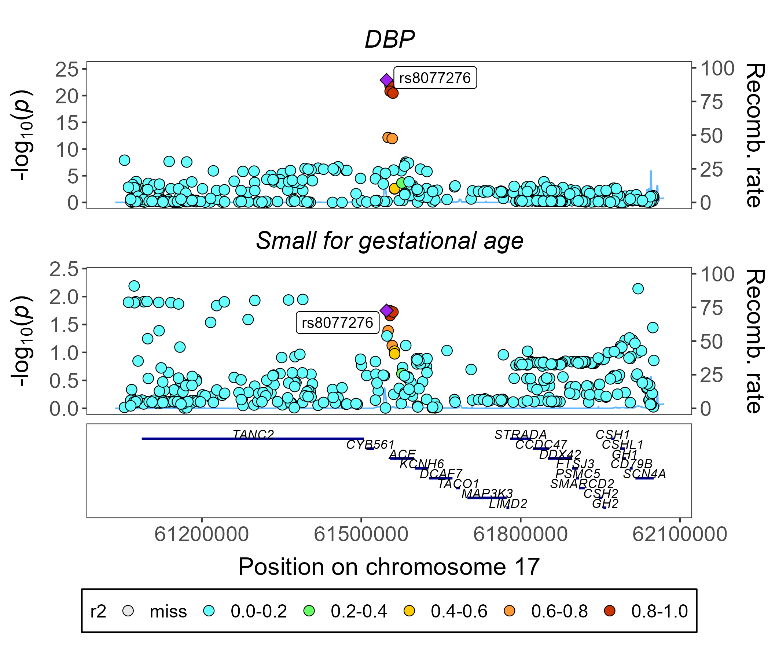

**Supplementary figure 6C**. Genetic association plots for blood pressure and pregnancy related outcomes for the *AGTR1* genomic region

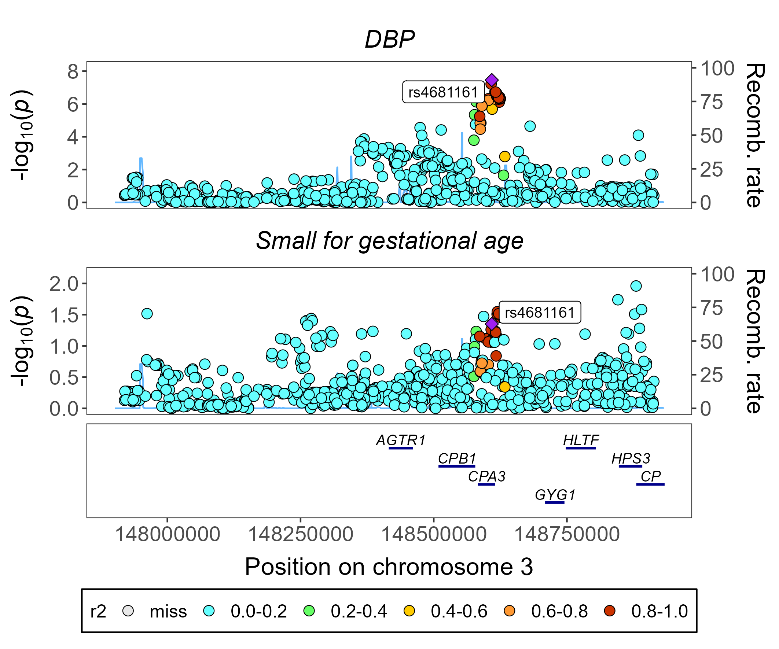

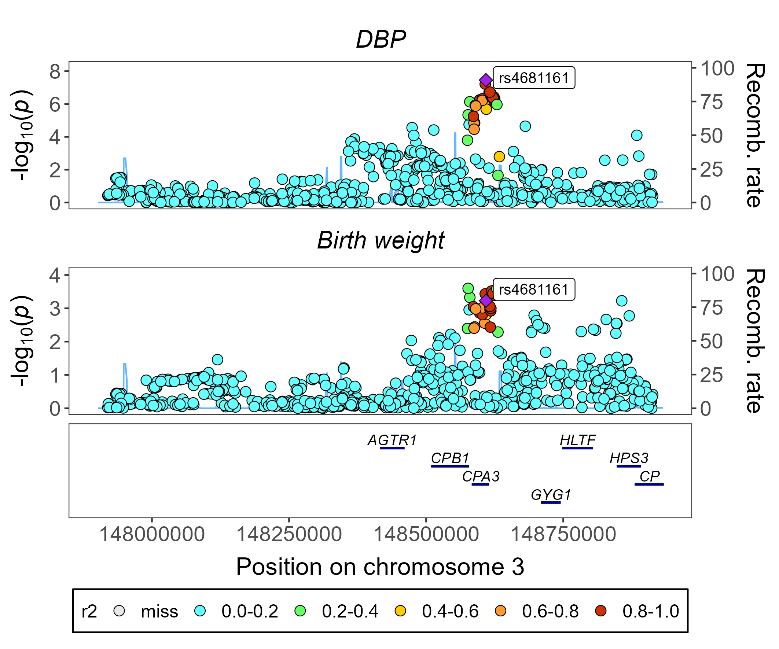

**Supplementary figure 6D**. Genetic association plots for blood pressure and pregnancy related outcomes for the *ADRB1* genomic region

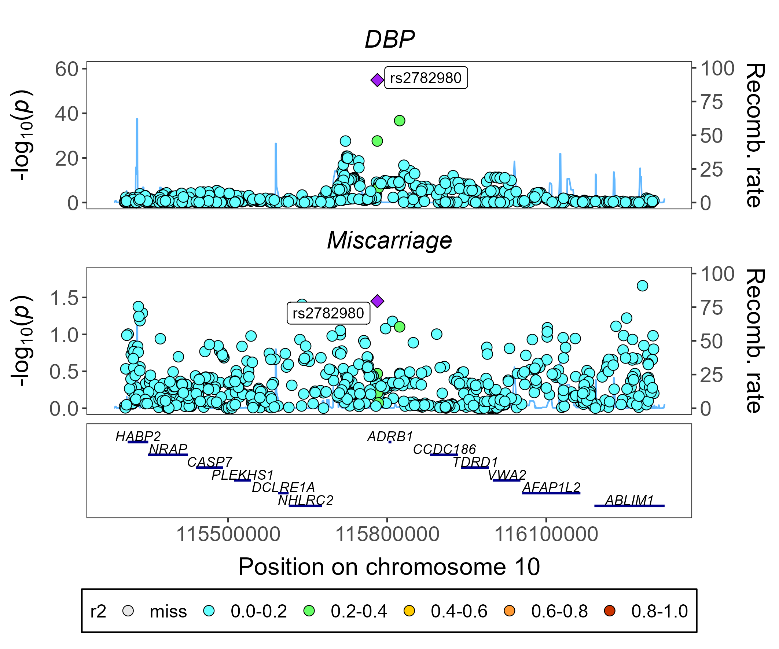

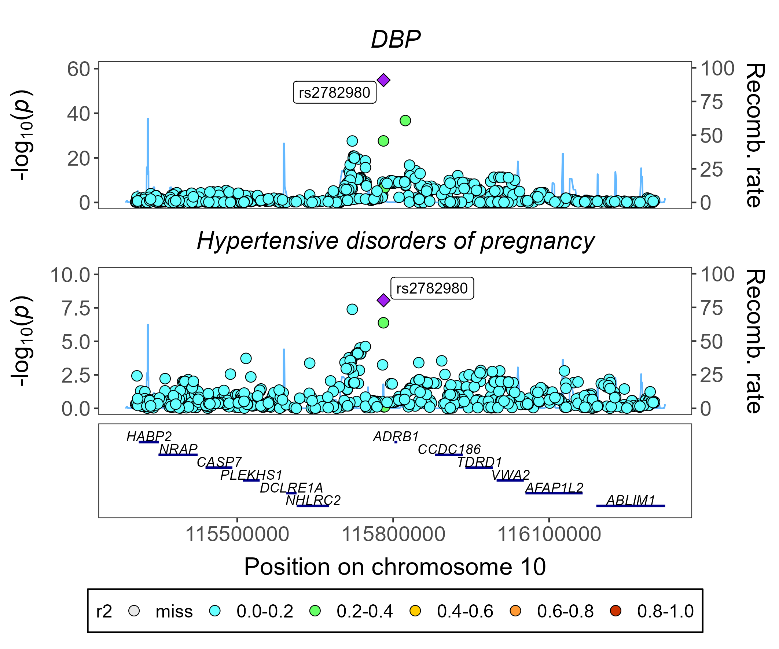

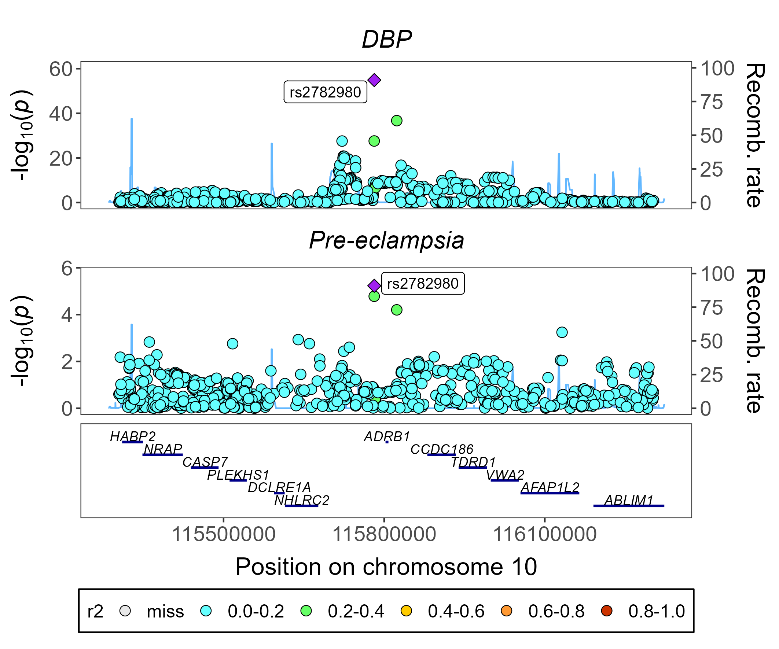

**Supplementary figure 6E**. Genetic association plots for blood pressure and pregnancy related outcomes for the *CACNA1C* genomic region

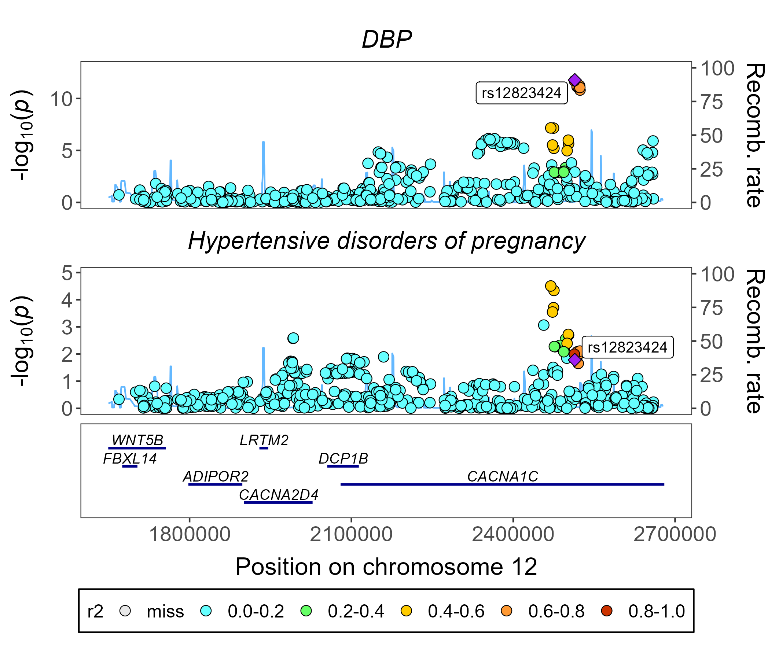

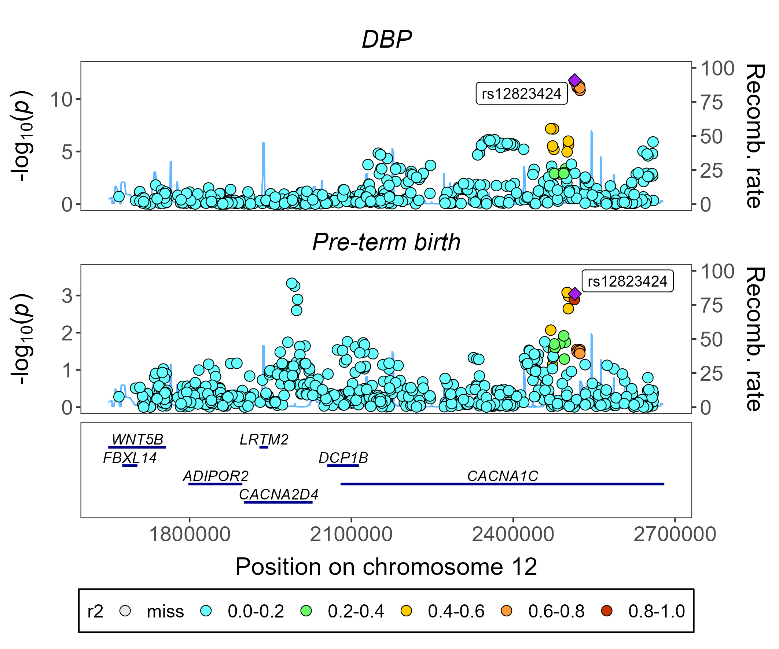

**Supplementary figure 6F**. Genetic association plots for blood pressure and pregnancy related outcomes for the *CACNAB2* genomic region

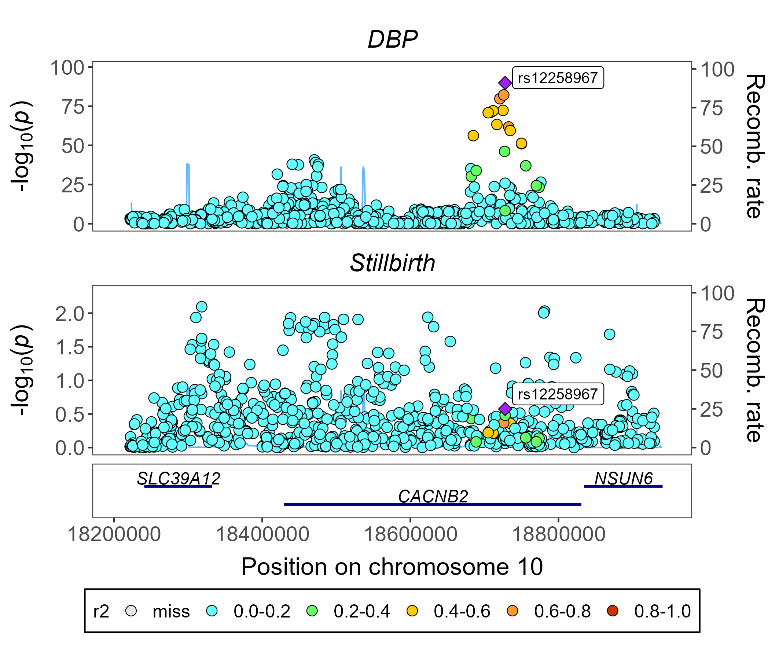

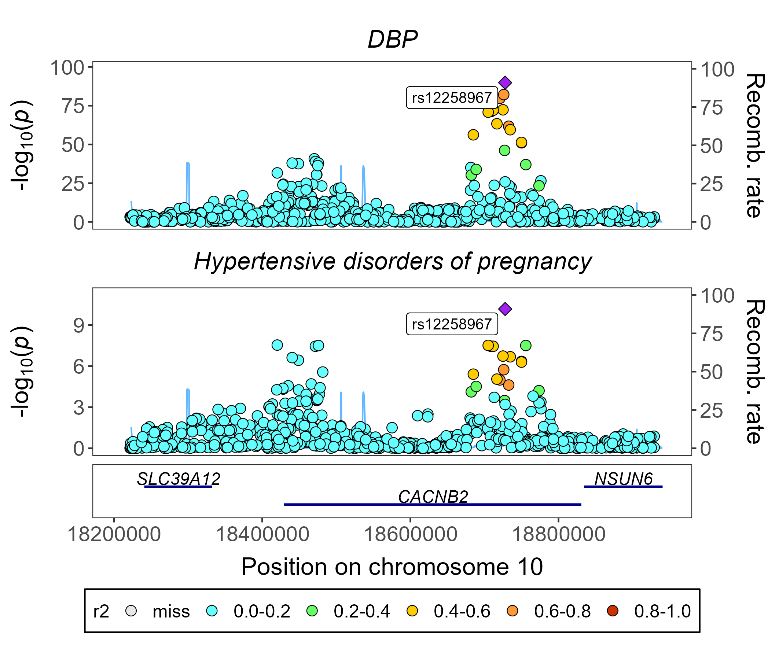

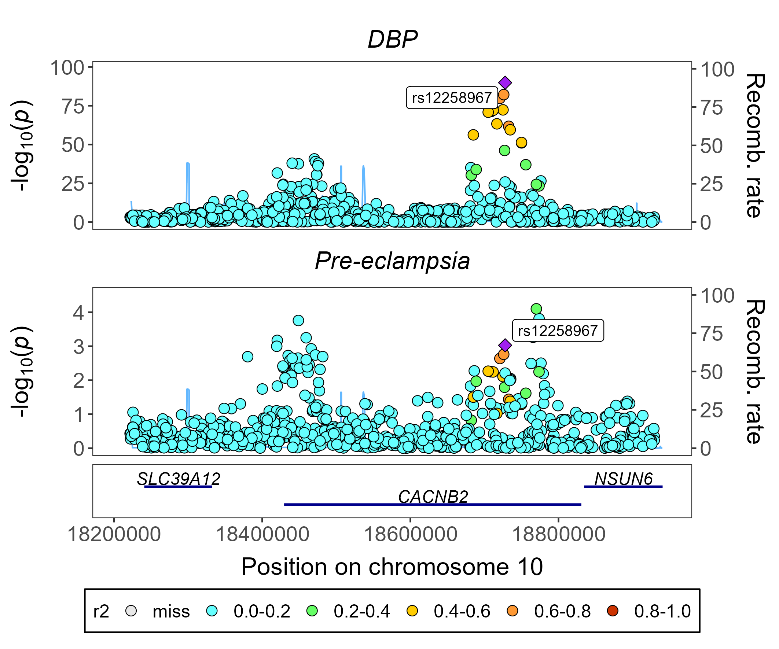

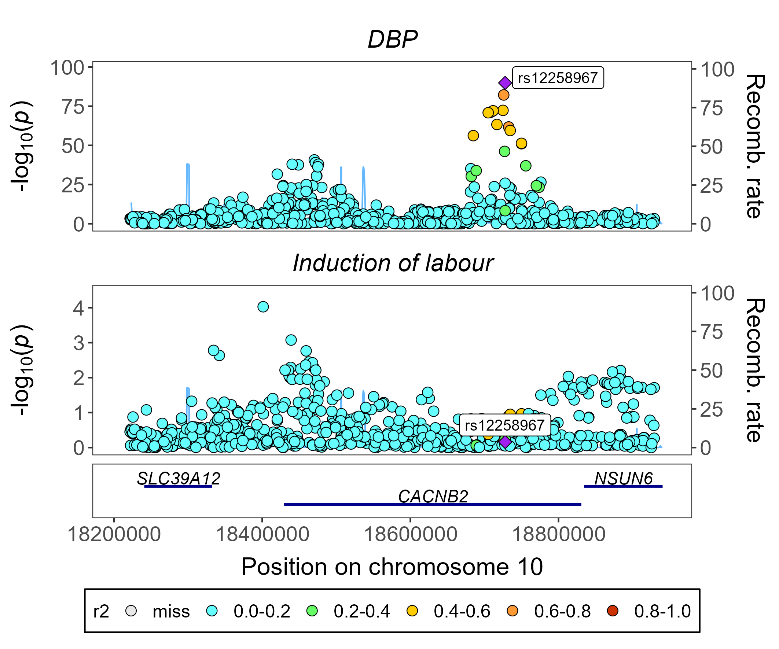

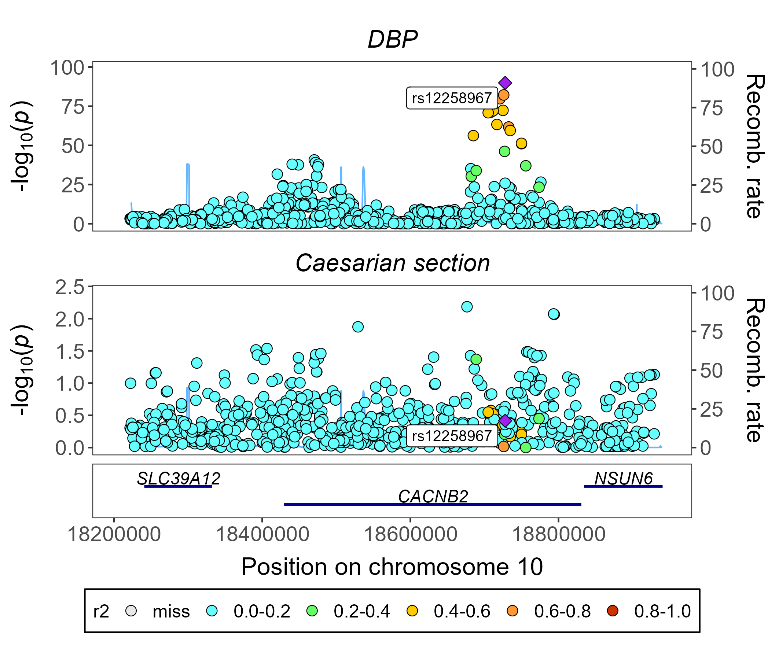

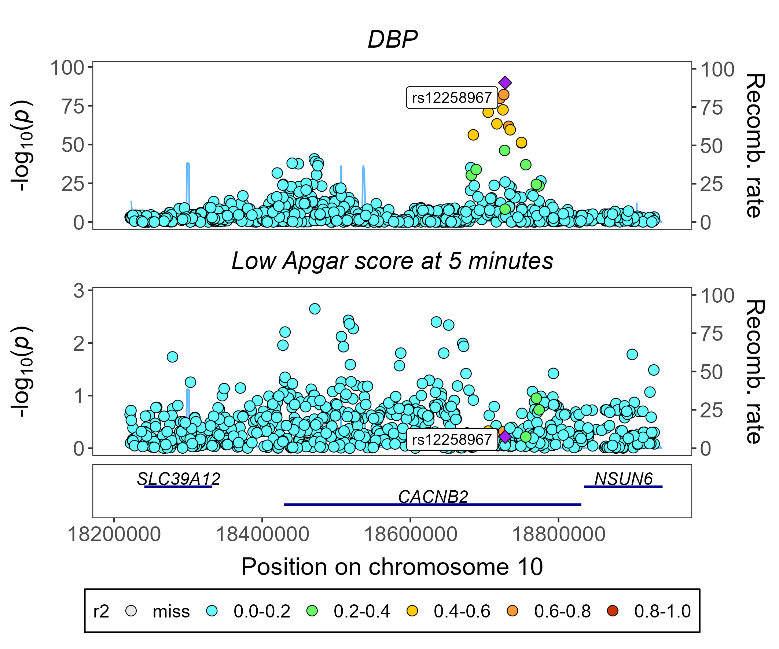

**Supplementary figure 6F (cont.)**. Genetic association plots for blood pressure and pregnancy related outcomes for the *CACNAB2* genomic region

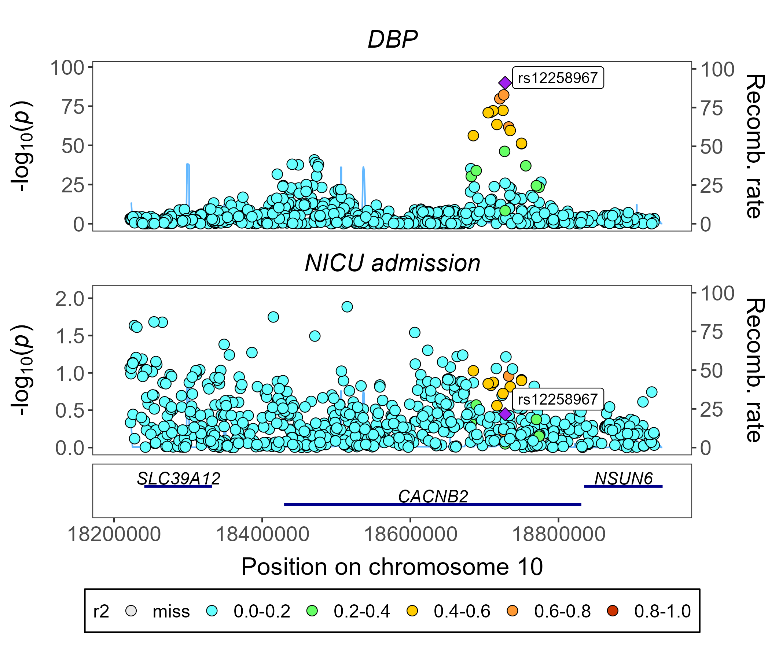

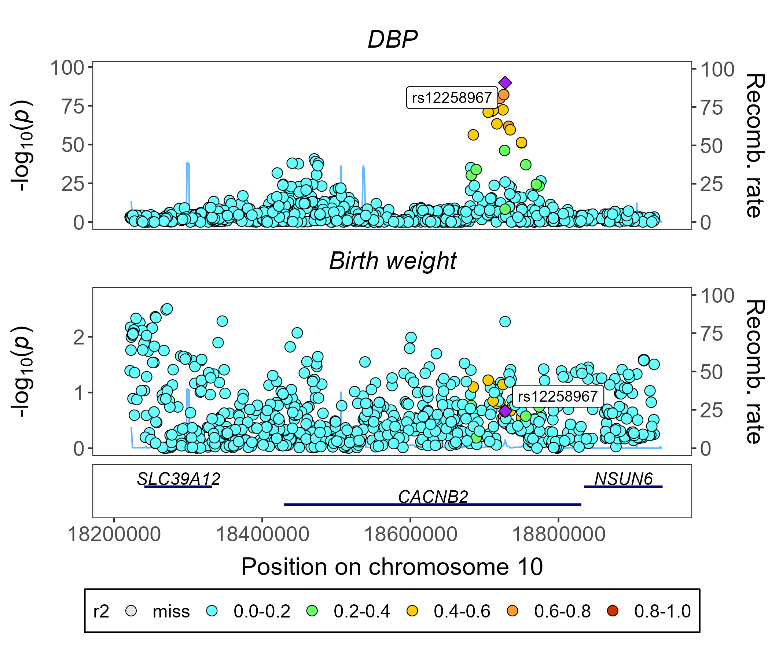

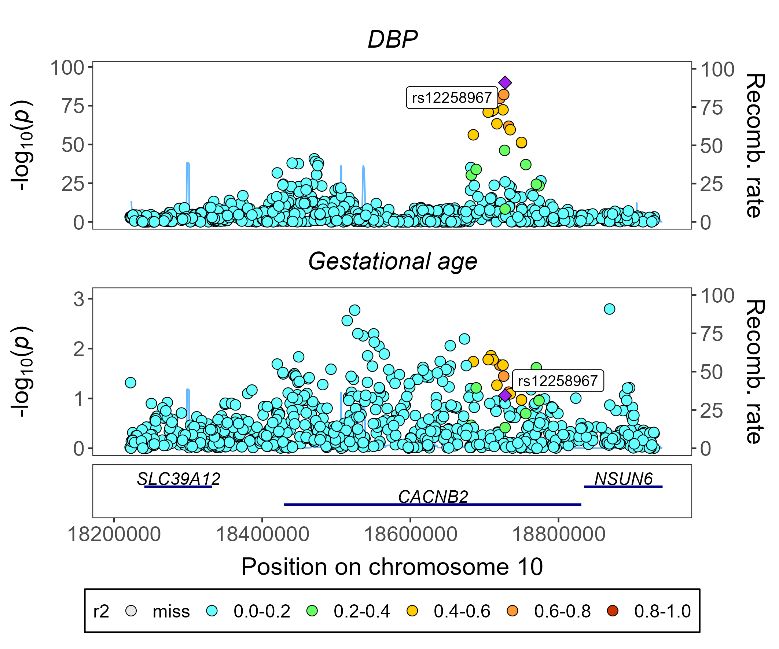

**Supplementary figure 6G**. Genetic association plots for blood pressure and pregnancy related outcomes for the *SLC12A3* genomic region

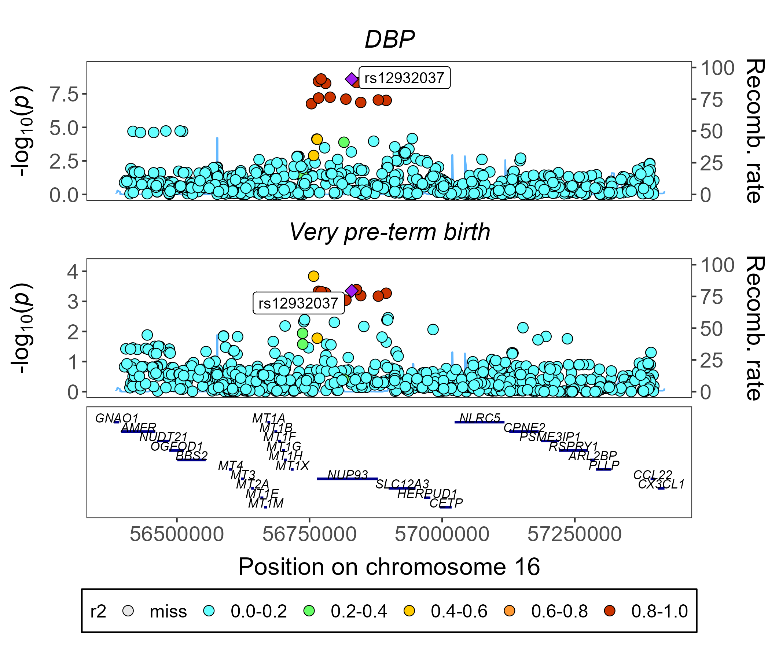

**Supplementary figure 7.** Mendelian randomization estimates for genetically-instrumented downregulation of drug targets on secondary continuous pregnancy-related outcomes based on maternal genetic effects (unadjusted and adjusted for fetal genotype) and fetal genetic effects (unadjusted and adjusted for maternal genotype)

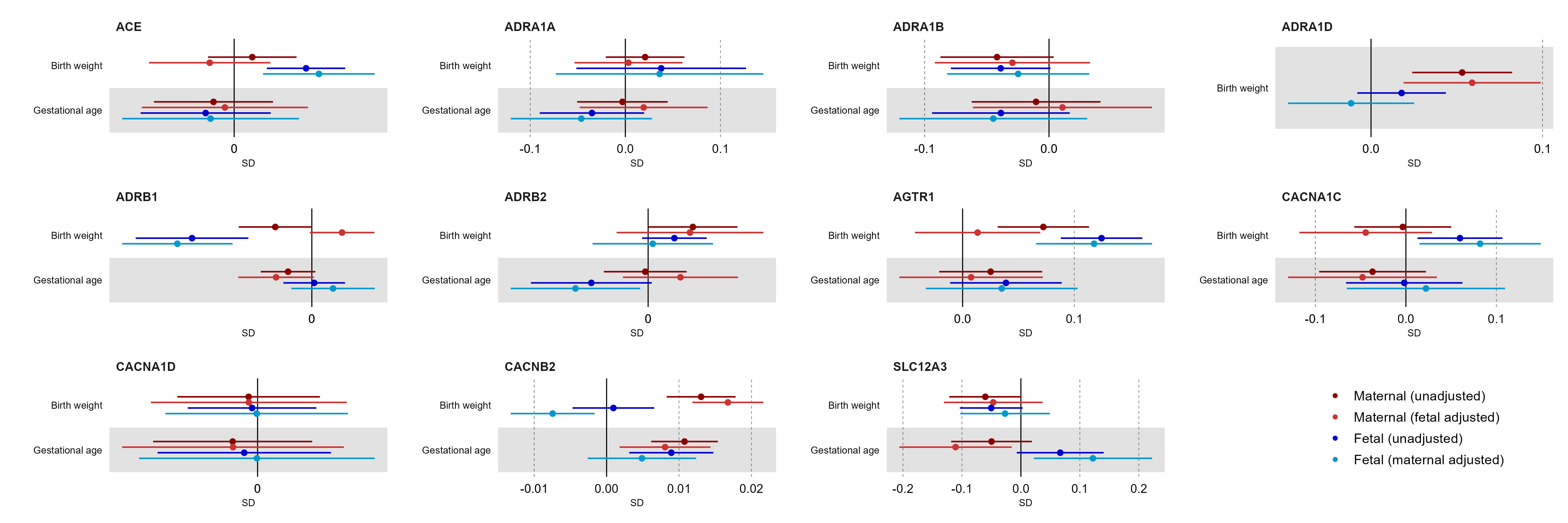
Results are expressed as mean difference in standard deviation (SD) unit (B) of outcome per 1mmHg unit decrease in systolic or diastolic blood pressure (BP) due to genetically-instrumented downregulation of the drug target gene. For comparison, we present the main results for maternal effects, unadjusted and unadjusted for offspring genotype, alongside results for fetal effects, unadjusted and unadjusted for maternal genotype. Point estimates and 95% confidence intervals are represented by circles and bars, respectively.

ACE: Angiotensin-converting enzyme; ADRA1A: adrenergic receptor α1A; ADRA1B: adrenergic receptor α1B; ADRA1D: adrenergic receptor α1D; ADRB1: adrenergic receptor β1; ADRB2: adrenergic receptor β2; AGTR1: angiotensin II receptor type 1; CACNA1C: Voltage-dependent L-type calcium channel subunit α1C; CACNA1D: Voltage-dependent L-type calcium channel subunit α1D; CACNB2: Voltage-dependent L-type calcium channel subunit β2; SLC12A3: Solute carrier family 12 member 3.

**Supplementary figure 8A.** Mendelian randomization estimates for genetically-instrumented downregulation of drug targets on primary pregnancy-related outcomes based on maternal genetic effects (unadjusted, adjusted for fetal or adjusted for fetal+paternal genotype), fetal genetic effects (unadjusted, adjusted for maternal or adjusted for maternal+paternal genotype), and paternal genetic effects (unadjusted or adjusted for fetal+maternal genotype)

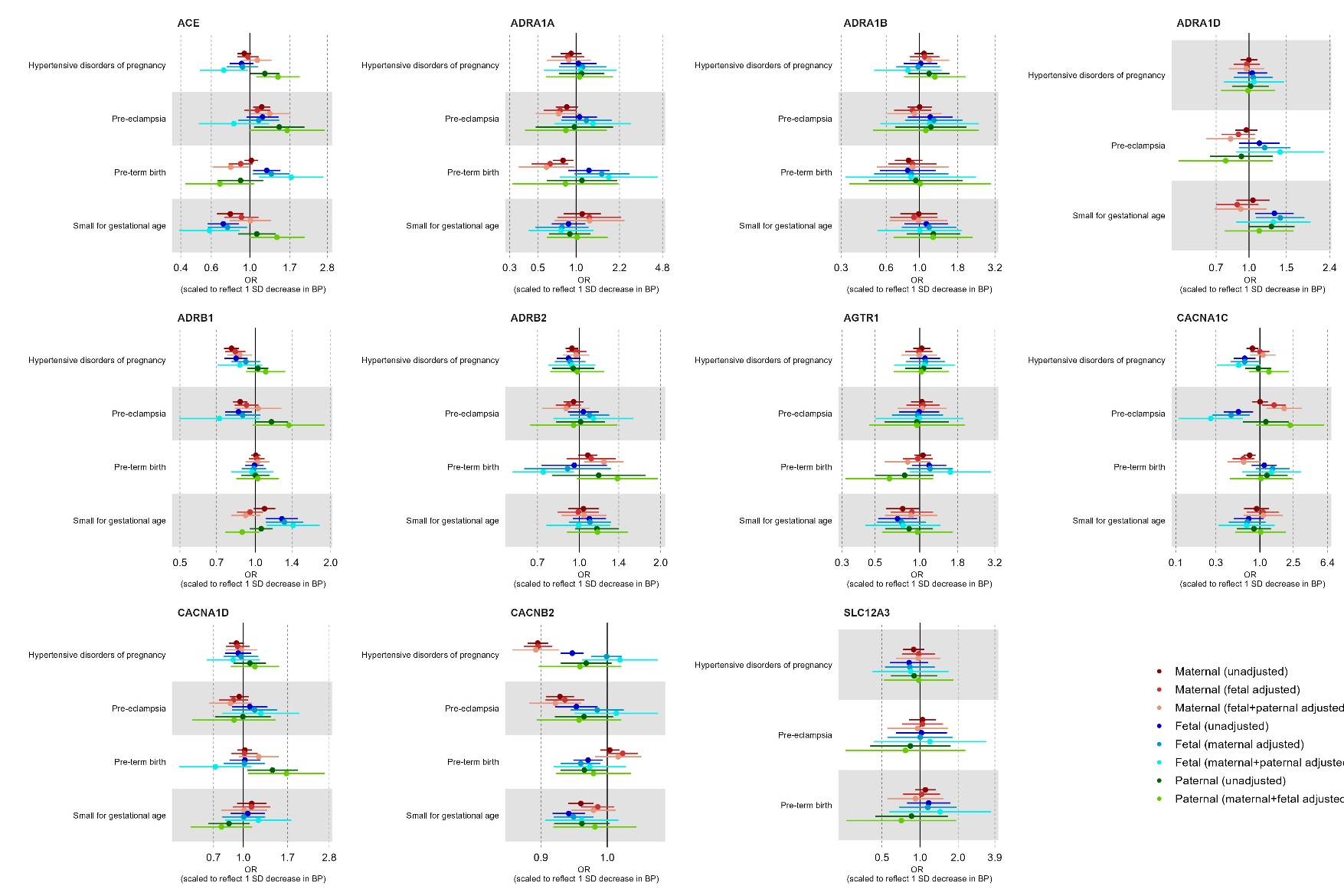

Results are expressed as odds ratio (OR) of outcome per 1mmHg unit decrease in systolic or diastolic blood pressure (BP) due to genetically-instrumented downregulation of the drug target gene. Point estimates and 95% confidence intervals are represented by circles and bars, respectively.

ACE: Angiotensin-converting enzyme; ADRA1A: adrenergic receptor α1A; ADRA1B: adrenergic receptor α1B; ADRA1D: adrenergic receptor α1D; ADRB1: adrenergic receptor β1; ADRB2: adrenergic receptor β2; AGTR1: angiotensin II receptor type 1; CACNA1C: Voltage-dependent L-type calcium channel subunit α1C; CACNA1D: Voltage-dependent L-type calcium channel subunit α1D; CACNB2: Voltage-dependent L-type calcium channel subunit β2; SLC12A3: Solute carrier family 12 member 3.

**Supplementary figure 8B.** Mendelian randomization estimates for genetically-instrumented downregulation of drug targets on secondary pregnancy-related outcomes based on maternal genetic effects (unadjusted, adjusted for fetal or adjusted for fetal+paternal genotype), fetal genetic effects (unadjusted, adjusted for maternal or adjusted for maternal+paternal genotype), and paternal genetic effects (unadjusted or adjusted for fetal+maternal genotype)

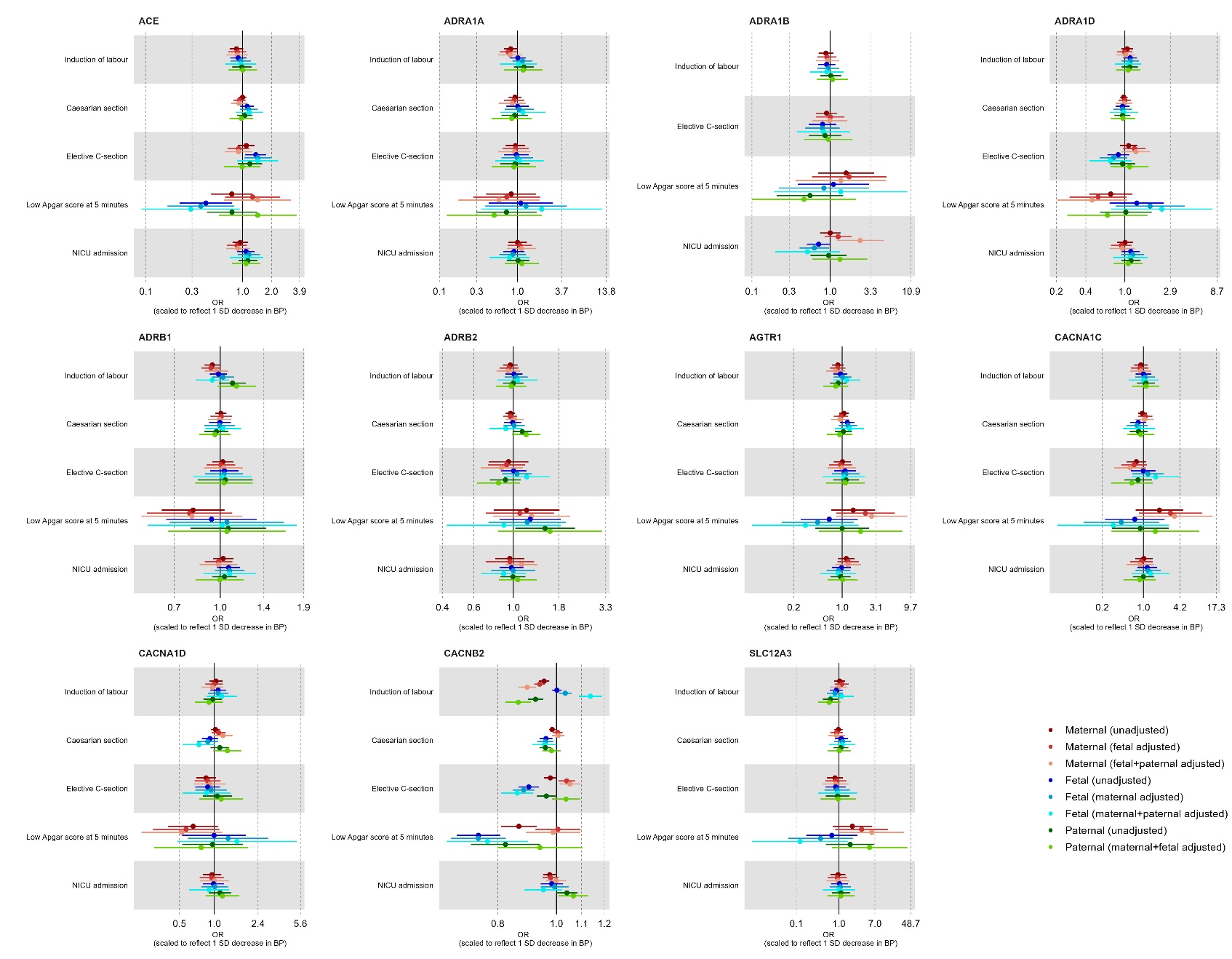

Results are expressed as odds ratio (OR) of outcome per 1mmHg unit decrease in systolic or diastolic blood pressure (BP) due to genetically-instrumented downregulation of the drug target gene. Point estimates and 95% confidence intervals are represented by circles and bars, respectively.

ACE: Angiotensin-converting enzyme; ADRA1A: adrenergic receptor α1A; ADRA1B: adrenergic receptor α1B; ADRA1D: adrenergic receptor α1D; ADRB1: adrenergic receptor β1; ADRB2: adrenergic receptor β2; AGTR1: angiotensin II receptor type 1; CACNA1C: Voltage-dependent L-type calcium channel subunit α1C; CACNA1D: Voltage-dependent L-type calcium channel subunit α1D; CACNB2: Voltage-dependent L-type calcium channel subunit β2; SLC12A3: Solute carrier family 12 member 3.

**Supplementary figure 8C.** Mendelian randomization estimates for genetically-instrumented downregulation of drug targets on secondary continuous pregnancy-related outcomes based on maternal genetic effects (unadjusted, adjusted for fetal or adjusted for fetal+paternal genotype), fetal genetic effects (unadjusted, adjusted for maternal or adjusted for maternal+paternal genotype), and paternal genetic effects (unadjusted or adjusted for fetal+maternal genotype)

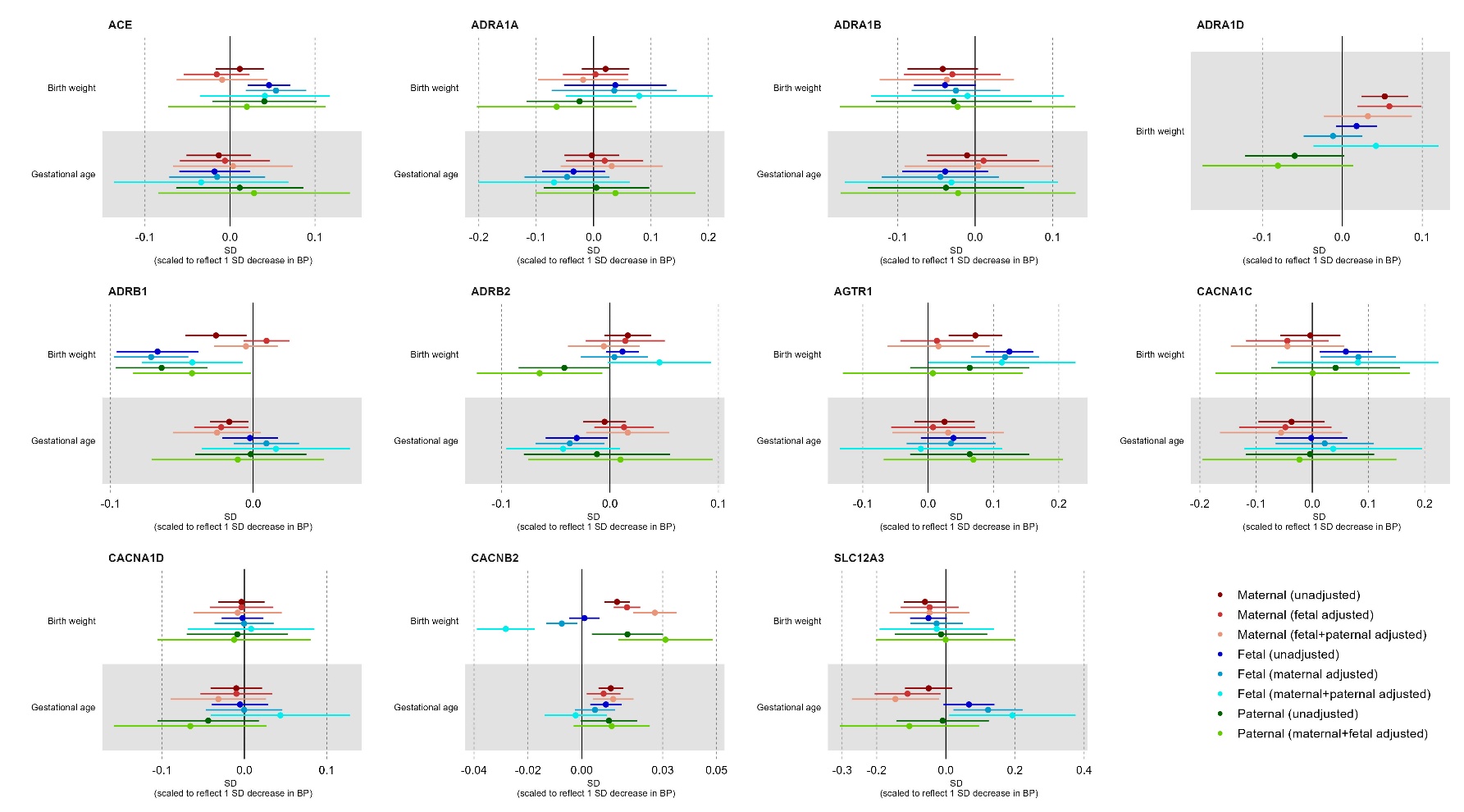

Results are expressed as mean standard deviation (SD) unit per 1mmHg unit decrease in systolic or diastolic blood pressure (BP) due to genetically-instrumented downregulation of the drug target gene. Point estimates and 95% confidence intervals are represented by circles and bars, respectively.

ACE: Angiotensin-converting enzyme; ADRA1A: adrenergic receptor α1A; ADRA1B: adrenergic receptor α1B; ADRA1D: adrenergic receptor α1D; ADRB1: adrenergic receptor β1; ADRB2: adrenergic receptor β2; AGTR1: angiotensin II receptor type 1; CACNA1C: Voltage-dependent L-type calcium channel subunit α1C; CACNA1D: Voltage-dependent L-type calcium channel subunit α1D; CACNB2: Voltage-dependent L-type calcium channel subunit β2; SLC12A3: Solute carrier family 12 member 3.

**Supplementary figure 9A**. MR estimates for the effect of modulating hypertension drug targets on primary (binary) pregnancy-related outcomes using standard methods (main analyses), MR-Link2 and GSMR2 to account for horizontal pleiotropy (sensitivity analyses)

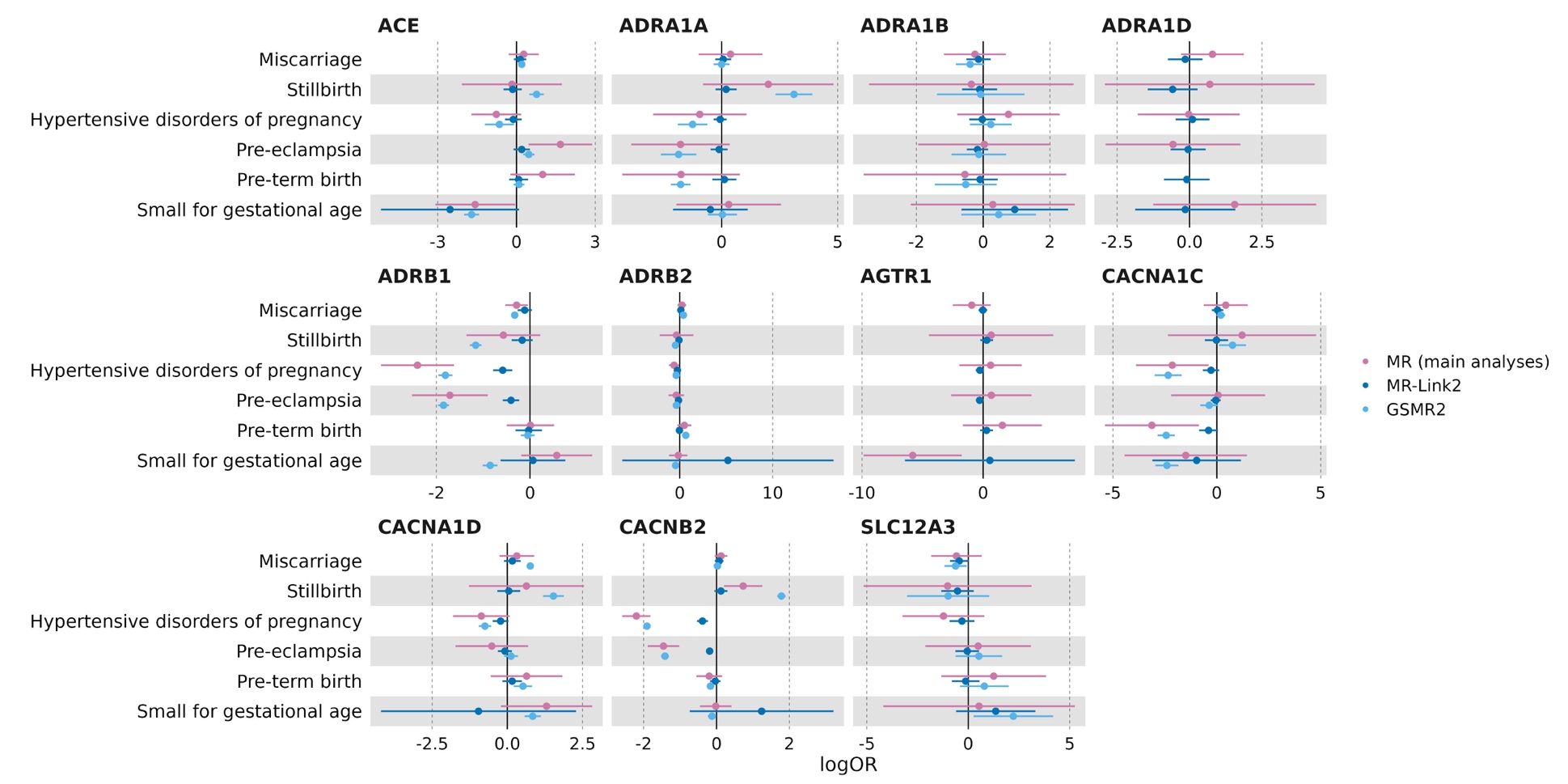

Results are expressed as log odds ratio (main analyses and GSMR2) or variance-explained units (MR-Link2) of primary pregnancy-related outcomes per 1mmHg decrease in systolic or diastolic blood pressure (BP) due to genetically-proxied downregulation of the drug target gene. Point estimates and 95% confidence intervals are represented by circles and bars, respectively. For some target-outcome pairs, GSMR2 selected too few variants, and estimates for this method are therefore unavailable.

ACE: Angiotensin-converting enzyme; ADRA1A: adrenergic receptor α1A; ADRA1B: adrenergic receptor α1B; ADRA1D: adrenergic receptor α1D; ADRB1: adrenergic receptor β1; ADRB2: adrenergic receptor β2; AGTR1: angiotensin II receptor type 1; CACNA1C: Voltage-dependent L-type calcium channel subunit α1C; CACNA1D: Voltage-dependent L-type calcium channel subunit α1D; CACNB2: Voltage-dependent L-type calcium channel subunit β2; SLC12A3: Solute carrier family 12 member 3.

**Supplementary figure 9B**. MR estimates for the effect of modulating hypertension drug targets on secondary (binary) pregnancy-related outcomes using standard methods (main analyses), MR-Link2 and GSMR2 to account for horizontal pleiotropy (sensitivity analyses)

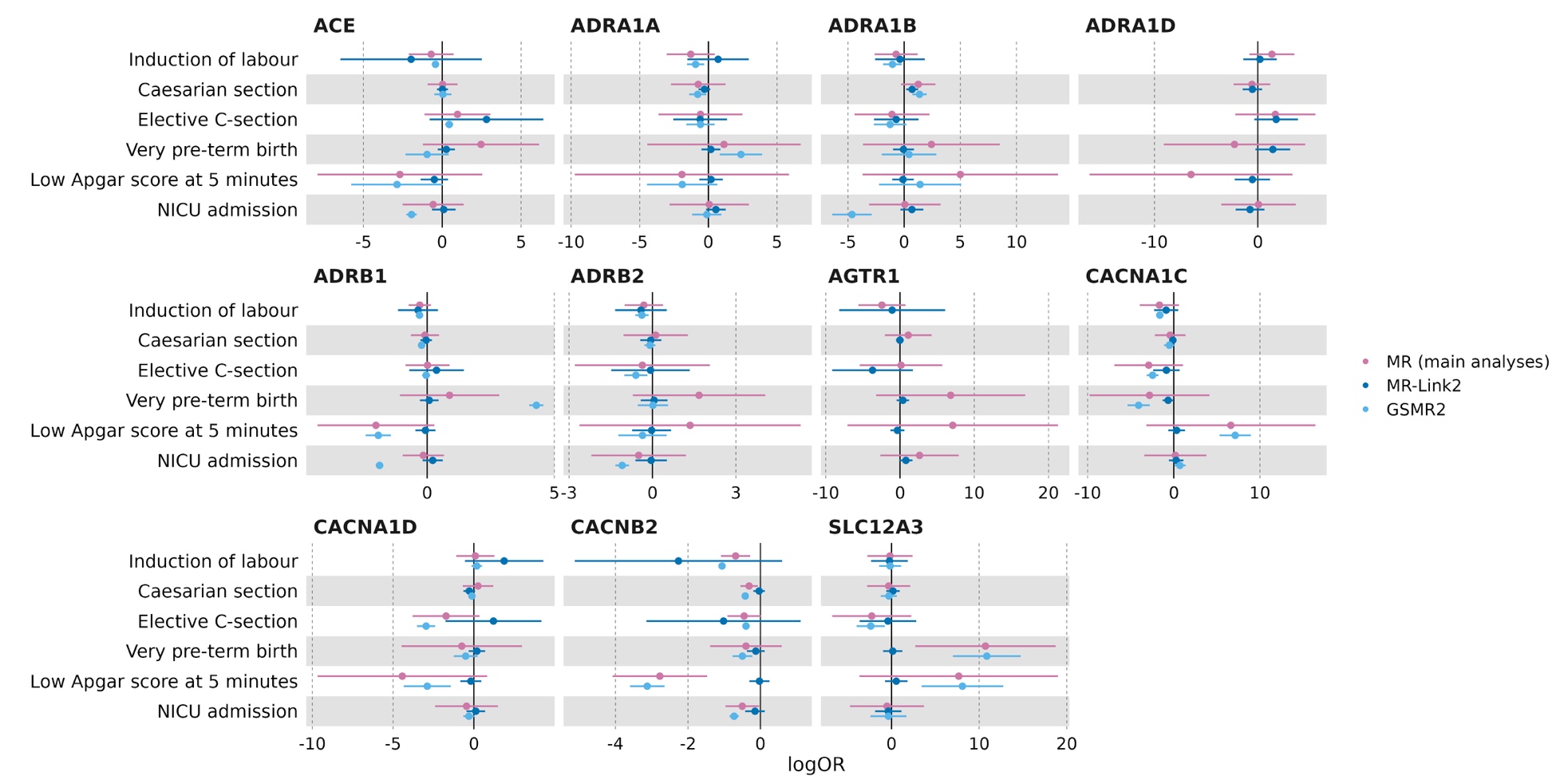

Results are expressed as log odds ratio (main analyses and GSMR2) or variance-explained units (MR-Link2) of secondary binary pregnancy-related outcomes per 1mmHg decrease in systolic or diastolic blood pressure (BP) due to genetically-proxied downregulation of the drug target gene. Point estimates and 95% confidence intervals are represented by circles and bars, respectively. For some target-outcome pairs, GSMR2 selected too few variants, and estimates for this method are therefore unavailable.

ACE: Angiotensin-converting enzyme; ADRA1A: adrenergic receptor α1A; ADRA1B: adrenergic receptor α1B; ADRA1D: adrenergic receptor α1D; ADRB1: adrenergic receptor β1; ADRB2: adrenergic receptor β2; AGTR1: angiotensin II receptor type 1; CACNA1C: Voltage-dependent L-type calcium channel subunit α1C; CACNA1D: Voltage-dependent L-type calcium channel subunit α1D; CACNB2: Voltage-dependent L-type calcium channel subunit β2; SLC12A3: Solute carrier family 12 member 3.

**Supplementary figure 9C**. MR estimates for the effect of modulating hypertension drug targets on secondary (continuous) pregnancy-related outcomes using standard methods (main analyses), MR-Link2 and GSMR2 to account for horizontal pleiotropy (sensitivity analyses)

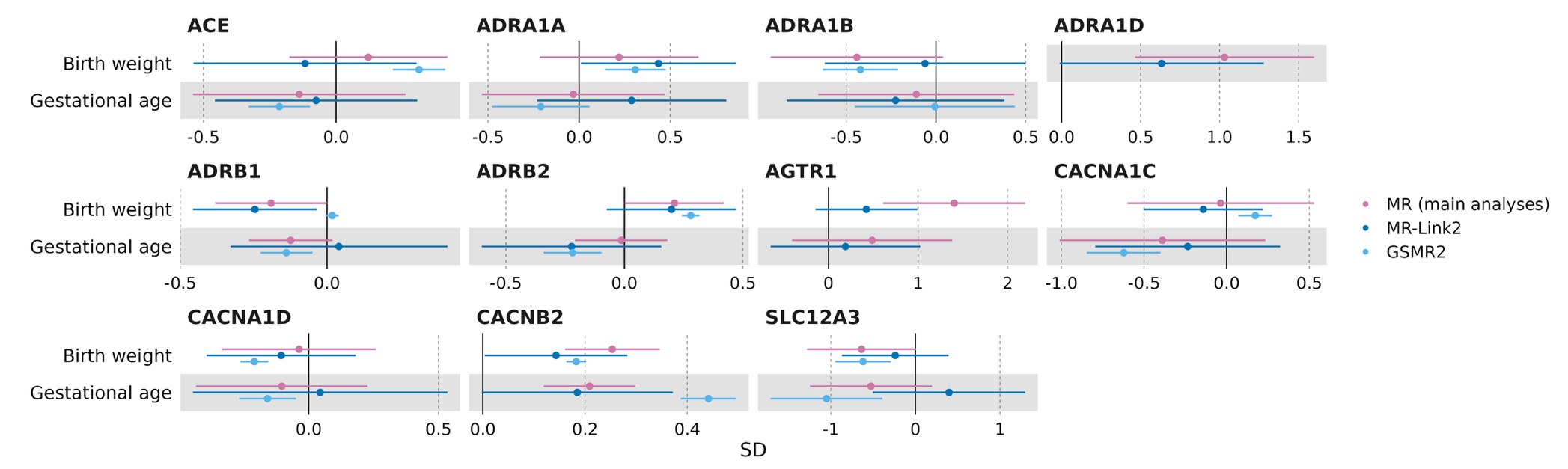

Results are expressed as SD units (main analyses and GSMR2) or variance-explained units (MR-Link2) of secondary continuous pregnancy-related outcomes per 1mmHg decrease in systolic or diastolic blood pressure (BP) due to genetically-proxied downregulation of the drug target gene. Point estimates and 95% confidence intervals are represented by circles and bars, respectively. For some target-outcome pairs, GSMR2 selected too few variants, and estimates for this method are therefore unavailable.

ACE: Angiotensin-converting enzyme; ADRA1A: adrenergic receptor α1A; ADRA1B: adrenergic receptor α1B; ADRA1D: adrenergic receptor α1D; ADRB1: adrenergic receptor β1; ADRB2: adrenergic receptor β2; AGTR1: angiotensin II receptor type 1; CACNA1C: Voltage-dependent L-type calcium channel subunit α1C; CACNA1D: Voltage-dependent L-type calcium channel subunit α1D; CACNB2: Voltage-dependent L-type calcium channel subunit β2; SLC12A3: Solute carrier family 12 member 3

**Supplementary figure 10**. MR estimates for the effect of modulating hypertension drug targets on positive control outcomes using standard methods (main analyses), and MR-Link2 and GSMR2 to account for horizontal pleiotropy (sensitivity analyses)

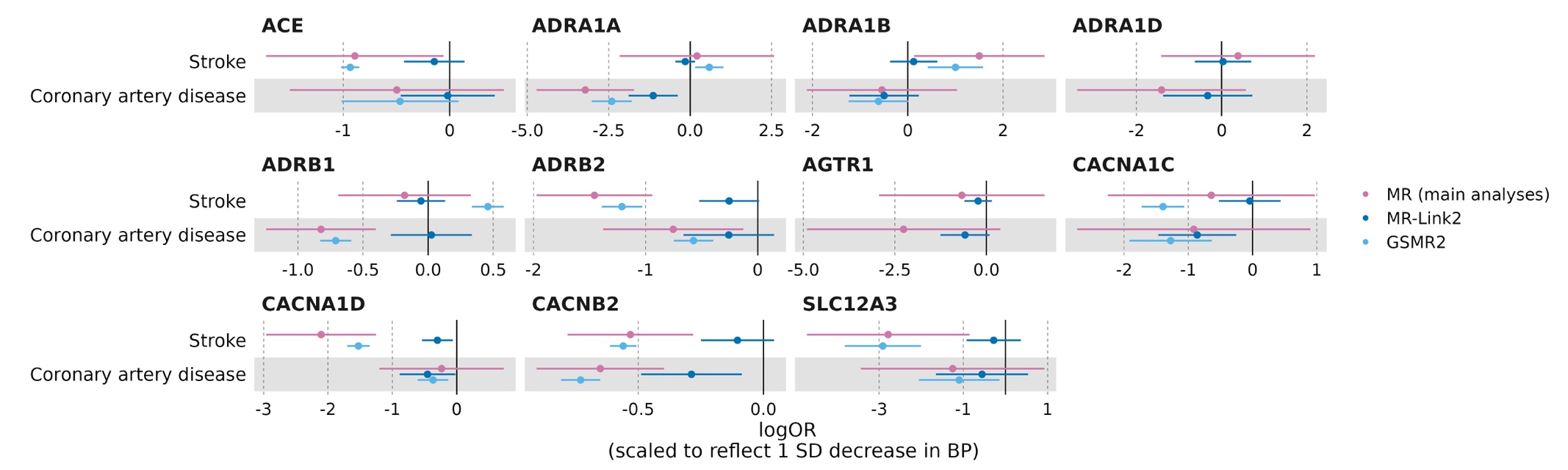

Results are expressed as log odds ratio (main analyses and GSMR2) or variance-explained units (MR-Link2) of positive control outcomes per 1mmHg decrease in systolic or diastolic blood pressure (BP) due to genetically-proxied downregulation of the drug target gene. Point estimates and 95% confidence intervals are represented by circles and bars, respectively. For some target-outcome pairs, GSMR2 selected too few variants, and estimates for this method are therefore unavailable.

ACE: Angiotensin-converting enzyme; ADRA1A: adrenergic receptor α1A; ADRA1B: adrenergic receptor α1B; ADRA1D: adrenergic receptor α1D; ADRB1: adrenergic receptor β1; ADRB2: adrenergic receptor β2; AGTR1: angiotensin II receptor type 1; CACNA1C: Voltage-dependent L-type calcium channel subunit α1C; CACNA1D: Voltage-dependent L-type calcium channel subunit α1D; CACNB2: Voltage-dependent L-type calcium channel subunit β2; SLC12A3: Solute carrier family 12 member 3
